# Birth weight, BMI in adulthood and latent autoimmune diabetes in adults: A Mendelian randomization study

**DOI:** 10.1101/2021.10.25.21265464

**Authors:** Yuxia Wei, Yiqiang Zhan, Josefin E. Löfvenborg, Tiinamaija Tuomi, Sofia Carlsson

## Abstract

**Aims:** Observational studies have found an increased risk of latent autoimmune diabetes in adults (LADA) associated with low birth weight and adult overweight/obesity. We aimed to investigate whether these associations are causal, using a two-sample Mendelian randomization (MR) design. In addition, we wanted to compare results for LADA and type 2 diabetes.

**Methods:** We identified 129 SNPs as instrumental variables (IVs) for birth weight from a genome-wide association study (GWAS) of the Early Growth Genetics Consortium (EGG) and the UK Biobank. We identified 820 SNPs as IVs for adult BMI from a GWAS of the UK Biobank and the Genetic Investigation of ANthropometric Traits consortium (GIANT). Summary statistics for the associations between IVs and LADA were extracted from the only GWAS involving 2,634 cases and 5,947 population controls. We used the inverse-variance weighted (IVW) estimator as our primary analysis, supplemented by a series of sensitivity analyses.

**Results:** Genetically determined birth weight was inversely associated with LADA (OR per SD [∼500 g] decrease in birth weight: 2.02, 95% CI: 1.37-2.97). In contrast, genetically predicted BMI in adulthood was positively associated with LADA (OR per SD [∼4.8 kg/m^2^] increase in BMI: 1.40, 95% CI: 1.14-1.71). Results persisted in a range of sensitivity analyses using other MR estimators or excluding some IVs. With respect to type 2 diabetes, the association with birth weight was not stronger than in LADA while the association with adult BMI was stronger than in LADA.

**Conclusions/ interpretation:** This study provides genetic support for a causal link between low birth weight, adult overweight/obesity, and LADA.

**Research in context:** - **What is already known about this subject? (maximum of 3 bullet points)** Previous Mendelian randomization studies have found that both lower birth weight and higher adult BMI are associated with an increased risk of type 2 diabetes. In contrast, the evidence on latent autoimmune diabetes in adults (LADA) is very limited. Only a few observational studies indicated an inverse association between birth weight and LADA, or a positive association between adult BMI and LADA.
- **What is the key question** Are birth weight and adult adiposity causally associated with LADA?
- **What are the new findings** Genetically determined birth weight was inversely associated with LADA while genetically predicted BMI in adulthood was positively associated with LADA. The association with birth weight was not weaker for LADA than for type 2 diabetes, while adult BMI had a greater impact on type 2 diabetes than on LADA.
- **How might this impact on clinical practice in the foreseeable future** Findings from this study indicate that measures should be taken to reduce the prevalence of adult overweight/obesity for the prevention of diabetes with and without an autoimmune component. The mechanism linking low birth weight to diabetes remains to be explored.

## Introduction

Latent autoimmune diabetes in adults (LADA) is a hybrid form of diabetes. Genetically, it is closely related to type 1 diabetes with a strong link to HLA genotypes[1, 2], while many of its clinical features such as metabolic syndrome are shared with type 2 diabetes[2]. LADA is characterized by pancreatic autoantibodies, slow progression to insulin dependence and is usually restricted to adult patients[2] although a similar phenomenon has been described in younger patients (LADY, Latent autoimmune diabetes in the young)[3]. Autoantibody testing is required to separate LADA from type 2 diabetes. Around 5–14% of adult patients diagnosed with “type 2 diabetes” in Europe, North America, and Asia have pancreatic autoantibodies[2].

Risk factors of type 2 diabetes are extensively studied[4, 5] and the disease can be prevented or postponed by maintaining a healthy weight and physical activity[6]. In comparison, the evidence on environmental/lifestyle risk factors of LADA is limited[7]. However, we have previously reported on an increased risk of LADA in relation to overweight/obesity [8] and low birth weight[9], in line with findings in type 2 diabetes[10, 11]. Observational studies are prone to residual confounding and reverse causation, while randomized control trials may be unfeasible in studying some risk factors of diseases. Individuals’ genotypes are randomly assigned from their parents before conception and thus are less likely to suffer from confounding or reverse causation[12]. Taking advantage of the natural experiments, Mendelian randomization (MR) studies use genetic variants as instrumental variables (IVs) for an environmentally modifiable exposure to make causal inference about the outcome[12].

Our aim was to investigate whether low birth weight and adult adiposity are implicated in the etiology of LADA by, for the first time, using a two-sample MR design. In addition, we wanted to compare these associations in LADA and type 2 diabetes.

## Methods

### Study design

This was a two-sample MR study using summary statistics from two separate genome-wide association studies (GWAS) of non-overlapping samples of the same underlying population; one provided measures of the associations between IVs and the exposure and the other on the associations between IVs and the outcome[13].

### Genetic instruments

#### Birth weight

Summary statistics (β coefficients and standard errors) for the associations between IVs and birth weight were extracted from the most recent meta-analysis of the Early Growth Genetics Consortium (EGG) and UK Biobank[14]. Information on birth weight had been collected by measurement at birth, obstetric records, medical registers, interviews with the mother, or self-report as adults in different included studies (**eTable 1**). The GWAS identified 145 independent SNPs associated with own birth weight at *P*<6.6×10^−9^ (genome-wide significance threshold revised by authors of the GWAS) in 298,140 European individuals. Among the 145 SNPs, 129 SNPs (**eTable 2**) were available in the GWAS dataset of LADA described below, explaining 2.61% of the variance in birth weight.

**Table 1.**
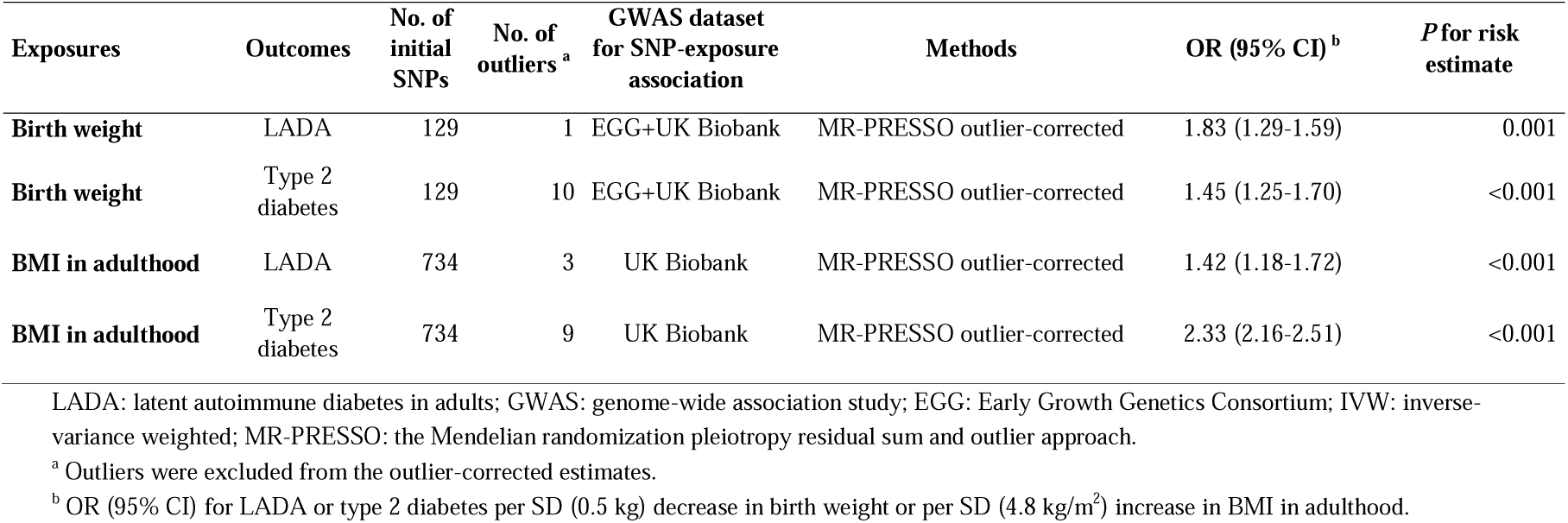
Comparison between LADA and type 2 diabetes.

| Exposures | Outcomes | No. of initial SNPs | No. of outliers<sup>a</sup> | GWAS dataset for SNP-exposure association | Methods | OR (95% CI)<sup>b</sup> | P for risk estimate |
| --- | --- | --- | --- | --- | --- | --- | --- |
| **Birth weight** | LADA | 129 | 1 | EGG+UK Biobank | MR-PRESSO outlier-corrected | 1.83 (1.29-1.59) | 0.001 |
| **Birth weight** | Type 2 diabetes | 129 | 10 | EGG+UK Biobank | MR-PRESSO outlier-corrected | 1.45 (1.25-1.70) | <0.001 |
| **BMI in adulthood** | LADA | 734 | 3 | UK Biobank | MR-PRESSO outlier-corrected | 1.42 (1.18-1.72) | <0.001 |
| **BMI in adulthood** | Type 2 diabetes | 734 | 9 | UK Biobank | MR-PRESSO outlier-corrected | 2.33 (2.16-2.51) | <0.001 |

LADA: latent autoimmune diabetes in adults; GWAS: genome-wide association study; EGG: Early Growth Genetics Consortium; IVW: inverse-variance weighted; MR-PRESSO: the Mendelian randomization pleiotropy residual sum and outlier approach.
<sup>a</sup> Outliers were excluded from the outlier-corrected estimates.
<sup>b</sup> OR (95% CI) for LADA or type 2 diabetes per SD (0.5 kg) decrease in birth weight or per SD (4.8 kg/m<sup>2</sup>) increase in BMI in adulthood.

#### Adult BMI

The primary set of IVs for BMI in adulthood was obtained from a meta-analysis including 681,275 individuals of European ancestry from UK Biobank and the Genetic Investigation of ANthropometric Traits (GIANT) consortium[15]. Information on BMI had mainly been obtained through clinical measurements (**eTable 1**). SNPs of this meta-analysis have been used as IVs for BMI in a previous MR study[16]. In the meta-analysis, independent SNPs were selected through LD clumping, in which SNPs with LD measure of r^2^>0.01 within a 10000-kb window were pruned, resulting in a total of 839 independent SNPs with *P* < 1 × 10^−8^ (genome-wide significance threshold revised by authors of the meta-analysis). Of these, 817 SNPs were available in the GWAS of LADA and proxy SNPs in linkage disequilibrium (LD; r^2^>0.8) with three of the unavailable SNPs were identified using LDlink[17]. The 820 SNPs explained 7.43% of the variance in adult BMI (**eTable 3**).

The GIANT consortium had samples overlapping with the GWAS of type 2 diabetes (described below). Therefore, we selected a secondary set of IVs (734 independent SNPs) for adult BMI exclusively from the UK Biobank study (https://gwas.mrcieu.ac.uk/) when comparing the BMI-LADA association with the BMI-type 2 diabetes association. The 734 SNPs explained 7.86% of the variance in adult BMI.

### GWAS of LADA

LADA was the primary outcome in the present study. We obtained summary statistics for the association between the abovementioned SNPs and LADA from the only GWAS of LADA hitherto. It included 2,634 LADA cases and 5,947 population controls of European ancestry from Sweden, Denmark, Germany, and the UK, and the analysis was adjusted for sex and principal components (to correct for potential bias due to population structure)[1]. LADA was defined based on: (1) adult-onset (age at diagnosis >20, 30, or 35 years); (2) the presence of diabetes-associated autoimmune autoantibodies, in particular glutamic acid decarboxylase autoantibody (GADA) positivity; and (3) lack of insulin requirement for 6 months or 1 year after diagnosis[1].

### GWAS of type 2 diabetes

Summary statistics for the SNP-type 2 diabetes association were obtained from the DIAGRAM (DIAbetes Genetics Replication And Meta-analysis) Consortium, which included 26,676 type 2 diabetes cases and 132,532 controls of European ancestry[18]. Summary statistics in this study were adjusted for age, sex, and principal components[18].

### Data Harmonization

We checked the effect allele, reference allele, and effect allele frequency in the GWAS datasets. The β coefficient for the SNP-exposure association was “flipped” if the effect allele for the SNP-exposure association was the reference allele for the SNP-outcome association. We paid attention to palindromic (A/T or C/G) SNPs and there is no ambiguity in matching effect alleles for these SNPs between GWAS datasets of the exposure and the outcome.

### Statistical analysis

We measured instrument strength of each SNP using F statistic[19], which equals the β coefficient for the SNP-exposure association divided by the square of the standard error for the β coefficient. A larger F statistic indicates stronger instrument strength[19]. Birth weight and adult BMI were all inverse-normally transformed and analyzed in an additive model in the GWAS. Therefore, the risk estimates based directly on the summary statistics were ORs and 95% CIs for LADA and type 2 diabetes per SD change in exposures.

#### Main analysis

The inverse-variance weighted (IVW) method was used to assess the risk of LADA in relation to birth weight based on 129 IVs and adult BMI based on 820 primary IVs. The IVW method can be fitted in weighted linear regression[20, 21]. It provides a more precise risk estimate than other methods when all the IVs are valid[22].

#### Sensitivity analyses

Central assumptions in an MR study are that an IV only affects the outcome through the exposure, not through a direct pathway to the outcome or via a confounder (**eFigure 1**)[23], otherwise there will be directional pleiotropy and the IV is invalid.

**Figure 1.**
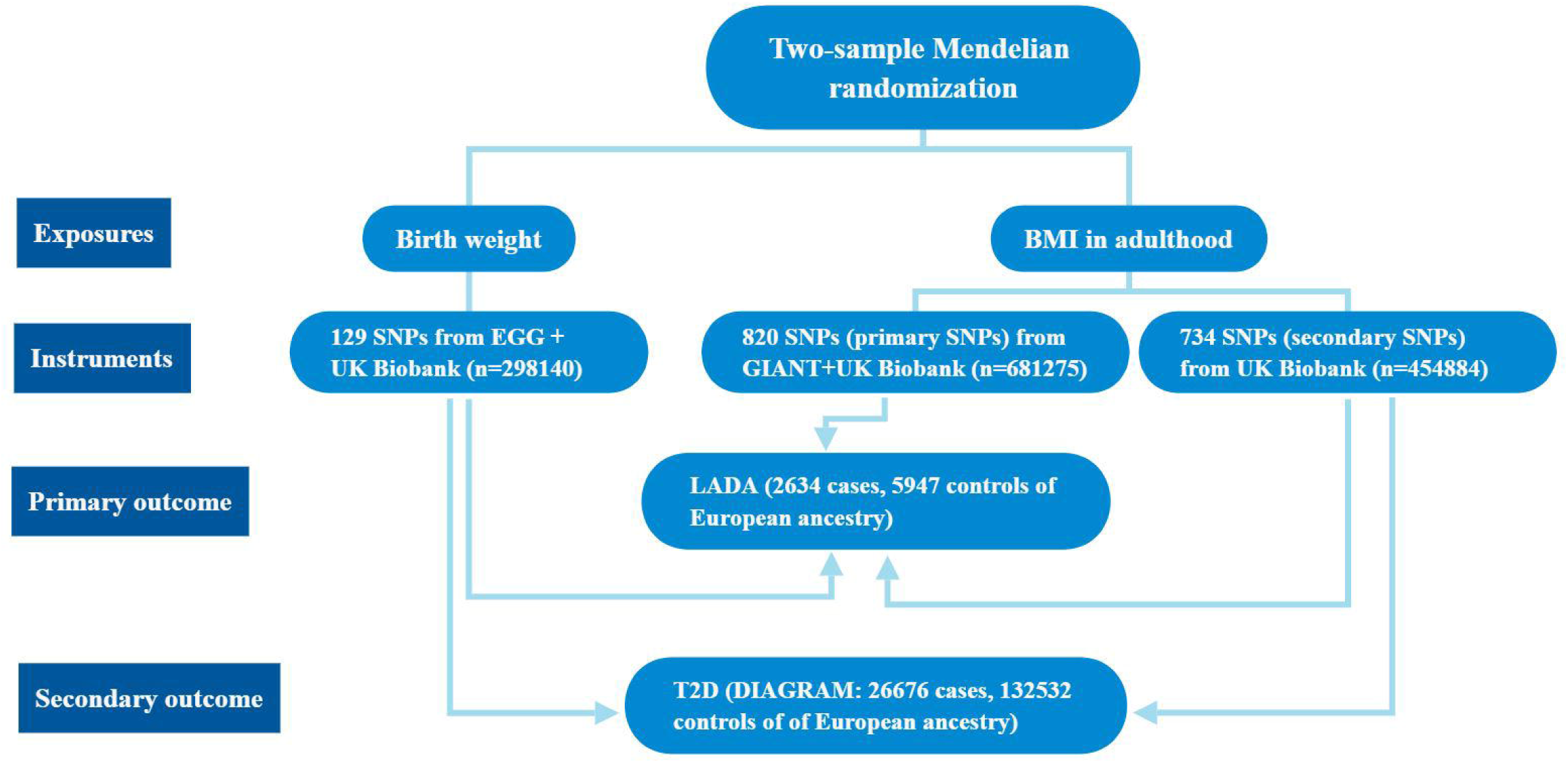
Flow chart of the study design in the present study. EGG: Early Growth Genetics Consortium; GIANT: Genetic Investigation of ANthropometric Traits consortium; LADA: latent autoimmune diabetes in adults; DIAGRAM: the DIAbetes Genetics Replication And Meta-analysis Consortium.

The IVW estimator used in the main analyses assumes that all the IVs are valid[22]. Several sensitivity analyses using other MR estimators were conducted to test the robustness of the results based on the IVW estimator. Some of them can detect potential directional pleiotropy, and some of them do not require all IVs to be valid. First we used the robust IVW method which replaces the standard linear regression in the IVW method with a robust regression[24]. This method has a greater power than IVW to reject causal null hypothesis when there is balanced pleiotropy (average pleiotropic effect: 0) and the instrument strength is independent of instrument’s direct effect (InSIDE[25] assumption) [22]. Furthermore, the weighted median method was used, which provides consistent estimates when >50% of IVs are valid and does not rely on the InSIDE assumption[26]. MR-Egger (Egger regression of Mendelian randomization) can give a causal estimate under the InSIDE[25] assumption even when all the IVs are invalid, with the slope coefficient of the regression model representing the logarithmic OR[25]. This method indicates overall directional pleiotropy[25] if the estimated intercept term in the regression model is non-zero[25]. Finally, the MR-PRESSO (the Mendelian randomization pleiotropy residual sum and outlier approach) estimator was used. MR-PRESSO is based on the IVW method and detects outliers (potential pleiotropic SNPs), produces corrected OR by removing outliers, and evaluates distortion of risk estimate by outliers [27].

In the present study, we further excluded outliers detected by MR-PRESSO from the IVs, and then re-analyzed the data using all different MR estimators including the IVW estimator.

We did other sensitivity analyses based on the IVW method by excluding some SNPs from the 129 IVs for birth weight and 820 primary IVs for adult BMI. These sensitivity analyses included several conservative analyses to minimize the possibility that the IVs affect the outcome through a pathway outside the exposure, and a leave-one-out analysis to investigate whether the association in the main analysis would disappear by excluding any one of the IVs. In the analysis of birth weight, conservative analysis 1 only included SNPs with fetal-only effects as IVs, to minimize the possibility that IVs affect LADA through maternal confounding factors; Conservative analysis 2 further excluded SNPs associated with diabetes-related traits at nominal significance level (Bonferroni-corrected), to minimize the possibility that IVs affect LADA directly; We further excluded SNPs associated with any trait (except birth weight) at genome-wide significance (conservative analysis 3), and SNPs associated with lifestyle factors (conservative analysis 4) and adult body size (conservative analysis 5) at nominal significance level (Bonferroni-corrected). In the analysis of adult BMI, we excluded SNPs associated with diabetes-related traits at nominal significance level (Bonferroni-corrected) or any trait (except adult body size) at genome-wide significance (conservative analysis 1), SNPs associated with lifestyle factors at nominal significance level (Bonferroni-corrected, conservative analysis 2), and SNPs associated with birth weight at nominal significance level (Bonferroni-corrected, conservative analysis 3). More details about conservative analyses are provided in **eFigure 2 and eTable 4**.

**Figure 2.**
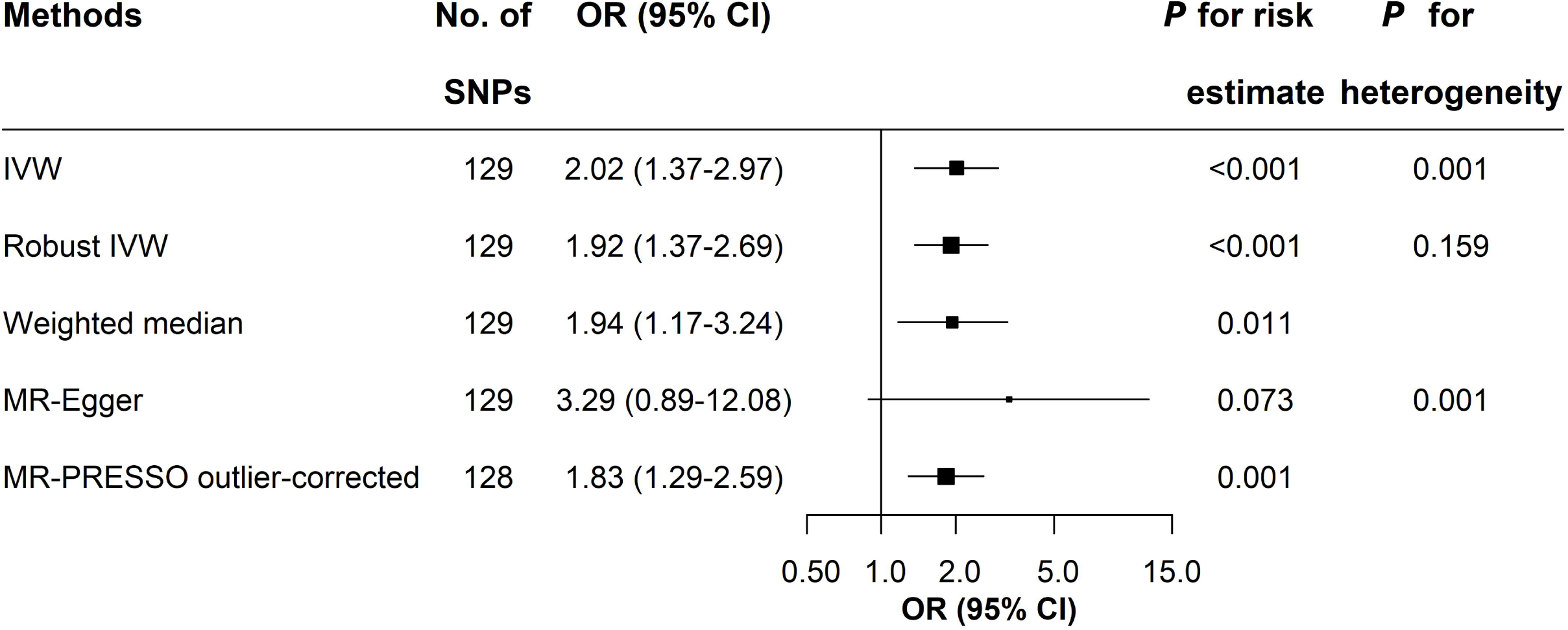
The risk of LADA in relation to one SD (0.5 kg) decrease in birth weight. IVW: inverse-variance weighted. MR-Egger: Egger regression of Mendelian randomization; MR-PRESSO: the Mendelian randomization pleiotropy residual sum and outlier approach; LADA: latent autoimmune diabetes in adults. MR-Egger intercept: -0.012, *P* for directional pleiotropy: 0.443. MR-PRESSO detected rs3184504 as the outlier and the outlier was excluded from the outlier corrected estimate (*P* for distortion of estimate: 0.457).

#### Comparison between LADA and type 2 diabetes

For comparison between LADA and type 2 diabetes, we used the same 129 IVs for birth weight and 734 secondary IVs for adult BMI in the analyses, to ensure the comparability of results. The associations of birth weight and adult BMI with type 2 diabetes were assessed using all the different MR estimators described above. Outlier-corrected ORs (95% CIs) from MR-PRESSO were used for comparison between LADA and T2D if MR-PRESSO detected outliers, otherwise the results of IVW were used for comparison.

MR analysis was conducted using *MendelianRandomization* and *MR-PRESSO* package in R 4.0.4. All statistical tests were two-sided, with *P*<0.05 indicating statistical significance.

### Ethical approval

Our study only used GWAS summary statistics, and no individual-level data were used. An ethical permit is not required according to Karolinska Institutet.

## Results

A flow chart of the study design is provided in **Figure 1**. The SD of birth weight and BMI in adulthood was approximately 500 g and 4.8 kg/m^2^, respectively (**eTable 1**). F statistics for IVs were all above 10 in the present study (**eTable 2** and **eTable 3**).

### Birth weight and LADA

Genetically determined birth weight based on 129 IVs was inversely associated with LADA (**eFigure 3**). In the main analysis, the OR of LADA was 2.02 (95% CI: 1.37-2.97) for each SD decrease in birth weight (**Figure 2**).

**Figure 3.**
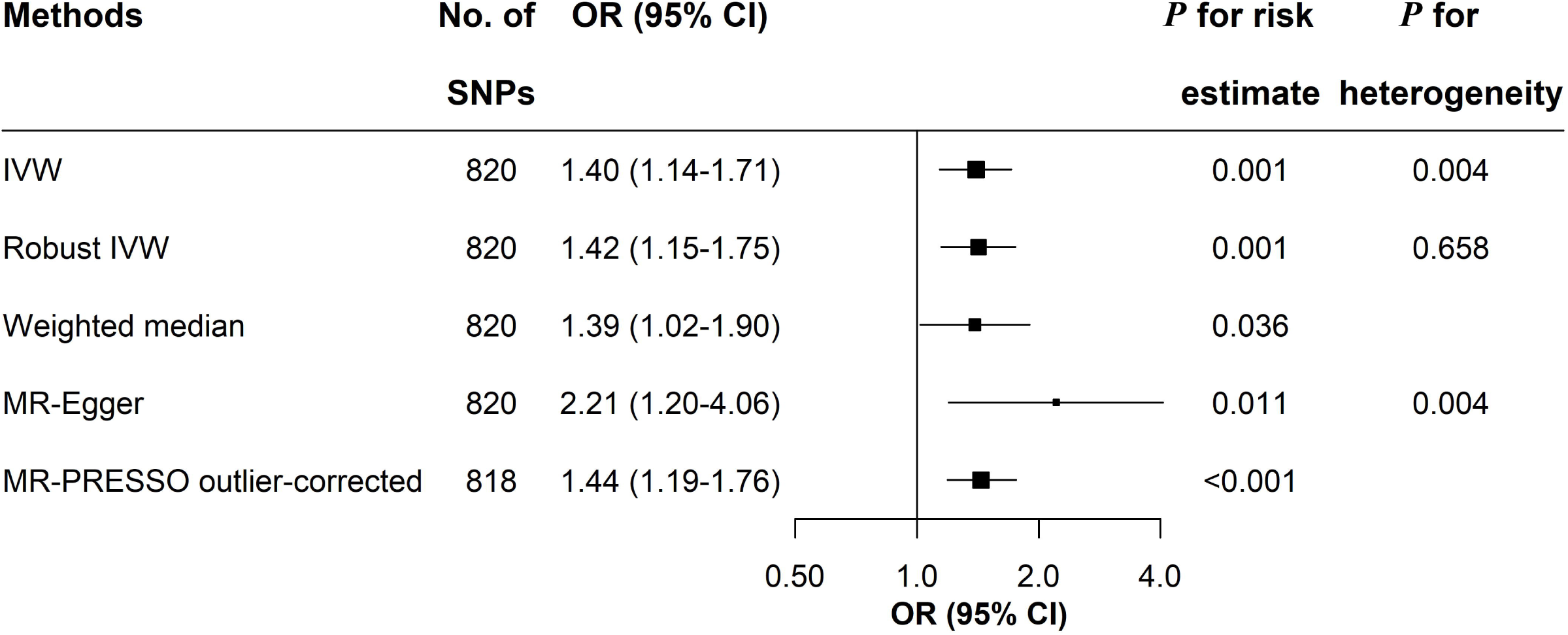
The risk of LADA in relation to one SD (4.8 kg/m^2^) increase in adult BMI. IVW: inverse-variance weighted. MR-Egger: Egger regression of Mendelian randomization; MR-PRESSO: the Mendelian randomization pleiotropy residual sum and outlier approach; LADA: latent autoimmune diabetes in adults. The MR-PRESSO identified two outliers: rs11066188 and rs10840606. Outliers were excluded from the outlier-corrected estimate (*P* for distortion of estimate: 0.777).

The inverse association between birth weight and LADA was also observed using other MR estimators (**Figure 2**). MR-Egger indicated no directional pleiotropy (intercept: -0.012, *P* for directional pleiotropy: 0.443). MR-PRESSO detected the SNP rs3184504 as the outlier and the outlier-corrected OR was 1.83 (95% CI: 1.29-2.59). No distortion in estimate by the outlier (*P*: 0.457) was detected in MR-PRESSO. After excluding rs3184504 from the IVs, results from different MR estimators also supported an inverse association between birth weight and LADA, and no directional pleiotropy was detected based on the 128 remaining IVs (**eFigure 4**).

The inverse association between birth weight and LADA was also observed in a series of conservative analyses by restricting IVs to SNPs with fetal-only effects, by further excluding SNPs associated with diabetes-related traits at *P*<0.05/129, by further excluding SNPs associated with any other trait at *P*<5×10^−8^, and by further excluding SNPs associated with lifestyle factors or adult BMI at *P*<0.05/129 (**eTable 5**). Leaving one of the 129 SNPs out each time did not change the direction of association (**eFigure 5**).

### Adult BMI and LADA

Genetically determined adult BMI based on the 820 primary IVs was positively associated with LADA (**eFigure 6**). One SD increase in adult BMI was associated with an OR of 1.40 (95% CI: 1.14-1.71) for LADA using the IVW method (**Figure 3**).

MR-Egger indicated no directional pleiotropy (intercept = -0.007, *P* for directional pleiotropy: 0.119) while MR-PRESSO detected two outliers, rs11066188 and rs10840606. Outliers did not distort the results (*P* for distortion test: 0.777) and the outlier-corrected OR was similar to the OR obtained with IVW. Robust IVW and weighted median also showed results comparable to those estimated by IVW (**Figure 3**). After excluding rs11066188 and rs10840606 from the IVs, no major change in results from different MR estimators was observed (**eFigure 7**).

All the three conservative analyses (**eTable 5**) and the leave-one-out analysis (data not shown) showed similar results with the main analysis.

### Comparison between LADA and type 2 diabetes

When assessing the association between birth weight and type 2 diabetes, MR-PRESSO detected 10 outliers and indicated distortion by outliers (*P* for distortion: 0.003, **eTable 6**). After correction for outliers, the OR of type 2 diabetes in relation to one SD decrease in genetically determined birth weight was 1.45 (95% CI: 1.25-1.70), which was slightly smaller than the magnitude of the birth weight-LADA association (**Table 1**).

MR-PRESSO detected 9 outliers when assessing the association between adult BMI and T2D based on the 734 secondary IVs (**eTable 7**). The OR of type 2 diabetes was 2.33 (95% CI: 2.16-2.51) for each SD increase in adult BMI after outlier removal (**Table 1** and **eTable 7**). For LADA, the same IVs yielded an outlier-corrected OR of 1.42 (95% CI: 1.18-1.72) (**Table 1, eTable 7**) which was similar to the association between LADA and adult BMI observed based on the 820 primary IVs.

## Discussion

### Main findings

This study provides genetic evidence that low birth weight and adult adiposity confer an increased risk of LADA. We also confirm findings of previous MR studies in showing that lower birth weight[28-30] and higher BMI[31-38] in adulthood are associated with increased risk of type 2 diabetes. To the best of our knowledge, this is the first MR study to explore the role of environmental/lifestyle factors in the etiology of LADA.

### Main findings in relation to previous studies

The results regarding birth weight are in line with those of our previous observational study indicating a two-fold increased risk of LADA in individuals with a birth weight <3 kg compared to ≥4kg[9]. Notably, the association with birth weight was not weaker for LADA than for type 2 diabetes which is also in line with previous observational data[9]. The mechanism linking birth weight to LADA and type 2 diabetes remains unclear. The Barker hypothesis proposes that an adverse intrauterine environment leads to both lower birth weight and higher risk of future cardiometabolic risk[39]. However, we restricted IVs to SNPs with fetal-only effects and there was no major change in OR. Therefore, the inverse association observed in the present study may not be explained by pleiotropy introduced by the intrauterine environment. The fetal insulin hypothesis proposes that genetically determined insulin resistance in the fetus results in impaired insulin-mediated fetal growth as well as insulin resistance in adult life[40]. However, our findings do not support this hypothesis, since the association between birth weight and LADA did not attenuate after excluding SNPs associated with diabetes-related traits (including insulin resistance). Our findings suggest a direct effect of low birth weight on the risk of LADA, but the underlying mechanism remains to be explored.

Findings regarding LADA and adult BMI were also in line with previous observational data[8]. Overweight and obesity are strongly associated with development of insulin resistance[41] and this may explain a causal link between adult adiposity and LADA. In support hereof, a positive association between BMI and insulin resistance was observed in LADA patients[8]. The association with BMI was stronger for type 2 diabetes than LADA, which is in line with previous findings[8]. This is to be expected since insulin resistance tends to be less pronounced in LADA compared to type 2 diabetes patients [2, 8, 42]. A previous MR study also found support for a link between childhood adiposity and type 1 diabetes[43]. This implies that overweight/obesity is implicated in the promotion of all major types of diabetes and emphasizes that it is crucial to prevent overweight in order to reduce the incidence of diabetes.

### Assessment of MR assumptions

A major concern in MR studies is the violation of IV assumptions. These assumptions cannot be fully tested, but we used several approaches to minimize this potential bias. First, we applied different MR estimators, some of which detected and corrected for potential directional pleiotropy from statistical perspective. There was no major change in ORs after excluding outliers, indicating the robustness of the results. Second, we excluded some SNPs in several conservative analyses, in which the associations of birth weight and adult BMI with LADA persisted. It should be noted that excluding these SNPs in the conservative analyses because of suggested pleiotropic effects in observational studies, does not prove that they are in fact invalid SNPs.

### Strengths and limitations

There are several strengths in the present study. First, the two-sample MR design using genetic variants as unbiased proxy minimizes confounding and reverse causation. Second, the application of different MR estimators and a series of conservative analyses reduces the risk of bias caused by directional pleiotropy. Third, we confirmed findings from previous studies on type 2 diabetes and used type 2 diabetes as a “positive control” to show that the instruments and methods used for LADA in the present study are reliable. This also provides a good opportunity to compare the etiology of LADA and type 2 diabetes. There are also some limitations. First, lacking individual data rules out the possibility of exploring potential nonlinear association between exposures and outcomes, which is a common limitation in MR studies based on summary statistics. The linear assumption is less likely to be violated in our MR study. For LADA, there is no evidence with sufficient statistical power to support a nonlinear association[8, 9]. Adult BMI seemed to be linearly (positively) associated with type 2 diabetes[33, 44]. Observational studies found that the linear (inverse) association between birth weight and type 2 diabetes held when birth weight<4.0 kg[45] or <4.5 kg[46]. The range of birth weight (study-specific: 2.5-4.5 kg or within the range of mean±5SD) in the GWAS[14] used by the present MR analysis is generally in the range of linear association. Moreover, deviation from the linear assumption is likely to reduce the statistical power in risk estimate, rather than generating spurious associations[47]. Second, findings from the present study were only applicable to the European population since the only GWAS on LADA was conducted in individuals of European ancestry. It is unclear to what extent the findings are generalizable to other populations.

In conclusion, these findings provide genetic support for a causal link between low birth weight, adult overweight/obesity and LADA. The results persisted in a series of sensitivity analyses. Measures should be taken to reduce the prevalence of adult overweight/obesity for the prevention of diabetes with and without an autoimmune component. The mechanism linking low birth weight to diabetes remains to be explored.

## Data Availability

This study only used summary data and these data are publicly available.

## Abbreviations

DIAGRAM: the DIAbetes Genetics Replication And Meta-analysis Consortium
EGG: the Early Growth Genetics Consortium
GIANT: the Genetic Investigation of ANthropometric Traits consortium
GWAS: genome-wide association study
InSIDE: the instrument strength is independent of instrument’s direct effect
IV: instrumental variable
IVW: inverse-variance weighted
LADA: latent autoimmune diabetes in adults
LADY: Latent autoimmune diabetes in the young
MR: Mendelian randomization
MR-Egger: Egger regression of Mendelian randomization
MR-PRESSO: the Mendelian randomization pleiotropy residual sum and outlier approach

## Acknowledgements

Not applicable.

## Authors’ contributions

YW and SC conceived and designed the study. YW collected the summary data, analyzed data, and drafted the first draft of the manuscript. YZ contributed to methodological issues. All authors critically revised the manuscript for valuable intellectual content. YW and SC contributed to the interpretation of the results. All authors reviewed and approved the final manuscript.

## Data availability

This study only used summary data and these data are publicly available.

