## Supplementary Material for "Birth weight, BMI in adulthood and latent autoimmune diabetes in adults: A Mendelian randomization study"

eTable 1. Basic characteristics of included GWAS studies for exposures

| **Year,**  **author** | **Studies** | **Sample**  **size** | **Ancestry** | **Exposure** | **Exposure measurement** | **SD of exposure** | **Exposure transformation** | **Quality control of SNPs** | | | | **Adjustment** |
| --- | --- | --- | --- | --- | --- | --- | --- | --- | --- | --- | --- | --- |
|  |  |  |  |  |  |  |  | **Call rate** | **INFO score** | **MAF** | **HWE *P*** |  |
| 2019,  Warrington NM ^[1]^ | EGG + UK Biobank | 298,140 | European | Birth weight | measurements at birth, obstetric records, medical registers, interviews with the mother, or self-report as adults | ~500 g | z-score transformed in men and women separately | study-specific: from >0.9 to >0.98 | unknown | study-specific: from >0.1% to >2% | study-specific: from >5e-8 to >1e-4 | study-specifc covariates such as gestational age and PCs |
| 2018,  Yengo L^[2]^ | UK Biobank +  GIANT | 681,275 (456,426 from UK Biobank) | European | adult BMI | mostly through measurement, some through self-report | ~4.8 kg/m^2^ | inverse-normally transformed | GIANT: study-specific UK Biobank:  >0.95 | GIANT: study-specific; UK Biobank:  >0.3 | GIANT: study-specific; UK Biobank:  >0.01% | GIANT: study-specific; UK Biobank:  > 1e-6 | UK Biobank: age, sex, recruitment centre, genotyping batches and 10 PCs. GIANT: age, age squared, and any necessary study-specific covariates |
| 2018,  Elsworth B | UK Biobank ^a^ | 454,884 | European | adult BMI | measured during the initial Assessment Centre visit | ~4.8 kg/m^2^ | inverse rank-normal transformed | unknown | >0.8 | >0.1% | > 1e-10 | age, sex, and PCs |

GWAS: genome-wide association study; MAF: major allele frequency; HWE: Hardy–Weinberg test; EGG: Early Growth Genetics Consortium; GIANT: Genetic Investigation of ANthropometric Traits consortium; PC: principal component.

^a^ Data were obtained from https://gwas.mrcieu.ac.uk/datasets/ukb-b-2303/.

### eTable 2. Detailed information on 129 instrumental variables for the association between birth weight and LADA

| **SNP** | **Effect allele** | **Other allele** | **SNP-birth weight association** | | | | |  | **SNP-LADA association** | | |
| --- | --- | --- | --- | --- | --- | --- | --- | --- | --- | --- | --- |
|  |  |  | **BETA** | **SE** | ***P*** | **R^2^** | **F  statistics** |  | **BETA** | **SE** | ***P*** |
| rs10883846 | C | T | 0.0168 | 0.0026 | 1.3E-10 | 1.3E-04 | 41.4 |  | 0.0323 | 0.0388 | 0.409 |
| rs7076938 | T | C | 0.0321 | 0.0029 | 2.1E-28 | 4.0E-04 | 122.3 |  | -0.0278 | 0.0421 | 0.511 |
| rs71486610 | C | G | 0.0203 | 0.0026 | 3.2E-15 | 2.1E-04 | 62.2 |  | 0.0519 | 0.0370 | 0.163 |
| rs4350272 | A | G | 0.0170 | 0.0029 | 3.6E-09 | 1.1E-04 | 34.9 |  | -0.0381 | 0.0420 | 0.367 |
| rs9645500 | G | T | 0.0243 | 0.0028 | 1.8E-18 | 2.5E-04 | 77.1 |  | -0.0828 | 0.0414 | 0.047 |
| rs1112718 | G | A | 0.0257 | 0.0026 | 3.8E-23 | 3.2E-04 | 98.3 |  | -0.0608 | 0.0380 | 0.112 |
| rs2274224 | C | G | 0.0214 | 0.0026 | 9.8E-17 | 2.2E-04 | 69.1 |  | 0.0067 | 0.0378 | 0.860 |
| rs4444073 | A | C | 0.0202 | 0.0026 | 2.7E-15 | 2.0E-04 | 62.6 |  | 0.0776 | 0.0374 | 0.039 |
| rs11042596 | T | G | 0.0269 | 0.0028 | 4.3E-22 | 3.2E-04 | 93.5 |  | -0.0140 | 0.0397 | 0.725 |
| rs234864 | A | G | 0.0157 | 0.0026 | 1.7E-09 | 1.2E-04 | 36.4 |  | -0.0568 | 0.0377 | 0.134 |
| rs5030317 | C | G | 0.0173 | 0.0029 | 2.7E-09 | 1.2E-04 | 35.4 |  | 0.0178 | 0.0426 | 0.678 |
| rs667515 | G | C | 0.0185 | 0.0027 | 9.3E-12 | 1.6E-04 | 46.6 |  | 0.0436 | 0.0388 | 0.264 |
| rs61885091 | A | G | 0.0229 | 0.0037 | 4.8E-10 | 1.5E-04 | 38.8 |  | -0.0213 | 0.0558 | 0.704 |
| rs10830963 | G | C | 0.0191 | 0.0029 | 2.8E-11 | 1.5E-04 | 44.4 |  | -0.0346 | 0.0416 | 0.408 |
| rs2647873 | A | G | 0.0181 | 0.0026 | 2.9E-12 | 1.6E-04 | 48.8 |  | -0.0531 | 0.0371 | 0.155 |
| rs3184504 | C | T | 0.0230 | 0.0026 | 2.6E-19 | 2.6E-04 | 80.9 |  | -0.2449 | 0.0375 | 0.000 |
| rs11055030 | G | C | 0.0200 | 0.0029 | 3.9E-12 | 1.6E-04 | 48.3 |  | 0.0551 | 0.0426 | 0.198 |
| rs2306547 | C | T | 0.0187 | 0.0026 | 4.4E-13 | 1.7E-04 | 52.6 |  | -0.0037 | 0.0373 | 0.921 |
| rs6582623 | C | T | 0.0236 | 0.0039 | 1.1E-09 | 1.3E-04 | 37.3 |  | -0.1080 | 0.0547 | 0.050 |
| rs7968682 | G | T | 0.0418 | 0.0026 | 4.2E-60 | 8.7E-04 | 267.6 |  | -0.0092 | 0.0374 | 0.806 |
| rs1480470 | G | A | 0.0243 | 0.0027 | 1.4E-19 | 2.8E-04 | 82.1 |  | 0.0598 | 0.0392 | 0.129 |
| rs9549046 | A | G | 0.0291 | 0.0041 | 8.0E-13 | 1.8E-04 | 51.4 |  | 0.0361 | 0.0574 | 0.532 |
| rs34217484 | A | T | 0.0192 | 0.0029 | 6.8E-11 | 1.4E-04 | 42.6 |  | -0.0245 | 0.0416 | 0.558 |
| rs9318511 | C | A | 0.0269 | 0.0039 | 6.0E-12 | 1.6E-04 | 47.4 |  | 0.1364 | 0.0590 | 0.022 |
| rs6575803 | C | T | 0.0316 | 0.0045 | 1.3E-12 | 1.9E-04 | 50.4 |  | 0.0121 | 0.0636 | 0.850 |
| rs75844534 | A | C | 0.0259 | 0.0039 | 4.9E-11 | 1.5E-04 | 43.3 |  | -0.1523 | 0.0605 | 0.012 |
| rs339969 | A | C | 0.0168 | 0.0027 | 2.2E-10 | 1.3E-04 | 40.4 |  | -0.0026 | 0.0383 | 0.946 |
| rs4932373 | A | C | 0.0201 | 0.0028 | 3.0E-13 | 1.8E-04 | 53.3 |  | -0.0212 | 0.0397 | 0.596 |
| rs55958435 | A | G | 0.0247 | 0.0030 | 1.6E-16 | 2.3E-04 | 68.2 |  | 0.0647 | 0.0424 | 0.129 |
| rs7402983 | A | C | 0.0241 | 0.0027 | 2.6E-19 | 2.8E-04 | 80.8 |  | -0.0611 | 0.0382 | 0.112 |
| rs11630479 | G | A | 0.0139 | 0.0028 | 8.9E-07 | 8.0E-05 | 24.2 |  | -0.0262 | 0.0418 | 0.533 |
| rs2045457 | G | A | 0.0162 | 0.0028 | 6.3E-09 | 1.1E-04 | 33.8 |  | -0.0480 | 0.0400 | 0.232 |
| rs40434 | G | A | 0.0167 | 0.0027 | 3.0E-10 | 1.3E-04 | 39.7 |  | -0.0224 | 0.0373 | 0.551 |
| rs28544888 | C | T | 0.0258 | 0.0046 | 1.6E-08 | 1.1E-04 | 31.9 |  | 0.0727 | 0.0671 | 0.281 |
| rs9909342 | A | G | 0.0179 | 0.0027 | 2.2E-11 | 1.5E-04 | 44.9 |  | -0.0297 | 0.0384 | 0.442 |
| rs7223535 | G | A | 0.0214 | 0.0029 | 2.1E-13 | 1.8E-04 | 54.0 |  | -0.0316 | 0.0427 | 0.462 |
| rs11867479 | T | C | 0.0172 | 0.0027 | 1.1E-10 | 1.4E-04 | 41.7 |  | 0.0133 | 0.0389 | 0.733 |
| rs10221267 | T | C | 0.0167 | 0.0026 | 6.5E-11 | 1.4E-04 | 42.7 |  | -0.0177 | 0.0372 | 0.635 |
| rs222857 | T | C | 0.0265 | 0.0026 | 1.1E-24 | 3.4E-04 | 105.4 |  | -0.0429 | 0.0379 | 0.261 |
| rs4511593 | T | C | 0.0175 | 0.0027 | 1.1E-10 | 1.4E-04 | 41.7 |  | 0.0099 | 0.0392 | 0.801 |
| rs11082304 | T | G | 0.0160 | 0.0026 | 4.2E-10 | 1.3E-04 | 39.1 |  | -0.0489 | 0.0375 | 0.194 |
| rs1129156 | T | C | 0.0174 | 0.0029 | 2.5E-09 | 1.2E-04 | 35.6 |  | 0.0336 | 0.0413 | 0.418 |
| rs147957154 | T | C | 0.0232 | 0.0039 | 2.8E-09 | 1.2E-04 | 35.4 |  | 0.0466 | 0.0551 | 0.400 |
| rs2779165 | G | C | 0.0221 | 0.0034 | 7.6E-11 | 1.5E-04 | 42.4 |  | -0.1001 | 0.0477 | 0.037 |
| rs516246 | C | T | 0.0175 | 0.0026 | 9.3E-12 | 1.5E-04 | 46.5 |  | -0.0276 | 0.0369 | 0.457 |
| rs255773 | C | T | 0.0182 | 0.0027 | 1.3E-11 | 1.6E-04 | 45.8 |  | 0.0508 | 0.0380 | 0.184 |
| rs8106042 | G | C | 0.0204 | 0.0029 | 2.2E-12 | 1.7E-04 | 49.4 |  | -0.0702 | 0.0422 | 0.098 |
| rs80278614 | A | G | 0.0404 | 0.0059 | 6.5E-12 | 1.7E-04 | 47.3 |  | 0.0188 | 0.0773 | 0.809 |
| rs905938 | C | T | 0.0261 | 0.0029 | 2.8E-19 | 2.6E-04 | 80.7 |  | -0.0132 | 0.0414 | 0.752 |
| rs670523 | G | A | 0.0188 | 0.0027 | 7.6E-12 | 1.6E-04 | 46.9 |  | -0.0158 | 0.0395 | 0.690 |
| rs72480273 | C | A | 0.0225 | 0.0034 | 4.0E-11 | 1.5E-04 | 43.7 |  | -0.1061 | 0.0501 | 0.035 |
| rs61830764 | A | G | 0.0166 | 0.0027 | 1.1E-09 | 1.3E-04 | 37.2 |  | 0.0147 | 0.0383 | 0.703 |
| rs3806315 | A | G | 0.0177 | 0.0027 | 2.8E-11 | 1.5E-04 | 44.4 |  | -0.0027 | 0.0381 | 0.943 |
| rs708122 | C | A | 0.0165 | 0.0028 | 2.5E-09 | 1.2E-04 | 35.6 |  | 0.0350 | 0.0399 | 0.384 |
| rs12401656 | G | A | 0.0252 | 0.0038 | 3.4E-11 | 1.5E-04 | 44.0 |  | -0.0251 | 0.0576 | 0.665 |
| rs6040076 | C | G | 0.0190 | 0.0026 | 4.4E-13 | 1.8E-04 | 52.6 |  | -0.0156 | 0.0373 | 0.678 |
| rs6033062 | A | T | 0.0162 | 0.0026 | 5.2E-10 | 1.3E-04 | 38.7 |  | -0.0249 | 0.0374 | 0.508 |
| rs11698914 | C | G | 0.0319 | 0.0031 | 1.2E-24 | 3.6E-04 | 105.2 |  | -0.0334 | 0.0435 | 0.445 |
| rs2889874 | G | T | 0.0161 | 0.0026 | 9.4E-10 | 1.3E-04 | 37.5 |  | -0.0098 | 0.0382 | 0.798 |
| rs1012167 | C | T | 0.0241 | 0.0027 | 1.2E-19 | 2.8E-04 | 82.3 |  | -0.0694 | 0.0386 | 0.074 |
| rs753381 | T | C | 0.0151 | 0.0026 | 3.4E-09 | 1.1E-04 | 35.0 |  | 0.0188 | 0.0376 | 0.619 |
| rs6026449 | C | T | 0.0170 | 0.0027 | 2.5E-10 | 1.3E-04 | 40.1 |  | -0.0170 | 0.0386 | 0.662 |
| rs73143584 | A | G | 0.0288 | 0.0043 | 1.8E-11 | 1.6E-04 | 45.2 |  | 0.0228 | 0.0607 | 0.709 |
| rs2229742 | G | C | 0.0272 | 0.0042 | 7.4E-11 | 1.5E-04 | 42.5 |  | 0.0107 | 0.0587 | 0.856 |
| rs220193 | A | G | 0.0206 | 0.0031 | 4.1E-11 | 1.5E-04 | 43.6 |  | -0.0424 | 0.0442 | 0.341 |
| rs134594 | C | T | 0.0168 | 0.0027 | 5.8E-10 | 1.3E-04 | 38.5 |  | -0.0321 | 0.0394 | 0.418 |
| rs41311445 | A | C | 0.0326 | 0.0045 | 3.3E-13 | 1.9E-04 | 53.1 |  | -0.1106 | 0.0609 | 0.071 |
| rs7285579 | C | T | 0.0173 | 0.0029 | 2.7E-09 | 1.3E-04 | 35.5 |  | 0.0305 | 0.0418 | 0.468 |
| rs2280235 | G | A | 0.0183 | 0.0030 | 6.9E-10 | 1.3E-04 | 38.1 |  | 0.0260 | 0.0436 | 0.553 |
| rs10181515 | T | C | 0.0214 | 0.0030 | 2.1E-12 | 1.6E-04 | 49.4 |  | -0.0762 | 0.0445 | 0.088 |
| rs2551347 | T | C | 0.0245 | 0.0030 | 1.9E-16 | 2.2E-04 | 67.8 |  | -0.0097 | 0.0433 | 0.823 |
| rs754868 | G | A | 0.0159 | 0.0026 | 6.7E-10 | 1.2E-04 | 38.2 |  | -0.0490 | 0.0381 | 0.201 |
| rs17034876 | T | C | 0.0422 | 0.0029 | 3.1E-47 | 7.5E-04 | 208.6 |  | -0.0676 | 0.0417 | 0.107 |
| rs4953353 | G | T | 0.0179 | 0.0027 | 3.5E-11 | 1.5E-04 | 44.0 |  | -0.0095 | 0.0389 | 0.809 |
| rs10495563 | A | G | 0.0221 | 0.0027 | 2.1E-16 | 2.2E-04 | 67.6 |  | -0.0731 | 0.0394 | 0.065 |
| rs11708067 | G | A | 0.0409 | 0.0030 | 1.6E-42 | 6.1E-04 | 186.9 |  | -0.0484 | 0.0440 | 0.274 |
| rs2306700 | T | C | 0.0228 | 0.0038 | 1.8E-09 | 1.2E-04 | 36.2 |  | 0.0003 | 0.0534 | 0.995 |
| rs10935733 | T | C | 0.0194 | 0.0026 | 2.3E-13 | 1.8E-04 | 53.8 |  | 0.0152 | 0.0382 | 0.693 |
| rs1482852 | A | G | 0.0504 | 0.0026 | 1.6E-82 | 1.2E-03 | 370.5 |  | -0.0312 | 0.0376 | 0.410 |
| rs11711420 | T | G | 0.0187 | 0.0030 | 3.2E-10 | 1.3E-04 | 39.6 |  | 0.0006 | 0.0438 | 0.989 |
| rs2168443 | T | A | 0.0166 | 0.0027 | 3.9E-10 | 1.3E-04 | 39.2 |  | -0.0009 | 0.0387 | 0.982 |
| rs6533183 | C | T | 0.0218 | 0.0027 | 6.8E-16 | 2.2E-04 | 65.3 |  | -0.0425 | 0.0386 | 0.274 |
| rs6845999 | T | C | 0.0263 | 0.0026 | 1.5E-24 | 3.4E-04 | 104.8 |  | -0.0445 | 0.0373 | 0.236 |
| rs4144829 | C | T | 0.0355 | 0.0029 | 4.3E-34 | 4.9E-04 | 148.4 |  | -0.0238 | 0.0420 | 0.572 |
| rs1981627 | G | A | 0.0171 | 0.0026 | 8.4E-11 | 1.4E-04 | 42.2 |  | 0.0379 | 0.0379 | 0.320 |
| rs2946179 | C | T | 0.0198 | 0.0029 | 1.1E-11 | 1.5E-04 | 46.1 |  | 0.1036 | 0.0433 | 0.017 |
| rs351930 | T | A | 0.0192 | 0.0032 | 2.9E-09 | 1.2E-04 | 35.3 |  | -0.0127 | 0.0472 | 0.790 |
| rs854037 | A | G | 0.0265 | 0.0033 | 9.4E-16 | 2.1E-04 | 64.6 |  | -0.0145 | 0.0480 | 0.763 |
| rs28365970 | C | A | 0.0199 | 0.0029 | 1.7E-11 | 1.5E-04 | 45.4 |  | -0.0336 | 0.0427 | 0.434 |
| rs76094073 | G | C | 0.0265 | 0.0039 | 1.6E-11 | 1.5E-04 | 45.4 |  | 0.0288 | 0.0563 | 0.611 |
| rs6925689 | T | C | 0.0150 | 0.0026 | 6.4E-09 | 1.1E-04 | 33.8 |  | -0.0300 | 0.0370 | 0.421 |
| rs6569647 | T | C | 0.0200 | 0.0032 | 6.3E-10 | 1.3E-04 | 38.3 |  | -0.0004 | 0.0470 | 0.993 |
| rs6930558 | T | G | 0.0218 | 0.0030 | 3.4E-13 | 1.8E-04 | 53.0 |  | 0.0160 | 0.0428 | 0.711 |
| rs962554 | T | C | 0.0168 | 0.0029 | 3.8E-09 | 1.2E-04 | 34.8 |  | -0.0028 | 0.0413 | 0.947 |
| rs10872678 | T | C | 0.0317 | 0.0028 | 9.8E-29 | 4.0E-04 | 123.9 |  | -0.0717 | 0.0412 | 0.084 |
| rs2934844 | T | A | 0.0208 | 0.0028 | 1.8E-13 | 1.9E-04 | 54.3 |  | 0.0490 | 0.0406 | 0.230 |
| rs35261542 | C | A | 0.0406 | 0.0029 | 2.8E-45 | 6.5E-04 | 199.6 |  | -0.0705 | 0.0410 | 0.087 |
| rs9379832 | A | G | 0.0220 | 0.0030 | 1.1E-13 | 1.9E-04 | 55.3 |  | 0.0636 | 0.0423 | 0.135 |
| rs9366778 | G | A | 0.0180 | 0.0027 | 2.9E-11 | 1.5E-04 | 44.3 |  | -0.0921 | 0.0393 | 0.020 |
| rs9267812 | T | C | 0.0230 | 0.0039 | 3.1E-09 | 1.2E-04 | 35.2 |  | -0.0329 | 0.0563 | 0.561 |
| rs1547669 | G | A | 0.0178 | 0.0026 | 6.2E-12 | 1.6E-04 | 47.3 |  | 0.0049 | 0.0373 | 0.896 |
| rs75104038 | A | G | 0.0449 | 0.0055 | 4.3E-16 | 2.3E-04 | 66.2 |  | -0.0226 | 0.0817 | 0.784 |
| rs9348981 | T | G | 0.0210 | 0.0029 | 2.2E-13 | 1.8E-04 | 53.9 |  | -0.0484 | 0.0411 | 0.242 |
| rs7744700 | T | A | 0.0199 | 0.0029 | 1.6E-11 | 1.6E-04 | 45.4 |  | 0.0795 | 0.0420 | 0.060 |
| rs6467157 | T | C | 0.0195 | 0.0029 | 1.5E-11 | 1.6E-04 | 45.7 |  | 0.0184 | 0.0415 | 0.659 |
| rs59084784 | A | C | 0.0166 | 0.0028 | 2.4E-09 | 1.2E-04 | 35.7 |  | -0.0539 | 0.0403 | 0.184 |
| rs34776209 | C | T | 0.0232 | 0.0030 | 8.5E-15 | 2.0E-04 | 60.3 |  | -0.0509 | 0.0440 | 0.250 |
| rs4719648 | C | T | 0.0191 | 0.0026 | 2.6E-13 | 1.8E-04 | 53.6 |  | 0.0112 | 0.0375 | 0.767 |
| rs11983722 | A | T | 0.0319 | 0.0054 | 3.1E-09 | 1.2E-04 | 35.2 |  | 0.0051 | 0.0781 | 0.949 |
| rs10265057 | G | A | 0.0273 | 0.0045 | 1.3E-09 | 1.2E-04 | 36.9 |  | 0.0838 | 0.0653 | 0.202 |
| rs2237467 | A | G | 0.0182 | 0.0031 | 5.3E-09 | 1.1E-04 | 34.1 |  | -0.0609 | 0.0435 | 0.163 |
| rs112139215 | A | C | 0.0475 | 0.0051 | 2.8E-20 | 2.8E-04 | 85.2 |  | -0.0653 | 0.0725 | 0.371 |
| rs2282978 | C | T | 0.0183 | 0.0027 | 1.7E-11 | 1.5E-04 | 45.4 |  | 0.0059 | 0.0395 | 0.882 |
| rs7819593 | C | T | 0.0218 | 0.0030 | 6.2E-13 | 1.7E-04 | 51.8 |  | -0.0099 | 0.0424 | 0.816 |
| rs13271368 | C | T | 0.0202 | 0.0030 | 2.3E-11 | 1.5E-04 | 44.8 |  | -0.0847 | 0.0429 | 0.050 |
| rs13257363 | G | A | 0.0176 | 0.0026 | 2.0E-11 | 1.5E-04 | 45.0 |  | 0.0797 | 0.0379 | 0.036 |
| rs9657468 | G | T | 0.0149 | 0.0028 | 7.9E-08 | 9.9E-05 | 28.9 |  | -0.0165 | 0.0395 | 0.679 |
| rs732563 | C | T | 0.0174 | 0.0026 | 1.3E-11 | 1.5E-04 | 45.9 |  | -0.0437 | 0.0372 | 0.242 |
| rs34036147 | T | C | 0.0185 | 0.0028 | 8.4E-11 | 1.5E-04 | 42.2 |  | -0.0555 | 0.0397 | 0.165 |
| rs13266210 | A | G | 0.0268 | 0.0031 | 1.5E-17 | 2.4E-04 | 72.8 |  | 0.0664 | 0.0467 | 0.157 |
| rs72656010 | T | C | 0.0283 | 0.0038 | 1.4E-13 | 1.8E-04 | 54.8 |  | -0.1531 | 0.0547 | 0.005 |
| rs62496903 | T | C | 0.0328 | 0.0048 | 6.7E-12 | 1.6E-04 | 47.2 |  | 0.0014 | 0.0696 | 0.984 |
| rs2418135 | A | G | 0.0199 | 0.0026 | 1.5E-14 | 2.0E-04 | 59.1 |  | 0.0600 | 0.0376 | 0.112 |
| rs1323438 | C | T | 0.0190 | 0.0029 | 5.6E-11 | 1.5E-04 | 43.0 |  | 0.0123 | 0.0408 | 0.763 |
| rs3933326 | G | A | 0.0213 | 0.0028 | 2.3E-14 | 2.0E-04 | 58.3 |  | -0.0194 | 0.0390 | 0.620 |
| rs10985827 | G | T | 0.0300 | 0.0037 | 6.1E-16 | 2.2E-04 | 65.5 |  | -0.0016 | 0.0558 | 0.978 |
| rs28505901 | A | G | 0.0244 | 0.0031 | 2.5E-15 | 2.2E-04 | 62.8 |  | -0.0237 | 0.0429 | 0.583 |
| rs7854962 | C | G | 0.0217 | 0.0032 | 1.0E-11 | 1.6E-04 | 46.4 |  | 0.0273 | 0.0470 | 0.564 |
| rs28457693 | G | A | 0.0442 | 0.0042 | 9.9E-26 | 3.8E-04 | 110.1 |  | 0.0002 | 0.0577 | 0.997 |

LADA: latent autoimmune diabetes in adults.

### eTable 3. Detailed information on 820 instrumental variables for the association between BMI in adulthood and LADA

| **SNP** | **Effect allele** | **Other allele** | **SNP-BMI association** | | | | |  | **SNP-LADA association** | | |
| --- | --- | --- | --- | --- | --- | --- | --- | --- | --- | --- | --- |
|  |  |  | **BETA** | **SE** | ***P*** | **R^2^** | **F**  **statistics** |  | **BETA** | **SE** | ***P*** |
| rs1227244 | G | A | 0.0106 | 0.0019 | 1.00E-08 | 5.02E-05 | 31.1 |  | -0.0117 | 0.0393 | 7.66E-01 |
| rs11119208 | A | G | 0.0095 | 0.0017 | 1.00E-08 | 4.28E-05 | 31.2 |  | -0.0239 | 0.0385 | 5.38E-01 |
| rs719802 | T | C | 0.0101 | 0.0018 | 9.50E-09 | 4.81E-05 | 31.5 |  | -0.0063 | 0.0378 | 8.69E-01 |
| rs1169091 | C | T | 0.0113 | 0.002 | 6.90E-09 | 5.11E-05 | 31.9 |  | -0.0239 | 0.0417 | 5.68E-01 |
| rs1277723 | A | G | 0.0113 | 0.002 | 1.00E-08 | 4.44E-05 | 31.9 |  | 0.0029 | 0.0445 | 9.47E-01 |
| rs2119753 | A | G | 0.0102 | 0.0018 | 6.80E-09 | 4.95E-05 | 32.1 |  | -0.0248 | 0.0382 | 5.19E-01 |
| rs7801551 | T | C | 0.0102 | 0.0018 | 9.70E-09 | 4.78E-05 | 32.1 |  | 0.0075 | 0.0389 | 8.49E-01 |
| rs4704513 | G | C | 0.0125 | 0.0022 | 7.40E-09 | 4.63E-05 | 32.3 |  | -0.0454 | 0.0469 | 3.35E-01 |
| rs9326846 | G | A | 0.0108 | 0.0019 | 7.10E-09 | 5.00E-05 | 32.3 |  | 0.0097 | 0.0391 | 8.06E-01 |
| rs9475173 | A | G | 0.0108 | 0.0019 | 6.60E-09 | 5.28E-05 | 32.3 |  | -0.0253 | 0.0381 | 5.09E-01 |
| rs1885728 | A | G | 0.0108 | 0.0019 | 1.00E-08 | 5.09E-05 | 32.3 |  | -0.0546 | 0.0403 | 1.77E-01 |
| rs2270778 | C | T | 0.0097 | 0.0017 | 4.30E-09 | 4.57E-05 | 32.6 |  | -0.0116 | 0.0378 | 7.61E-01 |
| rs10118866 | T | G | 0.012 | 0.0021 | 6.20E-09 | 5.00E-05 | 32.7 |  | -0.0526 | 0.0446 | 2.41E-01 |
| rs11089885 | C | T | 0.0103 | 0.0018 | 4.00E-09 | 5.27E-05 | 32.7 |  | -0.0276 | 0.0378 | 4.68E-01 |
| rs11790280 | C | T | 0.0103 | 0.0018 | 7.10E-09 | 5.03E-05 | 32.7 |  | -0.0326 | 0.0387 | 4.03E-01 |
| rs175165 | T | G | 0.0103 | 0.0018 | 5.20E-09 | 5.07E-05 | 32.7 |  | -0.0014 | 0.0378 | 9.71E-01 |
| rs4865796 | G | A | 0.0103 | 0.0018 | 5.20E-09 | 4.53E-05 | 32.7 |  | 0.0109 | 0.0401 | 7.87E-01 |
| rs1584121 | G | A | 0.0126 | 0.0022 | 6.00E-09 | 4.90E-05 | 32.8 |  | 0.0334 | 0.0464 | 4.74E-01 |
| rs16833232 | C | T | 0.0109 | 0.0019 | 4.60E-09 | 5.10E-05 | 32.9 |  | -0.0118 | 0.0401 | 7.70E-01 |
| rs1020548 | G | A | 0.0132 | 0.0023 | 6.60E-09 | 4.82E-05 | 32.9 |  | -0.0029 | 0.0496 | 9.54E-01 |
| rs11649864 | A | G | 0.0178 | 0.0031 | 6.70E-09 | 5.26E-05 | 33.0 |  | 0.0218 | 0.0655 | 7.41E-01 |
| rs252749 | G | A | 0.0115 | 0.002 | 8.20E-09 | 4.85E-05 | 33.1 |  | 0.0614 | 0.0440 | 1.65E-01 |
| rs2832283 | A | G | 0.0115 | 0.002 | 5.80E-09 | 4.55E-05 | 33.1 |  | -0.0117 | 0.0455 | 7.98E-01 |
| rs1402025 | C | T | 0.0121 | 0.0021 | 3.60E-09 | 5.13E-05 | 33.2 |  | 0.0116 | 0.0445 | 7.96E-01 |
| rs2143624 | A | G | 0.0098 | 0.0017 | 4.80E-09 | 4.47E-05 | 33.2 |  | 0.0356 | 0.0389 | 3.64E-01 |
| rs3829849 | T | C | 0.0098 | 0.0017 | 5.90E-09 | 4.42E-05 | 33.2 |  | -0.0109 | 0.0397 | 7.85E-01 |
| rs9547153 | G | A | 0.0098 | 0.0017 | 8.70E-09 | 4.54E-05 | 33.2 |  | 0.0226 | 0.0383 | 5.57E-01 |
| rs4430672 | T | C | 0.0127 | 0.0022 | 3.90E-09 | 5.15E-05 | 33.3 |  | 0.0381 | 0.0458 | 4.07E-01 |
| rs825680 | A | T | 0.0104 | 0.0018 | 6.70E-09 | 5.26E-05 | 33.4 |  | 0.0326 | 0.0382 | 3.95E-01 |
| rs535533 | C | T | 0.0104 | 0.0018 | 3.40E-09 | 5.22E-05 | 33.4 |  | 0.0205 | 0.0381 | 5.93E-01 |
| rs2477017 | A | G | 0.0104 | 0.0018 | 5.40E-09 | 5.10E-05 | 33.4 |  | -0.0324 | 0.0384 | 4.01E-01 |
| rs6898812 | G | T | 0.0104 | 0.0018 | 4.20E-09 | 5.33E-05 | 33.4 |  | -0.0414 | 0.0377 | 2.75E-01 |
| rs2619976 | T | C | 0.0104 | 0.0018 | 6.30E-09 | 5.25E-05 | 33.4 |  | 0.0227 | 0.0378 | 5.51E-01 |
| rs1345942 | C | T | 0.0104 | 0.0018 | 5.20E-09 | 5.05E-05 | 33.4 |  | 0.0710 | 0.0384 | 6.60E-02 |
| rs2732275 | A | G | 0.0104 | 0.0018 | 3.80E-09 | 5.12E-05 | 33.4 |  | 0.0264 | 0.0380 | 4.89E-01 |
| rs1608445 | G | A | 0.0104 | 0.0018 | 3.70E-09 | 5.32E-05 | 33.4 |  | -0.0006 | 0.0377 | 9.87E-01 |
| rs2195086 | G | T | 0.0133 | 0.0023 | 9.40E-09 | 4.73E-05 | 33.4 |  | -0.0478 | 0.0515 | 3.55E-01 |
| rs1117080 | C | G | 0.011 | 0.0019 | 7.00E-09 | 5.07E-05 | 33.5 |  | 0.0583 | 0.0409 | 1.56E-01 |
| rs1829130 | C | T | 0.011 | 0.0019 | 3.70E-09 | 5.28E-05 | 33.5 |  | 0.0479 | 0.0403 | 2.37E-01 |
| rs17105272 | T | C | 0.011 | 0.0019 | 3.20E-09 | 5.25E-05 | 33.5 |  | 0.0838 | 0.0396 | 3.52E-02 |
| rs961917 | C | G | 0.0116 | 0.002 | 2.60E-09 | 5.37E-05 | 33.6 |  | -0.0326 | 0.0412 | 4.31E-01 |
| rs10732321 | C | G | 0.0145 | 0.0025 | 5.10E-09 | 5.16E-05 | 33.6 |  | -0.0490 | 0.0525 | 3.53E-01 |
| rs925421 | A | G | 0.0116 | 0.002 | 5.80E-09 | 5.25E-05 | 33.6 |  | 0.0035 | 0.0419 | 9.34E-01 |
| rs7181610 | A | T | 0.0145 | 0.0025 | 7.80E-09 | 5.11E-05 | 33.6 |  | 0.0638 | 0.0556 | 2.54E-01 |
| rs10263780 | G | A | 0.0157 | 0.0027 | 8.30E-09 | 5.90E-05 | 33.8 |  | 0.0245 | 0.0555 | 6.61E-01 |
| rs3826705 | C | T | 0.0157 | 0.0027 | 6.60E-09 | 5.23E-05 | 33.8 |  | -0.0396 | 0.0560 | 4.82E-01 |
| rs4077093 | T | G | 0.0128 | 0.0022 | 5.10E-09 | 5.57E-05 | 33.9 |  | 0.0180 | 0.0459 | 6.97E-01 |
| rs7694732 | A | G | 0.0099 | 0.0017 | 8.70E-09 | 4.82E-05 | 33.9 |  | 0.0178 | 0.0372 | 6.33E-01 |
| rs653264 | G | A | 0.0099 | 0.0017 | 9.30E-09 | 4.88E-05 | 33.9 |  | 0.0220 | 0.0371 | 5.55E-01 |
| rs4953577 | T | C | 0.0099 | 0.0017 | 9.90E-09 | 4.85E-05 | 33.9 |  | 0.0151 | 0.0376 | 6.89E-01 |
| rs10942476 | G | A | 0.0099 | 0.0017 | 9.50E-09 | 4.90E-05 | 33.9 |  | -0.0156 | 0.0369 | 6.75E-01 |
| rs1158684 | A | G | 0.0099 | 0.0017 | 9.10E-09 | 4.90E-05 | 33.9 |  | 0.0180 | 0.0373 | 6.32E-01 |
| rs6561710 | G | A | 0.0105 | 0.0018 | 2.60E-09 | 5.36E-05 | 34.0 |  | 0.0009 | 0.0375 | 9.81E-01 |
| rs17272434 | G | A | 0.0105 | 0.0018 | 3.70E-09 | 4.73E-05 | 34.0 |  | -0.0223 | 0.0408 | 5.87E-01 |
| rs2923774 | A | G | 0.0105 | 0.0018 | 7.70E-09 | 4.96E-05 | 34.0 |  | -0.0088 | 0.0394 | 8.25E-01 |
| rs9318686 | C | T | 0.0105 | 0.0018 | 1.00E-08 | 4.93E-05 | 34.0 |  | 0.0084 | 0.0393 | 8.32E-01 |
| rs6777784 | T | G | 0.0105 | 0.0018 | 4.70E-09 | 5.18E-05 | 34.0 |  | 0.0441 | 0.0388 | 2.58E-01 |
| rs2269828 | G | A | 0.0105 | 0.0018 | 4.40E-09 | 4.89E-05 | 34.0 |  | -0.0100 | 0.0397 | 8.02E-01 |
| rs11926767 | T | C | 0.0105 | 0.0018 | 2.50E-09 | 5.26E-05 | 34.0 |  | -0.0637 | 0.0381 | 9.63E-02 |
| rs10757826 | A | G | 0.0105 | 0.0018 | 3.00E-09 | 4.84E-05 | 34.0 |  | 0.0146 | 0.0403 | 7.18E-01 |
| rs17448885 | C | G | 0.0105 | 0.0018 | 7.30E-09 | 4.99E-05 | 34.0 |  | -0.0304 | 0.0393 | 4.42E-01 |
| rs4936671 | C | G | 0.0105 | 0.0018 | 6.20E-09 | 5.12E-05 | 34.0 |  | 0.0391 | 0.0382 | 3.09E-01 |
| rs6901756 | T | C | 0.0146 | 0.0025 | 2.90E-09 | 4.57E-05 | 34.1 |  | 0.0081 | 0.0592 | 8.92E-01 |
| rs7209235 | G | A | 0.0111 | 0.0019 | 8.00E-09 | 5.22E-05 | 34.1 |  | 0.0630 | 0.0413 | 1.29E-01 |
| rs2973564 | A | G | 0.0111 | 0.0019 | 3.30E-09 | 5.01E-05 | 34.1 |  | 0.0062 | 0.0418 | 8.83E-01 |
| rs9904177 | G | A | 0.0117 | 0.002 | 2.30E-09 | 5.44E-05 | 34.2 |  | 0.0822 | 0.0419 | 5.12E-02 |
| rs9527455 | C | A | 0.0117 | 0.002 | 8.00E-09 | 4.88E-05 | 34.2 |  | 0.0447 | 0.0444 | 3.17E-01 |
| rs1941213 | A | C | 0.0117 | 0.002 | 3.20E-09 | 5.48E-05 | 34.2 |  | -0.0065 | 0.0409 | 8.75E-01 |
| rs9964756 | G | T | 0.0164 | 0.0028 | 2.90E-09 | 4.98E-05 | 34.3 |  | 0.0076 | 0.0613 | 9.02E-01 |
| rs17538472 | T | C | 0.0129 | 0.0022 | 4.20E-09 | 5.14E-05 | 34.4 |  | -0.0148 | 0.0471 | 7.54E-01 |
| rs2448241 | G | A | 0.0176 | 0.003 | 3.20E-09 | 5.43E-05 | 34.4 |  | -0.0058 | 0.0607 | 9.24E-01 |
| rs2304130 | A | G | 0.0176 | 0.003 | 2.90E-09 | 4.83E-05 | 34.4 |  | -0.1413 | 0.0629 | 2.55E-02 |
| rs12629015 | A | G | 0.0135 | 0.0023 | 2.10E-09 | 5.50E-05 | 34.5 |  | -0.0209 | 0.0470 | 6.58E-01 |
| rs11538 | G | A | 0.0135 | 0.0023 | 3.30E-09 | 5.39E-05 | 34.5 |  | -0.0031 | 0.0482 | 9.48E-01 |
| rs3764835 | G | A | 0.0141 | 0.0024 | 3.10E-09 | 5.15E-05 | 34.5 |  | 0.0433 | 0.0508 | 3.96E-01 |
| rs6968554 | G | A | 0.01 | 0.0017 | 3.50E-09 | 4.61E-05 | 34.6 |  | -0.0345 | 0.0392 | 3.81E-01 |
| rs2489676 | G | T | 0.01 | 0.0017 | 8.80E-09 | 4.95E-05 | 34.6 |  | -0.0165 | 0.0374 | 6.62E-01 |
| rs1668633 | T | C | 0.01 | 0.0017 | 8.70E-09 | 4.89E-05 | 34.6 |  | -0.0095 | 0.0379 | 8.02E-01 |
| rs17681708 | C | T | 0.0106 | 0.0018 | 7.20E-09 | 4.83E-05 | 34.7 |  | -0.0126 | 0.0409 | 7.59E-01 |
| rs1840969 | T | A | 0.0106 | 0.0018 | 1.90E-09 | 5.43E-05 | 34.7 |  | 0.0061 | 0.0376 | 8.71E-01 |
| rs9296723 | C | T | 0.0106 | 0.0018 | 4.00E-09 | 5.19E-05 | 34.7 |  | -0.0702 | 0.0389 | 7.25E-02 |
| rs17695092 | T | G | 0.0106 | 0.0018 | 3.40E-09 | 4.79E-05 | 34.7 |  | 0.0174 | 0.0399 | 6.64E-01 |
| rs903959 | A | T | 0.0106 | 0.0018 | 1.60E-09 | 5.38E-05 | 34.7 |  | -0.0289 | 0.0382 | 4.51E-01 |
| rs1524277 | C | T | 0.0106 | 0.0018 | 2.20E-09 | 5.61E-05 | 34.7 |  | -0.0347 | 0.0371 | 3.52E-01 |
| rs12628891 | C | T | 0.0112 | 0.0019 | 3.00E-09 | 5.43E-05 | 34.7 |  | 0.0066 | 0.0398 | 8.69E-01 |
| rs7636868 | G | A | 0.0112 | 0.0019 | 2.60E-09 | 5.35E-05 | 34.7 |  | -0.0037 | 0.0406 | 9.28E-01 |
| rs4954638 | A | C | 0.0118 | 0.002 | 2.90E-09 | 5.21E-05 | 34.8 |  | 0.0274 | 0.0443 | 5.39E-01 |
| rs17056301 | C | T | 0.0118 | 0.002 | 2.40E-09 | 5.41E-05 | 34.8 |  | -0.0207 | 0.0424 | 6.27E-01 |
| rs7950748 | T | A | 0.0118 | 0.002 | 4.80E-09 | 5.29E-05 | 34.8 |  | 0.0435 | 0.0429 | 3.14E-01 |
| rs6850639 | T | C | 0.0124 | 0.0021 | 1.80E-09 | 5.07E-05 | 34.9 |  | 0.0683 | 0.0470 | 1.48E-01 |
| rs1956153 | T | A | 0.013 | 0.0022 | 6.80E-09 | 4.96E-05 | 34.9 |  | -0.0293 | 0.0484 | 5.47E-01 |
| rs17599948 | A | G | 0.013 | 0.0022 | 4.60E-09 | 4.70E-05 | 34.9 |  | 0.1029 | 0.0518 | 4.84E-02 |
| rs16906845 | G | A | 0.0225 | 0.0038 | 2.20E-09 | 6.31E-05 | 35.1 |  | 0.0095 | 0.0742 | 8.99E-01 |
| rs9514131 | G | T | 0.0154 | 0.0026 | 5.60E-09 | 5.03E-05 | 35.1 |  | 0.0611 | 0.0596 | 3.08E-01 |
| rs833831 | T | G | 0.016 | 0.0027 | 2.60E-09 | 5.73E-05 | 35.1 |  | 0.0258 | 0.0546 | 6.38E-01 |
| rs7685048 | C | T | 0.0101 | 0.0017 | 4.10E-09 | 5.08E-05 | 35.3 |  | 0.0072 | 0.0373 | 8.47E-01 |
| rs1476322 | A | G | 0.0101 | 0.0017 | 5.00E-09 | 5.00E-05 | 35.3 |  | 0.0024 | 0.0375 | 9.49E-01 |
| rs243387 | G | A | 0.0101 | 0.0017 | 4.40E-09 | 5.06E-05 | 35.3 |  | 0.0714 | 0.0376 | 5.94E-02 |
| rs1394879 | C | G | 0.0101 | 0.0017 | 6.20E-09 | 4.96E-05 | 35.3 |  | -0.0028 | 0.0374 | 9.41E-01 |
| rs2371767 | C | G | 0.0107 | 0.0018 | 6.20E-09 | 4.58E-05 | 35.3 |  | -0.0539 | 0.0417 | 1.99E-01 |
| rs802460 | T | C | 0.0107 | 0.0018 | 2.80E-09 | 5.17E-05 | 35.3 |  | 0.0217 | 0.0388 | 5.78E-01 |
| rs2527380 | T | C | 0.0107 | 0.0018 | 5.10E-09 | 5.16E-05 | 35.3 |  | 0.0442 | 0.0399 | 2.70E-01 |
| rs731834 | A | C | 0.0107 | 0.0018 | 1.60E-09 | 5.72E-05 | 35.3 |  | 0.0308 | 0.0373 | 4.11E-01 |
| rs10243319 | T | C | 0.0107 | 0.0018 | 1.20E-09 | 5.47E-05 | 35.3 |  | -0.0157 | 0.0382 | 6.83E-01 |
| rs7534091 | G | A | 0.0113 | 0.0019 | 1.00E-09 | 4.80E-05 | 35.4 |  | -0.0082 | 0.0435 | 8.51E-01 |
| rs1511471 | A | G | 0.0113 | 0.0019 | 9.90E-10 | 5.53E-05 | 35.4 |  | 0.0545 | 0.0400 | 1.76E-01 |
| rs294704 | G | T | 0.0113 | 0.0019 | 4.00E-09 | 5.10E-05 | 35.4 |  | 0.0160 | 0.0415 | 7.02E-01 |
| rs9397928 | C | T | 0.0119 | 0.002 | 1.40E-09 | 5.49E-05 | 35.4 |  | 0.0374 | 0.0410 | 3.65E-01 |
| rs7117238 | G | A | 0.0131 | 0.0022 | 2.50E-09 | 4.80E-05 | 35.5 |  | 0.1452 | 0.0517 | 5.23E-03 |
| rs17720922 | T | C | 0.0131 | 0.0022 | 2.30E-09 | 5.28E-05 | 35.5 |  | -0.0609 | 0.0475 | 2.02E-01 |
| rs9817583 | A | G | 0.0137 | 0.0023 | 2.50E-09 | 5.30E-05 | 35.5 |  | 0.0354 | 0.0513 | 4.93E-01 |
| rs9318380 | A | G | 0.0143 | 0.0024 | 1.90E-09 | 5.40E-05 | 35.5 |  | 0.0734 | 0.0507 | 1.50E-01 |
| rs17757975 | T | C | 0.0143 | 0.0024 | 4.20E-09 | 5.16E-05 | 35.5 |  | 0.0743 | 0.0560 | 1.87E-01 |
| rs7770443 | A | C | 0.0149 | 0.0025 | 1.30E-09 | 5.77E-05 | 35.5 |  | 0.0087 | 0.0521 | 8.68E-01 |
| rs8126575 | T | G | 0.0149 | 0.0025 | 4.10E-09 | 5.34E-05 | 35.5 |  | 0.1225 | 0.0569 | 3.25E-02 |
| rs12449219 | G | C | 0.0155 | 0.0026 | 2.00E-09 | 5.64E-05 | 35.5 |  | 0.0309 | 0.0567 | 5.88E-01 |
| rs12675063 | T | A | 0.0156 | 0.0026 | 1.30E-09 | 4.88E-05 | 36.0 |  | 0.1117 | 0.0595 | 6.19E-02 |
| rs610634 | T | C | 0.0138 | 0.0023 | 1.00E-09 | 5.67E-05 | 36.0 |  | -0.0236 | 0.0484 | 6.28E-01 |
| rs972283 | A | G | 0.0096 | 0.0016 | 5.10E-09 | 4.60E-05 | 36.0 |  | 0.0319 | 0.0374 | 3.96E-01 |
| rs252819 | T | C | 0.0132 | 0.0022 | 4.00E-09 | 5.27E-05 | 36.0 |  | -0.0594 | 0.0491 | 2.30E-01 |
| rs7652415 | T | C | 0.0156 | 0.0026 | 9.70E-10 | 5.92E-05 | 36.0 |  | -0.0833 | 0.0547 | 1.30E-01 |
| rs6561766 | A | G | 0.0168 | 0.0028 | 1.20E-09 | 5.92E-05 | 36.0 |  | -0.1040 | 0.0559 | 6.46E-02 |
| rs2035831 | G | C | 0.0108 | 0.0018 | 5.50E-09 | 5.23E-05 | 36.0 |  | 0.0047 | 0.0393 | 9.06E-01 |
| rs1420341 | C | T | 0.0138 | 0.0023 | 2.10E-09 | 5.62E-05 | 36.0 |  | -0.0333 | 0.0467 | 4.78E-01 |
| rs11001963 | T | C | 0.0108 | 0.0018 | 1.20E-09 | 5.74E-05 | 36.0 |  | -0.0080 | 0.0373 | 8.32E-01 |
| rs12042959 | A | G | 0.0144 | 0.0024 | 3.40E-09 | 5.30E-05 | 36.0 |  | -0.0796 | 0.0520 | 1.28E-01 |
| rs7761673 | T | A | 0.0126 | 0.0021 | 1.90E-09 | 5.19E-05 | 36.0 |  | 0.0398 | 0.0463 | 3.93E-01 |
| rs16943356 | G | A | 0.0198 | 0.0033 | 3.00E-09 | 4.96E-05 | 36.0 |  | 0.1384 | 0.0763 | 7.14E-02 |
| rs11121210 | C | T | 0.0108 | 0.0018 | 1.30E-09 | 5.45E-05 | 36.0 |  | -0.0678 | 0.0389 | 8.30E-02 |
| rs3813680 | A | G | 0.0144 | 0.0024 | 1.60E-09 | 5.47E-05 | 36.0 |  | 0.0202 | 0.0513 | 6.96E-01 |
| rs2836961 | C | A | 0.0102 | 0.0017 | 1.10E-09 | 4.92E-05 | 36.0 |  | -0.0108 | 0.0379 | 7.77E-01 |
| rs896183 | A | G | 0.0102 | 0.0017 | 2.30E-09 | 5.05E-05 | 36.0 |  | -0.0314 | 0.0379 | 4.09E-01 |
| rs1467693 | A | T | 0.0114 | 0.0019 | 1.20E-09 | 5.37E-05 | 36.0 |  | 0.0118 | 0.0406 | 7.73E-01 |
| rs10491182 | T | C | 0.0187 | 0.0031 | 2.80E-09 | 4.71E-05 | 36.4 |  | -0.0527 | 0.0702 | 4.56E-01 |
| rs11138313 | A | G | 0.0169 | 0.0028 | 3.00E-09 | 5.07E-05 | 36.4 |  | 0.0593 | 0.0629 | 3.49E-01 |
| rs11792069 | A | G | 0.0145 | 0.0024 | 6.50E-10 | 5.93E-05 | 36.5 |  | -0.0570 | 0.0498 | 2.55E-01 |
| rs8071182 | A | G | 0.0133 | 0.0022 | 2.10E-09 | 5.07E-05 | 36.5 |  | -0.0069 | 0.0487 | 8.87E-01 |
| rs248139 | A | G | 0.0133 | 0.0022 | 1.20E-09 | 5.52E-05 | 36.5 |  | 0.0409 | 0.0464 | 3.81E-01 |
| rs2283093 | T | C | 0.0127 | 0.0021 | 3.10E-09 | 5.29E-05 | 36.6 |  | 0.0234 | 0.0459 | 6.12E-01 |
| rs12101393 | C | G | 0.0127 | 0.0021 | 1.50E-09 | 5.53E-05 | 36.6 |  | 0.0371 | 0.0446 | 4.08E-01 |
| rs1830074 | C | T | 0.0115 | 0.0019 | 1.40E-09 | 5.42E-05 | 36.6 |  | -0.0252 | 0.0406 | 5.37E-01 |
| rs12065553 | G | A | 0.0115 | 0.0019 | 8.90E-10 | 5.47E-05 | 36.6 |  | -0.0127 | 0.0412 | 7.59E-01 |
| rs491711 | A | C | 0.0115 | 0.0019 | 1.10E-09 | 5.72E-05 | 36.6 |  | -0.0327 | 0.0406 | 4.23E-01 |
| rs762147 | G | A | 0.0115 | 0.0019 | 3.30E-09 | 5.22E-05 | 36.6 |  | 0.0634 | 0.0429 | 1.41E-01 |
| rs17120344 | A | G | 0.0224 | 0.0037 | 1.50E-09 | 5.59E-05 | 36.7 |  | -0.0314 | 0.0804 | 6.98E-01 |
| rs17019087 | C | T | 0.0109 | 0.0018 | 7.80E-10 | 5.48E-05 | 36.7 |  | 0.0289 | 0.0393 | 4.64E-01 |
| rs6594967 | T | C | 0.0109 | 0.0018 | 3.40E-09 | 5.23E-05 | 36.7 |  | 0.0014 | 0.0398 | 9.73E-01 |
| rs10198345 | C | T | 0.0109 | 0.0018 | 2.20E-09 | 5.24E-05 | 36.7 |  | 0.0195 | 0.0400 | 6.28E-01 |
| rs9388766 | T | C | 0.0109 | 0.0018 | 7.60E-10 | 4.87E-05 | 36.7 |  | -0.0218 | 0.0406 | 5.94E-01 |
| rs12439829 | T | A | 0.0109 | 0.0018 | 1.10E-09 | 5.63E-05 | 36.7 |  | -0.0085 | 0.0376 | 8.21E-01 |
| rs13105058 | T | C | 0.0109 | 0.0018 | 3.10E-09 | 5.21E-05 | 36.7 |  | -0.0427 | 0.0397 | 2.84E-01 |
| rs11251352 | G | A | 0.0109 | 0.0018 | 7.00E-10 | 5.71E-05 | 36.7 |  | 0.0170 | 0.0380 | 6.57E-01 |
| rs6819344 | A | C | 0.0103 | 0.0017 | 6.10E-10 | 5.09E-05 | 36.7 |  | 0.0566 | 0.0375 | 1.33E-01 |
| rs1884389 | C | T | 0.0103 | 0.0017 | 4.00E-09 | 5.20E-05 | 36.7 |  | -0.0226 | 0.0376 | 5.51E-01 |
| rs11604688 | C | T | 0.0103 | 0.0017 | 3.60E-09 | 5.23E-05 | 36.7 |  | 0.1080 | 0.0377 | 4.43E-03 |
| rs1554194 | C | G | 0.0103 | 0.0017 | 3.40E-09 | 5.29E-05 | 36.7 |  | 0.0386 | 0.0377 | 3.08E-01 |
| rs9489622 | A | G | 0.0103 | 0.0017 | 2.80E-09 | 5.30E-05 | 36.7 |  | -0.0326 | 0.0372 | 3.83E-01 |
| rs4906908 | G | T | 0.0103 | 0.0017 | 2.50E-09 | 5.29E-05 | 36.7 |  | -0.0403 | 0.0376 | 2.87E-01 |
| rs138289 | A | T | 0.0103 | 0.0017 | 3.30E-09 | 5.30E-05 | 36.7 |  | 0.0295 | 0.0377 | 4.37E-01 |
| rs10733051 | A | G | 0.0097 | 0.0016 | 2.90E-09 | 4.70E-05 | 36.8 |  | 0.0621 | 0.0371 | 9.62E-02 |
| rs6827083 | G | A | 0.0097 | 0.0016 | 4.80E-09 | 4.61E-05 | 36.8 |  | 0.0095 | 0.0373 | 7.99E-01 |
| rs11581304 | C | T | 0.0182 | 0.003 | 7.10E-10 | 5.79E-05 | 36.8 |  | -0.0428 | 0.0664 | 5.21E-01 |
| rs11577094 | T | C | 0.0182 | 0.003 | 6.90E-10 | 4.94E-05 | 36.8 |  | 0.0398 | 0.0662 | 5.50E-01 |
| rs17424278 | C | A | 0.017 | 0.0028 | 1.90E-09 | 5.45E-05 | 36.9 |  | 0.0505 | 0.0614 | 4.14E-01 |
| rs2279620 | C | G | 0.017 | 0.0028 | 1.60E-09 | 5.38E-05 | 36.9 |  | -0.0247 | 0.0601 | 6.82E-01 |
| rs11173522 | A | C | 0.0128 | 0.0021 | 1.10E-09 | 5.39E-05 | 37.2 |  | 0.0324 | 0.0451 | 4.75E-01 |
| rs2023671 | G | T | 0.0122 | 0.002 | 5.80E-10 | 5.61E-05 | 37.2 |  | -0.0335 | 0.0426 | 4.34E-01 |
| rs16982345 | A | G | 0.0122 | 0.002 | 2.80E-09 | 5.56E-05 | 37.2 |  | 0.1214 | 0.0434 | 5.41E-03 |
| rs9426003 | G | A | 0.0116 | 0.0019 | 1.40E-09 | 5.60E-05 | 37.3 |  | 0.0538 | 0.0416 | 1.98E-01 |
| rs6759670 | C | A | 0.0116 | 0.0019 | 1.90E-09 | 5.37E-05 | 37.3 |  | 0.0249 | 0.0418 | 5.53E-01 |
| rs2600226 | C | T | 0.0116 | 0.0019 | 3.70E-10 | 5.95E-05 | 37.3 |  | 0.0268 | 0.0393 | 4.99E-01 |
| rs2155645 | C | T | 0.0116 | 0.0019 | 7.10E-10 | 5.19E-05 | 37.3 |  | -0.0207 | 0.0427 | 6.30E-01 |
| rs11754747 | T | C | 0.0116 | 0.0019 | 1.40E-09 | 5.19E-05 | 37.3 |  | 0.0304 | 0.0431 | 4.83E-01 |
| rs17709991 | C | T | 0.011 | 0.0018 | 2.40E-09 | 4.75E-05 | 37.3 |  | -0.0024 | 0.0421 | 9.55E-01 |
| rs40245 | A | T | 0.011 | 0.0018 | 1.90E-09 | 5.57E-05 | 37.3 |  | 0.0687 | 0.0388 | 7.89E-02 |
| rs13266989 | C | G | 0.011 | 0.0018 | 3.80E-10 | 5.81E-05 | 37.3 |  | 0.0116 | 0.0379 | 7.61E-01 |
| rs4856794 | G | T | 0.011 | 0.0018 | 1.50E-09 | 5.50E-05 | 37.3 |  | -0.1171 | 0.0395 | 3.19E-03 |
| rs424539 | G | C | 0.011 | 0.0018 | 4.90E-10 | 5.75E-05 | 37.3 |  | 0.0159 | 0.0383 | 6.81E-01 |
| rs7826312 | C | T | 0.0104 | 0.0017 | 4.90E-10 | 5.24E-05 | 37.4 |  | 0.0097 | 0.0379 | 7.99E-01 |
| rs676749 | T | A | 0.0104 | 0.0017 | 1.70E-09 | 5.41E-05 | 37.4 |  | 0.0131 | 0.0373 | 7.26E-01 |
| rs3806116 | T | G | 0.0104 | 0.0017 | 7.70E-10 | 5.11E-05 | 37.4 |  | -0.0812 | 0.0381 | 3.42E-02 |
| rs6121381 | T | A | 0.0147 | 0.0024 | 9.40E-10 | 5.60E-05 | 37.5 |  | -0.0259 | 0.0515 | 6.17E-01 |
| rs8016771 | G | T | 0.019 | 0.0031 | 1.10E-09 | 5.77E-05 | 37.6 |  | -0.0395 | 0.0639 | 5.39E-01 |
| rs8081039 | T | C | 0.0233 | 0.0038 | 6.10E-10 | 5.87E-05 | 37.6 |  | -0.0230 | 0.0769 | 7.66E-01 |
| rs6564360 | G | A | 0.0135 | 0.0022 | 1.20E-09 | 5.62E-05 | 37.7 |  | 0.0077 | 0.0476 | 8.72E-01 |
| rs7674623 | T | C | 0.0135 | 0.0022 | 5.70E-10 | 5.78E-05 | 37.7 |  | 0.1251 | 0.0467 | 7.73E-03 |
| rs2809395 | A | T | 0.0135 | 0.0022 | 2.20E-09 | 5.37E-05 | 37.7 |  | 0.0680 | 0.0485 | 1.63E-01 |
| rs4877313 | A | T | 0.0129 | 0.0021 | 4.10E-10 | 5.68E-05 | 37.7 |  | -0.0028 | 0.0443 | 9.50E-01 |
| rs965961 | A | G | 0.0172 | 0.0028 | 6.90E-10 | 5.45E-05 | 37.7 |  | 0.0956 | 0.0636 | 1.35E-01 |
| rs7134628 | A | G | 0.0172 | 0.0028 | 1.20E-09 | 5.45E-05 | 37.7 |  | -0.0087 | 0.0581 | 8.82E-01 |
| rs4523610 | C | T | 0.0123 | 0.002 | 1.30E-09 | 5.34E-05 | 37.8 |  | -0.0109 | 0.0437 | 8.04E-01 |
| rs17446091 | C | T | 0.0123 | 0.002 | 1.80E-09 | 4.97E-05 | 37.8 |  | -0.0346 | 0.0458 | 4.52E-01 |
| rs4518345 | G | A | 0.0117 | 0.0019 | 1.00E-09 | 5.57E-05 | 37.9 |  | 0.0187 | 0.0416 | 6.54E-01 |
| rs1911746 | T | C | 0.0117 | 0.0019 | 1.60E-09 | 4.60E-05 | 37.9 |  | -0.0549 | 0.0468 | 2.43E-01 |
| rs1853639 | G | A | 0.0111 | 0.0018 | 1.20E-09 | 5.70E-05 | 38.0 |  | -0.0232 | 0.0392 | 5.57E-01 |
| rs6556301 | G | T | 0.0111 | 0.0018 | 4.10E-10 | 5.67E-05 | 38.0 |  | -0.0192 | 0.0394 | 6.29E-01 |
| rs2451746 | C | G | 0.0111 | 0.0018 | 4.60E-10 | 5.74E-05 | 38.0 |  | -0.0571 | 0.0391 | 1.46E-01 |
| rs10408013 | T | C | 0.0111 | 0.0018 | 8.00E-10 | 5.08E-05 | 38.0 |  | 0.0124 | 0.0420 | 7.69E-01 |
| rs11670142 | G | T | 0.0111 | 0.0018 | 4.70E-10 | 5.89E-05 | 38.0 |  | 0.0333 | 0.0380 | 3.83E-01 |
| rs2429150 | C | A | 0.0111 | 0.0018 | 2.70E-10 | 5.99E-05 | 38.0 |  | 0.0317 | 0.0379 | 4.07E-01 |
| rs10830452 | G | A | 0.0111 | 0.0018 | 2.10E-09 | 5.44E-05 | 38.0 |  | 0.0565 | 0.0395 | 1.55E-01 |
| rs10745785 | C | T | 0.0111 | 0.0018 | 1.20E-09 | 5.47E-05 | 38.0 |  | -0.0270 | 0.0390 | 4.91E-01 |
| rs1260326 | C | T | 0.0105 | 0.0017 | 3.90E-10 | 5.30E-05 | 38.1 |  | -0.0002 | 0.0384 | 9.95E-01 |
| rs9845966 | T | G | 0.0105 | 0.0017 | 2.50E-10 | 5.46E-05 | 38.1 |  | -0.0154 | 0.0372 | 6.82E-01 |
| rs4670627 | T | C | 0.0105 | 0.0017 | 2.60E-10 | 5.43E-05 | 38.1 |  | -0.0379 | 0.0374 | 3.13E-01 |
| rs2293605 | C | T | 0.0167 | 0.0027 | 3.60E-10 | 6.34E-05 | 38.3 |  | -0.0595 | 0.0549 | 2.81E-01 |
| rs1363695 | C | T | 0.013 | 0.0021 | 3.30E-10 | 6.03E-05 | 38.3 |  | -0.0724 | 0.0433 | 9.62E-02 |
| rs7615297 | C | G | 0.0149 | 0.0024 | 5.70E-10 | 5.55E-05 | 38.5 |  | 0.0086 | 0.0541 | 8.74E-01 |
| rs7600699 | G | C | 0.0149 | 0.0024 | 6.40E-10 | 5.90E-05 | 38.5 |  | 0.0167 | 0.0508 | 7.44E-01 |
| rs2874800 | G | A | 0.0118 | 0.0019 | 2.60E-10 | 6.07E-05 | 38.6 |  | -0.0610 | 0.0394 | 1.24E-01 |
| rs10915840 | G | A | 0.0118 | 0.0019 | 1.30E-09 | 5.65E-05 | 38.6 |  | -0.0267 | 0.0418 | 5.25E-01 |
| rs6921533 | T | C | 0.0118 | 0.0019 | 5.60E-10 | 5.71E-05 | 38.6 |  | 0.0481 | 0.0408 | 2.41E-01 |
| rs460799 | G | A | 0.0118 | 0.0019 | 6.40E-10 | 5.46E-05 | 38.6 |  | -0.0096 | 0.0417 | 8.20E-01 |
| rs6781254 | T | C | 0.0112 | 0.0018 | 4.90E-10 | 5.30E-05 | 38.7 |  | 0.0075 | 0.0407 | 8.54E-01 |
| rs13250058 | T | G | 0.0112 | 0.0018 | 2.90E-10 | 5.49E-05 | 38.7 |  | 0.0088 | 0.0405 | 8.29E-01 |
| rs223051 | T | C | 0.0112 | 0.0018 | 1.10E-09 | 5.50E-05 | 38.7 |  | -0.0102 | 0.0395 | 7.97E-01 |
| rs2718786 | A | G | 0.0112 | 0.0018 | 2.70E-10 | 5.92E-05 | 38.7 |  | 0.0785 | 0.0385 | 4.30E-02 |
| rs1937684 | A | T | 0.0112 | 0.0018 | 8.70E-10 | 5.64E-05 | 38.7 |  | 0.0504 | 0.0395 | 2.05E-01 |
| rs13298062 | A | G | 0.0137 | 0.0022 | 3.80E-10 | 6.05E-05 | 38.8 |  | 0.0264 | 0.0463 | 5.71E-01 |
| rs1455137 | C | A | 0.0106 | 0.0017 | 4.40E-10 | 5.30E-05 | 38.9 |  | 0.0218 | 0.0380 | 5.68E-01 |
| rs2317299 | T | C | 0.0106 | 0.0017 | 1.30E-09 | 5.54E-05 | 38.9 |  | 0.0472 | 0.0374 | 2.09E-01 |
| rs223391 | G | A | 0.0131 | 0.0021 | 1.60E-10 | 5.38E-05 | 38.9 |  | -0.0075 | 0.0462 | 8.72E-01 |
| rs213533 | C | A | 0.0156 | 0.0025 | 4.80E-10 | 5.96E-05 | 38.9 |  | 0.0221 | 0.0549 | 6.89E-01 |
| rs1465900 | A | C | 0.0125 | 0.002 | 4.80E-10 | 5.34E-05 | 39.1 |  | 0.0358 | 0.0446 | 4.26E-01 |
| rs3769948 | A | G | 0.0125 | 0.002 | 2.90E-10 | 5.81E-05 | 39.1 |  | 0.0505 | 0.0419 | 2.31E-01 |
| rs4911442 | A | G | 0.015 | 0.0024 | 5.40E-10 | 5.00E-05 | 39.1 |  | -0.0212 | 0.0582 | 7.18E-01 |
| rs17619973 | A | G | 0.02 | 0.0032 | 2.40E-10 | 5.52E-05 | 39.1 |  | 0.0215 | 0.0717 | 7.66E-01 |
| rs420158 | C | T | 0.0119 | 0.0019 | 2.10E-10 | 5.53E-05 | 39.2 |  | 0.0073 | 0.0428 | 8.66E-01 |
| rs7123876 | C | T | 0.0119 | 0.0019 | 2.20E-10 | 5.25E-05 | 39.2 |  | 0.0852 | 0.0428 | 4.79E-02 |
| rs7929418 | T | C | 0.0113 | 0.0018 | 1.70E-10 | 6.06E-05 | 39.4 |  | -0.0059 | 0.0380 | 8.78E-01 |
| rs1899898 | T | C | 0.0113 | 0.0018 | 6.80E-10 | 5.56E-05 | 39.4 |  | -0.0129 | 0.0402 | 7.50E-01 |
| rs17573940 | G | A | 0.0113 | 0.0018 | 8.00E-10 | 5.28E-05 | 39.4 |  | 0.0138 | 0.0412 | 7.39E-01 |
| rs10989568 | A | G | 0.0107 | 0.0017 | 6.60E-10 | 5.69E-05 | 39.6 |  | 0.0170 | 0.0376 | 6.52E-01 |
| rs621042 | C | A | 0.0107 | 0.0017 | 6.30E-10 | 5.67E-05 | 39.6 |  | -0.0152 | 0.0374 | 6.86E-01 |
| rs2198679 | A | G | 0.0107 | 0.0017 | 5.60E-10 | 5.70E-05 | 39.6 |  | 0.0131 | 0.0373 | 7.27E-01 |
| rs2450444 | G | A | 0.0107 | 0.0017 | 3.70E-10 | 5.20E-05 | 39.6 |  | 0.0254 | 0.0394 | 5.21E-01 |
| rs702820 | C | T | 0.017 | 0.0027 | 1.90E-10 | 5.91E-05 | 39.6 |  | 0.0099 | 0.0580 | 8.66E-01 |
| rs1304070 | A | G | 0.0126 | 0.002 | 3.70E-10 | 5.77E-05 | 39.7 |  | -0.0324 | 0.0436 | 4.60E-01 |
| rs459552 | T | A | 0.0126 | 0.002 | 1.70E-10 | 5.64E-05 | 39.7 |  | 0.0409 | 0.0442 | 3.57E-01 |
| rs17450772 | C | T | 0.0189 | 0.003 | 2.90E-10 | 7.13E-05 | 39.7 |  | -0.0522 | 0.0595 | 3.83E-01 |
| rs9806058 | A | T | 0.0164 | 0.0026 | 4.00E-10 | 5.80E-05 | 39.8 |  | -0.0067 | 0.0566 | 9.07E-01 |
| rs9848399 | A | G | 0.0164 | 0.0026 | 2.10E-10 | 5.87E-05 | 39.8 |  | 0.0187 | 0.0552 | 7.37E-01 |
| rs4916229 | G | C | 0.0183 | 0.0029 | 4.90E-10 | 5.63E-05 | 39.8 |  | 0.0003 | 0.0628 | 9.96E-01 |
| rs11118308 | A | G | 0.0101 | 0.0016 | 4.80E-10 | 5.08E-05 | 39.8 |  | 0.0014 | 0.0374 | 9.71E-01 |
| rs249292 | T | C | 0.012 | 0.0019 | 1.90E-10 | 6.17E-05 | 39.9 |  | 0.0611 | 0.0414 | 1.42E-01 |
| rs774211 | T | C | 0.0139 | 0.0022 | 4.60E-10 | 5.11E-05 | 39.9 |  | 0.0050 | 0.0518 | 9.24E-01 |
| rs4722398 | T | C | 0.0158 | 0.0025 | 3.60E-10 | 5.78E-05 | 39.9 |  | -0.0132 | 0.0542 | 8.08E-01 |
| rs7865157 | T | C | 0.0177 | 0.0028 | 3.70E-10 | 5.98E-05 | 40.0 |  | -0.0562 | 0.0589 | 3.43E-01 |
| rs17608150 | T | C | 0.0196 | 0.0031 | 1.30E-10 | 5.35E-05 | 40.0 |  | -0.0121 | 0.0713 | 8.66E-01 |
| rs2837398 | C | A | 0.0114 | 0.0018 | 1.10E-10 | 6.26E-05 | 40.1 |  | -0.0733 | 0.0377 | 5.35E-02 |
| rs4814512 | A | C | 0.0133 | 0.0021 | 2.30E-10 | 6.03E-05 | 40.1 |  | 0.0657 | 0.0444 | 1.41E-01 |
| rs4663629 | A | G | 0.0133 | 0.0021 | 1.10E-10 | 6.42E-05 | 40.1 |  | 0.0249 | 0.0440 | 5.74E-01 |
| rs10163018 | C | T | 0.0114 | 0.0018 | 1.50E-10 | 6.09E-05 | 40.1 |  | 0.0112 | 0.0383 | 7.71E-01 |
| rs13201877 | G | A | 0.0152 | 0.0024 | 2.70E-10 | 5.56E-05 | 40.1 |  | -0.0025 | 0.0543 | 9.63E-01 |
| rs16940823 | C | A | 0.0146 | 0.0023 | 9.00E-11 | 6.57E-05 | 40.3 |  | 0.0217 | 0.0471 | 6.48E-01 |
| rs779206 | G | A | 0.0127 | 0.002 | 1.10E-10 | 6.30E-05 | 40.3 |  | 0.0265 | 0.0425 | 5.35E-01 |
| rs9463175 | C | T | 0.0108 | 0.0017 | 4.20E-10 | 5.30E-05 | 40.4 |  | 0.0309 | 0.0387 | 4.27E-01 |
| rs6852276 | G | A | 0.0108 | 0.0017 | 8.50E-11 | 5.61E-05 | 40.4 |  | 0.0689 | 0.0377 | 6.93E-02 |
| rs11577179 | G | A | 0.0108 | 0.0017 | 1.50E-10 | 5.45E-05 | 40.4 |  | -0.0194 | 0.0388 | 6.18E-01 |
| rs4783241 | G | C | 0.0108 | 0.0017 | 4.00E-10 | 5.83E-05 | 40.4 |  | -0.0015 | 0.0373 | 9.68E-01 |
| rs16965225 | T | G | 0.0216 | 0.0034 | 1.70E-10 | 5.87E-05 | 40.4 |  | 0.0954 | 0.0767 | 2.16E-01 |
| rs10009336 | C | T | 0.014 | 0.0022 | 2.20E-10 | 5.37E-05 | 40.5 |  | 0.0239 | 0.0496 | 6.32E-01 |
| rs7334078 | T | C | 0.0121 | 0.0019 | 2.20E-10 | 6.01E-05 | 40.6 |  | 0.0915 | 0.0414 | 2.80E-02 |
| rs11115176 | T | C | 0.0121 | 0.0019 | 2.00E-10 | 5.34E-05 | 40.6 |  | 0.0133 | 0.0441 | 7.64E-01 |
| rs2174367 | G | T | 0.0121 | 0.0019 | 9.30E-11 | 6.30E-05 | 40.6 |  | -0.0063 | 0.0408 | 8.77E-01 |
| rs7802342 | G | T | 0.0121 | 0.0019 | 3.10E-10 | 6.02E-05 | 40.6 |  | -0.0166 | 0.0410 | 6.88E-01 |
| rs2170382 | T | C | 0.0172 | 0.0027 | 2.40E-10 | 6.26E-05 | 40.6 |  | -0.0408 | 0.0596 | 4.96E-01 |
| rs3851083 | G | A | 0.0102 | 0.0016 | 4.10E-10 | 5.11E-05 | 40.6 |  | -0.0254 | 0.0375 | 5.01E-01 |
| rs17236194 | C | T | 0.0153 | 0.0024 | 8.30E-11 | 5.45E-05 | 40.6 |  | 0.0654 | 0.0544 | 2.32E-01 |
| rs4072917 | A | G | 0.0115 | 0.0018 | 6.90E-11 | 6.59E-05 | 40.8 |  | 0.0335 | 0.0375 | 3.74E-01 |
| rs1863652 | G | A | 0.0115 | 0.0018 | 1.40E-10 | 5.98E-05 | 40.8 |  | -0.0142 | 0.0398 | 7.23E-01 |
| rs11917965 | C | A | 0.0115 | 0.0018 | 7.20E-11 | 6.30E-05 | 40.8 |  | 0.0030 | 0.0384 | 9.38E-01 |
| rs852056 | T | C | 0.0128 | 0.002 | 1.80E-10 | 6.00E-05 | 41.0 |  | -0.0070 | 0.0441 | 8.74E-01 |
| rs1865341 | T | C | 0.0128 | 0.002 | 2.50E-10 | 6.01E-05 | 41.0 |  | 0.0516 | 0.0431 | 2.35E-01 |
| rs17551974 | C | A | 0.0141 | 0.0022 | 1.90E-10 | 5.82E-05 | 41.1 |  | 0.0551 | 0.0486 | 2.60E-01 |
| rs12597712 | G | C | 0.0109 | 0.0017 | 1.40E-10 | 5.79E-05 | 41.1 |  | -0.0135 | 0.0375 | 7.20E-01 |
| rs7560871 | A | G | 0.0218 | 0.0034 | 9.60E-11 | 6.46E-05 | 41.1 |  | 0.0095 | 0.0721 | 8.96E-01 |
| rs7805441 | T | C | 0.0109 | 0.0017 | 3.40E-10 | 5.94E-05 | 41.1 |  | -0.0327 | 0.0375 | 3.86E-01 |
| rs8036040 | A | C | 0.0109 | 0.0017 | 2.70E-10 | 5.94E-05 | 41.1 |  | 0.0540 | 0.0373 | 1.50E-01 |
| rs2470520 | C | T | 0.0109 | 0.0017 | 5.00E-10 | 5.69E-05 | 41.1 |  | -0.0290 | 0.0377 | 4.45E-01 |
| rs5751239 | T | C | 0.0109 | 0.0017 | 3.60E-10 | 5.93E-05 | 41.1 |  | -0.0750 | 0.0372 | 4.49E-02 |
| rs7006629 | T | C | 0.0109 | 0.0017 | 2.50E-10 | 5.92E-05 | 41.1 |  | 0.0973 | 0.0369 | 8.76E-03 |
| rs2282802 | A | G | 0.0109 | 0.0017 | 4.00E-10 | 5.87E-05 | 41.1 |  | -0.0640 | 0.0373 | 8.82E-02 |
| rs784944 | A | G | 0.0122 | 0.0019 | 3.90E-10 | 5.60E-05 | 41.2 |  | -0.0566 | 0.0433 | 1.94E-01 |
| rs1544459 | C | T | 0.0103 | 0.0016 | 2.20E-10 | 5.26E-05 | 41.4 |  | -0.0655 | 0.0371 | 7.93E-02 |
| rs2605603 | G | A | 0.0103 | 0.0016 | 2.50E-10 | 5.30E-05 | 41.4 |  | -0.0017 | 0.0375 | 9.63E-01 |
| rs4802778 | G | A | 0.0116 | 0.0018 | 8.40E-11 | 6.58E-05 | 41.5 |  | -0.0159 | 0.0378 | 6.77E-01 |
| rs6764533 | A | G | 0.0116 | 0.0018 | 1.40E-10 | 6.19E-05 | 41.5 |  | 0.0217 | 0.0384 | 5.76E-01 |
| rs755407 | T | C | 0.0116 | 0.0018 | 2.30E-10 | 5.44E-05 | 41.5 |  | 0.0433 | 0.0412 | 2.97E-01 |
| rs10803762 | A | G | 0.0116 | 0.0018 | 3.50E-10 | 5.87E-05 | 41.5 |  | 0.0448 | 0.0401 | 2.67E-01 |
| rs159032 | T | C | 0.0129 | 0.002 | 1.90E-10 | 6.14E-05 | 41.6 |  | 0.0911 | 0.0423 | 3.23E-02 |
| rs2815324 | C | T | 0.0129 | 0.002 | 1.00E-10 | 5.65E-05 | 41.6 |  | -0.0212 | 0.0440 | 6.32E-01 |
| rs2866816 | T | C | 0.0129 | 0.002 | 5.60E-11 | 6.41E-05 | 41.6 |  | 0.0121 | 0.0422 | 7.77E-01 |
| rs12148330 | A | T | 0.0142 | 0.0022 | 1.50E-10 | 5.98E-05 | 41.7 |  | -0.0436 | 0.0486 | 3.72E-01 |
| rs2074881 | C | T | 0.0155 | 0.0024 | 4.90E-11 | 5.23E-05 | 41.7 |  | 0.0934 | 0.0520 | 7.44E-02 |
| rs1523768 | G | A | 0.011 | 0.0017 | 2.60E-10 | 5.27E-05 | 41.9 |  | -0.0341 | 0.0399 | 3.96E-01 |
| rs9615905 | T | C | 0.011 | 0.0017 | 2.70E-10 | 5.99E-05 | 41.9 |  | 0.0163 | 0.0377 | 6.66E-01 |
| rs331949 | C | T | 0.011 | 0.0017 | 1.00E-10 | 5.66E-05 | 41.9 |  | -0.0850 | 0.0385 | 2.80E-02 |
| rs9361779 | A | C | 0.011 | 0.0017 | 1.60E-10 | 5.97E-05 | 41.9 |  | 0.0605 | 0.0375 | 1.09E-01 |
| rs7220138 | G | C | 0.0123 | 0.0019 | 2.70E-10 | 6.40E-05 | 41.9 |  | -0.0922 | 0.0412 | 2.61E-02 |
| rs11170468 | A | C | 0.0123 | 0.0019 | 1.90E-10 | 5.40E-05 | 41.9 |  | -0.0193 | 0.0430 | 6.56E-01 |
| rs5742914 | T | C | 0.0175 | 0.0027 | 8.80E-11 | 7.17E-05 | 42.0 |  | 0.0657 | 0.0545 | 2.31E-01 |
| rs10058464 | C | A | 0.0175 | 0.0027 | 1.30E-10 | 6.09E-05 | 42.0 |  | -0.0773 | 0.0603 | 2.03E-01 |
| rs13432055 | C | T | 0.0117 | 0.0018 | 8.90E-11 | 5.59E-05 | 42.3 |  | 0.0221 | 0.0415 | 5.97E-01 |
| rs1485038 | T | C | 0.0143 | 0.0022 | 7.40E-11 | 6.63E-05 | 42.3 |  | -0.0143 | 0.0463 | 7.59E-01 |
| rs1526665 | C | T | 0.0117 | 0.0018 | 4.70E-11 | 6.61E-05 | 42.3 |  | 0.0350 | 0.0379 | 3.58E-01 |
| rs11739877 | T | C | 0.0117 | 0.0018 | 6.60E-11 | 6.50E-05 | 42.3 |  | 0.0309 | 0.0379 | 4.18E-01 |
| rs4523552 | T | C | 0.0137 | 0.0021 | 4.00E-11 | 6.50E-05 | 42.6 |  | -0.0001 | 0.0437 | 9.98E-01 |
| rs4796243 | G | A | 0.0124 | 0.0019 | 6.10E-11 | 6.49E-05 | 42.6 |  | -0.0683 | 0.0412 | 9.92E-02 |
| rs4858887 | C | A | 0.0124 | 0.0019 | 2.10E-11 | 6.68E-05 | 42.6 |  | -0.0882 | 0.0406 | 3.10E-02 |
| rs10811901 | A | G | 0.0111 | 0.0017 | 2.50E-11 | 6.07E-05 | 42.6 |  | -0.0167 | 0.0376 | 6.59E-01 |
| rs1903579 | C | G | 0.0111 | 0.0017 | 1.10E-10 | 6.10E-05 | 42.6 |  | 0.0230 | 0.0373 | 5.39E-01 |
| rs11075489 | C | T | 0.0111 | 0.0017 | 1.30E-10 | 6.15E-05 | 42.6 |  | -0.0158 | 0.0374 | 6.75E-01 |
| rs1399471 | G | C | 0.0131 | 0.002 | 2.70E-11 | 6.62E-05 | 42.9 |  | 0.0035 | 0.0425 | 9.34E-01 |
| rs12905439 | C | G | 0.0118 | 0.0018 | 1.40E-10 | 6.24E-05 | 43.0 |  | -0.0082 | 0.0389 | 8.34E-01 |
| rs7784465 | C | T | 0.0164 | 0.0025 | 1.10E-10 | 6.45E-05 | 43.0 |  | 0.0531 | 0.0541 | 3.30E-01 |
| rs7748777 | A | G | 0.0105 | 0.0016 | 1.60E-10 | 5.47E-05 | 43.1 |  | 0.0268 | 0.0376 | 4.78E-01 |
| rs17001561 | A | G | 0.0151 | 0.0023 | 3.80E-11 | 6.04E-05 | 43.1 |  | -0.0648 | 0.0521 | 2.16E-01 |
| rs11753081 | T | G | 0.0138 | 0.0021 | 8.90E-11 | 5.50E-05 | 43.2 |  | -0.0930 | 0.0479 | 5.37E-02 |
| rs6023633 | G | A | 0.0138 | 0.0021 | 5.10E-11 | 6.89E-05 | 43.2 |  | -0.0440 | 0.0439 | 3.18E-01 |
| rs2616192 | T | G | 0.0125 | 0.0019 | 2.10E-11 | 6.83E-05 | 43.3 |  | 0.0117 | 0.0391 | 7.67E-01 |
| rs12149756 | G | A | 0.0125 | 0.0019 | 3.70E-11 | 5.94E-05 | 43.3 |  | -0.0054 | 0.0431 | 9.01E-01 |
| rs1787267 | G | C | 0.0237 | 0.0036 | 3.40E-11 | 6.60E-05 | 43.3 |  | -0.0382 | 0.0727 | 6.01E-01 |
| rs11781222 | T | C | 0.0158 | 0.0024 | 5.30E-11 | 5.60E-05 | 43.3 |  | 0.0430 | 0.0594 | 4.72E-01 |
| rs486359 | C | G | 0.0112 | 0.0017 | 1.60E-11 | 6.27E-05 | 43.4 |  | -0.0090 | 0.0372 | 8.09E-01 |
| rs7941030 | C | T | 0.0112 | 0.0017 | 2.00E-11 | 5.95E-05 | 43.4 |  | -0.0429 | 0.0382 | 2.64E-01 |
| rs823074 | T | C | 0.0112 | 0.0017 | 1.60E-10 | 6.08E-05 | 43.4 |  | 0.0207 | 0.0374 | 5.82E-01 |
| rs4759075 | T | C | 0.0112 | 0.0017 | 1.40E-11 | 6.03E-05 | 43.4 |  | 0.0270 | 0.0378 | 4.78E-01 |
| rs9688431 | T | C | 0.0231 | 0.0035 | 2.40E-11 | 6.05E-05 | 43.6 |  | 0.0257 | 0.0753 | 7.34E-01 |
| rs2425840 | C | A | 0.0119 | 0.0018 | 1.60E-11 | 6.83E-05 | 43.7 |  | -0.0178 | 0.0383 | 6.44E-01 |
| rs6767619 | C | G | 0.0119 | 0.0018 | 8.10E-11 | 6.40E-05 | 43.7 |  | -0.0002 | 0.0395 | 9.95E-01 |
| rs1912631 | G | A | 0.0119 | 0.0018 | 1.30E-11 | 6.84E-05 | 43.7 |  | -0.0536 | 0.0381 | 1.62E-01 |
| rs1371108 | A | C | 0.0119 | 0.0018 | 9.00E-11 | 6.21E-05 | 43.7 |  | -0.0296 | 0.0396 | 4.58E-01 |
| rs10883553 | A | C | 0.0119 | 0.0018 | 1.90E-11 | 7.01E-05 | 43.7 |  | 0.0464 | 0.0371 | 2.14E-01 |
| rs7147503 | C | T | 0.0119 | 0.0018 | 2.00E-11 | 6.60E-05 | 43.7 |  | -0.0212 | 0.0385 | 5.84E-01 |
| rs1187352 | C | T | 0.0119 | 0.0018 | 6.00E-11 | 6.43E-05 | 43.7 |  | 0.0112 | 0.0392 | 7.77E-01 |
| rs6661316 | T | C | 0.0106 | 0.0016 | 6.80E-11 | 5.49E-05 | 43.9 |  | -0.0501 | 0.0375 | 1.84E-01 |
| rs7973955 | G | A | 0.0126 | 0.0019 | 3.70E-11 | 6.47E-05 | 44.0 |  | -0.0300 | 0.0413 | 4.71E-01 |
| rs8089514 | A | T | 0.0126 | 0.0019 | 1.10E-11 | 7.31E-05 | 44.0 |  | 0.0270 | 0.0391 | 4.93E-01 |
| rs1006353 | A | G | 0.0126 | 0.0019 | 2.30E-11 | 5.93E-05 | 44.0 |  | -0.0338 | 0.0423 | 4.26E-01 |
| rs7249143 | T | G | 0.0126 | 0.0019 | 3.60E-11 | 6.77E-05 | 44.0 |  | -0.0643 | 0.0413 | 1.21E-01 |
| rs2543132 | C | G | 0.0146 | 0.0022 | 5.00E-11 | 6.47E-05 | 44.0 |  | 0.0501 | 0.0482 | 3.02E-01 |
| rs10461497 | T | C | 0.0113 | 0.0017 | 5.40E-11 | 6.35E-05 | 44.2 |  | 0.0035 | 0.0375 | 9.27E-01 |
| rs2100814 | A | G | 0.0113 | 0.0017 | 1.20E-10 | 6.18E-05 | 44.2 |  | -0.0488 | 0.0384 | 2.07E-01 |
| rs1128249 | T | G | 0.0113 | 0.0017 | 8.50E-12 | 6.13E-05 | 44.2 |  | -0.0316 | 0.0377 | 4.04E-01 |
| rs2012502 | A | C | 0.0113 | 0.0017 | 4.20E-11 | 5.96E-05 | 44.2 |  | 0.0715 | 0.0384 | 6.42E-02 |
| rs4989244 | G | A | 0.0113 | 0.0017 | 7.50E-11 | 6.18E-05 | 44.2 |  | 0.0536 | 0.0376 | 1.57E-01 |
| rs761423 | T | C | 0.0113 | 0.0017 | 5.50E-11 | 6.32E-05 | 44.2 |  | -0.0080 | 0.0375 | 8.32E-01 |
| rs12206564 | C | T | 0.0113 | 0.0017 | 5.40E-11 | 6.38E-05 | 44.2 |  | -0.0185 | 0.0372 | 6.22E-01 |
| rs13168288 | A | G | 0.0133 | 0.002 | 6.40E-11 | 6.64E-05 | 44.2 |  | -0.0125 | 0.0428 | 7.71E-01 |
| rs980329 | C | T | 0.0133 | 0.002 | 2.50E-11 | 6.60E-05 | 44.2 |  | 0.0269 | 0.0426 | 5.31E-01 |
| rs6804181 | A | T | 0.0153 | 0.0023 | 5.50E-11 | 6.87E-05 | 44.3 |  | -0.0295 | 0.0496 | 5.55E-01 |
| rs6849518 | T | C | 0.0173 | 0.0026 | 2.60E-11 | 6.61E-05 | 44.3 |  | 0.0056 | 0.0555 | 9.20E-01 |
| rs12779943 | T | C | 0.0213 | 0.0032 | 2.00E-11 | 6.84E-05 | 44.3 |  | 0.0382 | 0.0694 | 5.84E-01 |
| rs2063177 | A | G | 0.012 | 0.0018 | 1.60E-11 | 6.59E-05 | 44.4 |  | 0.0130 | 0.0390 | 7.41E-01 |
| rs13425435 | A | C | 0.012 | 0.0018 | 2.70E-11 | 6.39E-05 | 44.4 |  | 0.0386 | 0.0398 | 3.35E-01 |
| rs1634350 | A | C | 0.012 | 0.0018 | 7.60E-12 | 7.02E-05 | 44.4 |  | -0.0119 | 0.0375 | 7.53E-01 |
| rs17776719 | G | A | 0.0167 | 0.0025 | 1.70E-11 | 6.95E-05 | 44.6 |  | -0.0119 | 0.0526 | 8.23E-01 |
| rs10438964 | C | T | 0.0127 | 0.0019 | 7.60E-11 | 6.44E-05 | 44.7 |  | 0.0212 | 0.0418 | 6.15E-01 |
| rs4682718 | G | A | 0.0154 | 0.0023 | 9.90E-12 | 7.00E-05 | 44.8 |  | 0.0611 | 0.0474 | 2.00E-01 |
| rs10499275 | C | G | 0.0154 | 0.0023 | 3.40E-11 | 6.54E-05 | 44.8 |  | 0.0117 | 0.0487 | 8.11E-01 |
| rs3902951 | G | T | 0.0134 | 0.002 | 7.00E-12 | 6.65E-05 | 44.9 |  | 0.0544 | 0.0428 | 2.06E-01 |
| rs12422552 | G | C | 0.0134 | 0.002 | 1.60E-11 | 7.02E-05 | 44.9 |  | -0.0089 | 0.0431 | 8.37E-01 |
| rs13209968 | C | G | 0.0114 | 0.0017 | 3.10E-11 | 6.49E-05 | 45.0 |  | -0.0364 | 0.0374 | 3.33E-01 |
| rs4745794 | G | A | 0.0114 | 0.0017 | 7.20E-11 | 6.49E-05 | 45.0 |  | 0.0811 | 0.0371 | 2.99E-02 |
| rs1430387 | T | C | 0.0114 | 0.0017 | 5.80E-11 | 6.37E-05 | 45.0 |  | 0.0216 | 0.0378 | 5.71E-01 |
| rs10768994 | T | C | 0.0114 | 0.0017 | 6.40E-12 | 6.38E-05 | 45.0 |  | -0.0893 | 0.0376 | 1.84E-02 |
| rs7683836 | G | A | 0.0114 | 0.0017 | 6.30E-11 | 6.46E-05 | 45.0 |  | -0.0607 | 0.0377 | 1.09E-01 |
| rs10818938 | A | G | 0.0114 | 0.0017 | 5.10E-11 | 6.34E-05 | 45.0 |  | -0.0589 | 0.0375 | 1.18E-01 |
| rs1658820 | T | G | 0.0141 | 0.0021 | 5.90E-12 | 7.39E-05 | 45.1 |  | 0.0022 | 0.0432 | 9.60E-01 |
| rs12448738 | C | A | 0.0168 | 0.0025 | 2.80E-11 | 6.68E-05 | 45.2 |  | -0.0603 | 0.0553 | 2.78E-01 |
| rs2237403 | C | T | 0.0121 | 0.0018 | 3.30E-11 | 6.59E-05 | 45.2 |  | -0.0634 | 0.0391 | 1.07E-01 |
| rs9299 | T | C | 0.0121 | 0.0018 | 3.60E-11 | 6.69E-05 | 45.2 |  | 0.1024 | 0.0391 | 9.22E-03 |
| rs11129662 | G | A | 0.0121 | 0.0018 | 4.50E-11 | 6.49E-05 | 45.2 |  | 0.0845 | 0.0386 | 2.97E-02 |
| rs820071 | G | T | 0.0121 | 0.0018 | 1.40E-11 | 6.65E-05 | 45.2 |  | 0.0918 | 0.0392 | 1.98E-02 |
| rs9367368 | T | C | 0.0121 | 0.0018 | 1.00E-11 | 6.19E-05 | 45.2 |  | 0.0208 | 0.0398 | 6.03E-01 |
| rs1285245 | G | C | 0.0121 | 0.0018 | 1.70E-11 | 6.80E-05 | 45.2 |  | -0.0361 | 0.0384 | 3.50E-01 |
| rs2009416 | C | T | 0.0121 | 0.0018 | 1.10E-11 | 6.75E-05 | 45.2 |  | 0.0103 | 0.0388 | 7.91E-01 |
| rs962796 | T | C | 0.0148 | 0.0022 | 5.90E-12 | 7.14E-05 | 45.3 |  | 0.0371 | 0.0464 | 4.26E-01 |
| rs1982441 | T | G | 0.0175 | 0.0026 | 7.00E-12 | 7.29E-05 | 45.3 |  | 0.0150 | 0.0515 | 7.71E-01 |
| rs12776880 | A | T | 0.0128 | 0.0019 | 3.00E-11 | 7.10E-05 | 45.4 |  | 0.0285 | 0.0404 | 4.84E-01 |
| rs2731277 | C | T | 0.0135 | 0.002 | 8.80E-12 | 6.90E-05 | 45.6 |  | -0.0642 | 0.0430 | 1.37E-01 |
| rs4717623 | C | T | 0.0135 | 0.002 | 4.20E-12 | 6.93E-05 | 45.6 |  | -0.0434 | 0.0414 | 2.97E-01 |
| rs1199334 | A | G | 0.0142 | 0.0021 | 7.30E-12 | 6.15E-05 | 45.7 |  | 0.0027 | 0.0481 | 9.56E-01 |
| rs4307239 | G | A | 0.0115 | 0.0017 | 3.90E-11 | 6.57E-05 | 45.8 |  | 0.0008 | 0.0378 | 9.83E-01 |
| rs9951893 | C | T | 0.0115 | 0.0017 | 3.30E-11 | 6.60E-05 | 45.8 |  | -0.0146 | 0.0374 | 6.98E-01 |
| rs6545709 | G | A | 0.0203 | 0.003 | 1.50E-11 | 7.35E-05 | 45.8 |  | -0.0586 | 0.0621 | 3.48E-01 |
| rs2051559 | C | T | 0.0176 | 0.0026 | 5.00E-12 | 7.04E-05 | 45.8 |  | 0.0222 | 0.0548 | 6.87E-01 |
| rs6014523 | T | C | 0.0149 | 0.0022 | 1.40E-11 | 7.02E-05 | 45.9 |  | 0.0160 | 0.0480 | 7.40E-01 |
| rs10883759 | G | A | 0.0122 | 0.0018 | 5.40E-12 | 6.28E-05 | 45.9 |  | 0.0849 | 0.0410 | 3.98E-02 |
| rs4653017 | T | C | 0.0122 | 0.0018 | 4.50E-11 | 6.46E-05 | 45.9 |  | -0.0384 | 0.0401 | 3.41E-01 |
| rs10824218 | A | T | 0.0122 | 0.0018 | 7.20E-12 | 7.36E-05 | 45.9 |  | 0.0054 | 0.0371 | 8.84E-01 |
| rs9814633 | A | G | 0.0122 | 0.0018 | 2.10E-11 | 6.71E-05 | 45.9 |  | -0.0437 | 0.0391 | 2.66E-01 |
| rs7760082 | G | A | 0.0122 | 0.0018 | 2.90E-11 | 6.64E-05 | 45.9 |  | -0.0179 | 0.0396 | 6.53E-01 |
| rs4082793 | C | T | 0.0122 | 0.0018 | 3.60E-12 | 7.26E-05 | 45.9 |  | 0.0300 | 0.0377 | 4.30E-01 |
| rs7925748 | G | A | 0.0122 | 0.0018 | 3.30E-12 | 7.16E-05 | 45.9 |  | -0.0034 | 0.0379 | 9.30E-01 |
| rs2467594 | G | A | 0.0122 | 0.0018 | 6.50E-12 | 6.89E-05 | 45.9 |  | 0.0543 | 0.0386 | 1.62E-01 |
| rs10975933 | C | G | 0.0122 | 0.0018 | 2.70E-11 | 6.72E-05 | 45.9 |  | -0.0216 | 0.0393 | 5.86E-01 |
| rs6676084 | C | T | 0.0122 | 0.0018 | 4.90E-12 | 6.36E-05 | 45.9 |  | 0.0099 | 0.0404 | 8.07E-01 |
| rs4858193 | T | C | 0.0129 | 0.0019 | 1.60E-11 | 6.68E-05 | 46.1 |  | -0.0133 | 0.0405 | 7.43E-01 |
| rs16846136 | A | C | 0.0129 | 0.0019 | 3.20E-11 | 6.52E-05 | 46.1 |  | 0.0179 | 0.0416 | 6.68E-01 |
| rs12888545 | G | A | 0.0136 | 0.002 | 9.10E-12 | 6.97E-05 | 46.2 |  | 0.0621 | 0.0423 | 1.45E-01 |
| rs12922346 | C | G | 0.0136 | 0.002 | 1.00E-11 | 7.22E-05 | 46.2 |  | 0.0499 | 0.0417 | 2.34E-01 |
| rs849135 | A | G | 0.0109 | 0.0016 | 2.00E-11 | 5.94E-05 | 46.4 |  | -0.0775 | 0.0372 | 3.81E-02 |
| rs6011457 | T | A | 0.0116 | 0.0017 | 2.70E-11 | 6.73E-05 | 46.6 |  | 0.0714 | 0.0371 | 5.59E-02 |
| rs12476772 | A | C | 0.0123 | 0.0018 | 8.30E-12 | 6.72E-05 | 46.7 |  | 0.0751 | 0.0394 | 5.81E-02 |
| rs10840606 | G | A | 0.0164 | 0.0024 | 3.20E-12 | 7.73E-05 | 46.7 |  | -0.2034 | 0.0487 | 3.33E-05 |
| rs6713781 | G | C | 0.0123 | 0.0018 | 3.80E-12 | 7.34E-05 | 46.7 |  | -0.0060 | 0.0375 | 8.73E-01 |
| rs7844647 | T | C | 0.0123 | 0.0018 | 2.80E-11 | 5.94E-05 | 46.7 |  | 0.0073 | 0.0424 | 8.64E-01 |
| rs7578575 | A | T | 0.013 | 0.0019 | 1.90E-11 | 7.06E-05 | 46.8 |  | 0.0080 | 0.0417 | 8.50E-01 |
| rs17789218 | C | T | 0.013 | 0.0019 | 7.40E-12 | 6.15E-05 | 46.8 |  | 0.0078 | 0.0443 | 8.60E-01 |
| rs934515 | A | G | 0.0185 | 0.0027 | 7.60E-12 | 7.11E-05 | 46.9 |  | -0.0149 | 0.0593 | 8.02E-01 |
| rs3930349 | C | A | 0.0144 | 0.0021 | 4.20E-12 | 7.11E-05 | 47.0 |  | 0.0139 | 0.0452 | 7.59E-01 |
| rs13063194 | C | T | 0.0144 | 0.0021 | 1.10E-11 | 6.79E-05 | 47.0 |  | 0.0504 | 0.0477 | 2.94E-01 |
| rs2850969 | C | T | 0.0165 | 0.0024 | 1.30E-11 | 6.66E-05 | 47.3 |  | -0.1429 | 0.0573 | 1.31E-02 |
| rs17014375 | G | T | 0.0172 | 0.0025 | 1.10E-11 | 6.90E-05 | 47.3 |  | -0.0880 | 0.0546 | 1.09E-01 |
| rs1891215 | C | T | 0.0117 | 0.0017 | 1.30E-11 | 6.79E-05 | 47.4 |  | -0.0054 | 0.0376 | 8.87E-01 |
| rs13110266 | G | A | 0.0117 | 0.0017 | 1.90E-12 | 6.61E-05 | 47.4 |  | 0.0130 | 0.0384 | 7.37E-01 |
| rs7172627 | G | A | 0.0117 | 0.0017 | 1.10E-11 | 6.82E-05 | 47.4 |  | 0.0641 | 0.0372 | 8.67E-02 |
| rs1964927 | G | A | 0.0124 | 0.0018 | 5.00E-12 | 7.10E-05 | 47.5 |  | -0.0509 | 0.0379 | 1.82E-01 |
| rs263041 | A | G | 0.0124 | 0.0018 | 2.20E-12 | 7.14E-05 | 47.5 |  | 0.0065 | 0.0386 | 8.68E-01 |
| rs11635675 | T | G | 0.0124 | 0.0018 | 1.30E-11 | 7.03E-05 | 47.5 |  | -0.0040 | 0.0389 | 9.19E-01 |
| rs7024334 | T | G | 0.0138 | 0.002 | 3.10E-12 | 6.66E-05 | 47.6 |  | 0.0708 | 0.0443 | 1.12E-01 |
| rs7083450 | T | C | 0.0159 | 0.0023 | 1.70E-12 | 6.84E-05 | 47.8 |  | 0.0054 | 0.0502 | 9.15E-01 |
| rs1409818 | T | C | 0.0201 | 0.0029 | 2.50E-12 | 8.26E-05 | 48.0 |  | -0.0372 | 0.0596 | 5.35E-01 |
| rs936227 | G | A | 0.0118 | 0.0017 | 2.30E-12 | 6.62E-05 | 48.2 |  | -0.0272 | 0.0384 | 4.80E-01 |
| rs1730859 | G | A | 0.0118 | 0.0017 | 1.10E-11 | 6.26E-05 | 48.2 |  | 0.0591 | 0.0400 | 1.41E-01 |
| rs4880341 | C | T | 0.0118 | 0.0017 | 1.10E-11 | 6.86E-05 | 48.2 |  | 0.0317 | 0.0374 | 3.99E-01 |
| rs1421334 | A | C | 0.0125 | 0.0018 | 1.00E-12 | 7.75E-05 | 48.2 |  | 0.0846 | 0.0374 | 2.47E-02 |
| rs7133378 | A | G | 0.0125 | 0.0018 | 9.40E-13 | 6.88E-05 | 48.2 |  | -0.0603 | 0.0404 | 1.38E-01 |
| rs33436 | G | A | 0.0125 | 0.0018 | 3.00E-12 | 7.29E-05 | 48.2 |  | -0.0883 | 0.0390 | 2.46E-02 |
| rs7206608 | G | C | 0.0132 | 0.0019 | 1.30E-12 | 7.51E-05 | 48.3 |  | -0.0269 | 0.0397 | 5.01E-01 |
| rs1035010 | T | C | 0.0139 | 0.002 | 6.20E-12 | 7.37E-05 | 48.3 |  | -0.0056 | 0.0430 | 8.97E-01 |
| rs2875762 | C | G | 0.0139 | 0.002 | 1.20E-11 | 7.19E-05 | 48.3 |  | -0.0131 | 0.0448 | 7.71E-01 |
| rs11736228 | A | T | 0.0139 | 0.002 | 4.10E-12 | 7.41E-05 | 48.3 |  | 0.0197 | 0.0429 | 6.49E-01 |
| rs12035149 | G | C | 0.0146 | 0.0021 | 3.90E-12 | 7.39E-05 | 48.3 |  | 0.0018 | 0.0448 | 9.67E-01 |
| rs10510999 | T | C | 0.0146 | 0.0021 | 7.80E-12 | 6.68E-05 | 48.3 |  | 0.0261 | 0.0459 | 5.71E-01 |
| rs6419734 | T | C | 0.0174 | 0.0025 | 1.90E-12 | 7.81E-05 | 48.4 |  | 0.0188 | 0.0523 | 7.20E-01 |
| rs3887080 | A | G | 0.0181 | 0.0026 | 5.10E-12 | 6.99E-05 | 48.5 |  | -0.0470 | 0.0571 | 4.13E-01 |
| rs12325419 | G | A | 0.0188 | 0.0027 | 1.40E-12 | 7.52E-05 | 48.5 |  | 0.0147 | 0.0561 | 7.94E-01 |
| rs16907751 | C | T | 0.0209 | 0.003 | 1.60E-12 | 8.19E-05 | 48.5 |  | 0.0394 | 0.0638 | 5.39E-01 |
| rs4740383 | A | G | 0.0126 | 0.0018 | 1.90E-12 | 7.71E-05 | 49.0 |  | -0.0351 | 0.0376 | 3.53E-01 |
| rs17531363 | A | C | 0.0133 | 0.0019 | 2.00E-12 | 7.48E-05 | 49.0 |  | -0.0647 | 0.0408 | 1.15E-01 |
| rs845084 | A | G | 0.014 | 0.002 | 1.30E-12 | 7.69E-05 | 49.0 |  | -0.0301 | 0.0425 | 4.81E-01 |
| rs6690764 | G | A | 0.0154 | 0.0022 | 6.30E-12 | 7.82E-05 | 49.0 |  | 0.0295 | 0.0453 | 5.18E-01 |
| rs1512914 | G | T | 0.0126 | 0.0018 | 8.10E-13 | 7.55E-05 | 49.0 |  | -0.0257 | 0.0379 | 4.99E-01 |
| rs7607351 | T | C | 0.0119 | 0.0017 | 8.40E-12 | 6.90E-05 | 49.0 |  | -0.0191 | 0.0375 | 6.12E-01 |
| rs16932761 | G | A | 0.014 | 0.002 | 1.70E-12 | 7.43E-05 | 49.0 |  | -0.0084 | 0.0433 | 8.47E-01 |
| rs2029331 | G | C | 0.014 | 0.002 | 2.00E-12 | 7.25E-05 | 49.0 |  | 0.0727 | 0.0440 | 1.00E-01 |
| rs1814170 | A | T | 0.0203 | 0.0029 | 2.10E-12 | 7.77E-05 | 49.0 |  | -0.0472 | 0.0590 | 4.26E-01 |
| rs11614340 | C | T | 0.0133 | 0.0019 | 8.50E-13 | 7.56E-05 | 49.0 |  | -0.0618 | 0.0391 | 1.17E-01 |
| rs10433609 | A | T | 0.0162 | 0.0023 | 1.70E-12 | 7.35E-05 | 49.6 |  | -0.0481 | 0.0486 | 3.25E-01 |
| rs6443750 | C | T | 0.0148 | 0.0021 | 3.20E-12 | 6.83E-05 | 49.7 |  | 0.0002 | 0.0480 | 9.97E-01 |
| rs7186893 | G | T | 0.0141 | 0.002 | 6.50E-13 | 7.81E-05 | 49.7 |  | 0.0779 | 0.0420 | 6.54E-02 |
| rs4718966 | T | C | 0.0127 | 0.0018 | 4.60E-13 | 7.85E-05 | 49.8 |  | -0.0047 | 0.0379 | 9.02E-01 |
| rs6449532 | C | T | 0.0127 | 0.0018 | 1.20E-12 | 7.38E-05 | 49.8 |  | -0.0028 | 0.0391 | 9.44E-01 |
| rs1263618 | C | T | 0.012 | 0.0017 | 5.30E-12 | 6.21E-05 | 49.8 |  | -0.0413 | 0.0399 | 3.04E-01 |
| rs11066188 | G | A | 0.012 | 0.0017 | 8.10E-13 | 7.01E-05 | 49.8 |  | -0.2250 | 0.0377 | 2.94E-09 |
| rs1007934 | G | A | 0.012 | 0.0017 | 7.60E-12 | 6.99E-05 | 49.8 |  | 0.0078 | 0.0379 | 8.38E-01 |
| rs4985155 | A | G | 0.012 | 0.0017 | 3.00E-12 | 6.43E-05 | 49.8 |  | -0.0286 | 0.0398 | 4.74E-01 |
| rs3766430 | C | T | 0.0113 | 0.0016 | 3.90E-12 | 6.30E-05 | 49.9 |  | 0.0246 | 0.0376 | 5.15E-01 |
| rs16966801 | G | A | 0.0156 | 0.0022 | 1.10E-12 | 7.83E-05 | 50.3 |  | 0.0370 | 0.0476 | 4.39E-01 |
| rs6512302 | C | G | 0.0142 | 0.002 | 2.10E-12 | 7.54E-05 | 50.4 |  | -0.0105 | 0.0424 | 8.06E-01 |
| rs7421089 | T | C | 0.0135 | 0.0019 | 7.50E-13 | 7.49E-05 | 50.5 |  | 0.0386 | 0.0408 | 3.47E-01 |
| rs6504165 | T | C | 0.0135 | 0.0019 | 4.00E-13 | 7.06E-05 | 50.5 |  | -0.0174 | 0.0426 | 6.84E-01 |
| rs10971721 | C | T | 0.0199 | 0.0028 | 5.70E-13 | 7.69E-05 | 50.5 |  | 0.0885 | 0.0553 | 1.11E-01 |
| rs1707322 | G | A | 0.0128 | 0.0018 | 4.90E-13 | 6.80E-05 | 50.6 |  | -0.0671 | 0.0405 | 9.97E-02 |
| rs9530843 | A | C | 0.0128 | 0.0018 | 4.80E-13 | 8.09E-05 | 50.6 |  | 0.0190 | 0.0375 | 6.14E-01 |
| rs3811514 | T | C | 0.0121 | 0.0017 | 1.20E-12 | 7.32E-05 | 50.7 |  | 0.0431 | 0.0374 | 2.52E-01 |
| rs765875 | C | T | 0.0121 | 0.0017 | 3.00E-12 | 7.31E-05 | 50.7 |  | -0.0025 | 0.0374 | 9.48E-01 |
| rs2174307 | C | G | 0.0121 | 0.0017 | 4.90E-12 | 7.07E-05 | 50.7 |  | -0.0073 | 0.0377 | 8.47E-01 |
| rs1521527 | G | C | 0.0121 | 0.0017 | 3.10E-12 | 7.29E-05 | 50.7 |  | -0.0265 | 0.0374 | 4.82E-01 |
| rs6595205 | C | G | 0.0114 | 0.0016 | 2.00E-12 | 6.47E-05 | 50.8 |  | 0.0449 | 0.0373 | 2.32E-01 |
| rs4677812 | C | A | 0.0136 | 0.0019 | 1.40E-12 | 7.47E-05 | 51.2 |  | -0.0330 | 0.0414 | 4.28E-01 |
| rs326889 | C | T | 0.0129 | 0.0018 | 2.40E-13 | 7.94E-05 | 51.4 |  | 0.0628 | 0.0378 | 9.89E-02 |
| rs9571687 | C | A | 0.0129 | 0.0018 | 2.80E-12 | 7.35E-05 | 51.4 |  | 0.0302 | 0.0398 | 4.50E-01 |
| rs4936175 | C | T | 0.0122 | 0.0017 | 1.40E-12 | 7.35E-05 | 51.5 |  | -0.0554 | 0.0379 | 1.47E-01 |
| rs7313220 | A | G | 0.0122 | 0.0017 | 1.40E-12 | 7.44E-05 | 51.5 |  | 0.0351 | 0.0372 | 3.48E-01 |
| rs9349239 | A | G | 0.0122 | 0.0017 | 1.30E-12 | 7.44E-05 | 51.5 |  | -0.0096 | 0.0370 | 7.97E-01 |
| rs1955540 | C | T | 0.0158 | 0.0022 | 6.20E-13 | 7.75E-05 | 51.6 |  | -0.0108 | 0.0471 | 8.20E-01 |
| rs2246012 | C | T | 0.0158 | 0.0022 | 3.10E-13 | 6.80E-05 | 51.6 |  | 0.0625 | 0.0504 | 2.18E-01 |
| rs3845802 | G | T | 0.0115 | 0.0016 | 1.20E-12 | 6.61E-05 | 51.7 |  | 0.0008 | 0.0372 | 9.82E-01 |
| rs4722672 | C | T | 0.0151 | 0.0021 | 1.80E-12 | 6.87E-05 | 51.7 |  | 0.0284 | 0.0469 | 5.47E-01 |
| rs11792311 | G | A | 0.0144 | 0.002 | 1.30E-12 | 7.54E-05 | 51.8 |  | 0.0070 | 0.0432 | 8.72E-01 |
| rs538579 | C | G | 0.0137 | 0.0019 | 1.30E-13 | 8.21E-05 | 52.0 |  | 0.0525 | 0.0406 | 1.99E-01 |
| rs1365466 | C | T | 0.0137 | 0.0019 | 3.30E-13 | 7.21E-05 | 52.0 |  | -0.0287 | 0.0430 | 5.07E-01 |
| rs12989476 | T | C | 0.013 | 0.0018 | 5.00E-13 | 7.71E-05 | 52.2 |  | -0.0198 | 0.0392 | 6.15E-01 |
| rs1625427 | T | C | 0.013 | 0.0018 | 1.40E-12 | 7.69E-05 | 52.2 |  | -0.0471 | 0.0393 | 2.34E-01 |
| rs954018 | G | A | 0.013 | 0.0018 | 2.10E-13 | 7.17E-05 | 52.2 |  | -0.0815 | 0.0402 | 4.38E-02 |
| rs4963120 | T | C | 0.013 | 0.0018 | 3.60E-13 | 8.31E-05 | 52.2 |  | -0.0017 | 0.0378 | 9.64E-01 |
| rs945211 | C | G | 0.013 | 0.0018 | 4.50E-13 | 7.82E-05 | 52.2 |  | 0.0345 | 0.0392 | 3.82E-01 |
| rs4148155 | A | G | 0.0188 | 0.0026 | 5.00E-13 | 7.07E-05 | 52.3 |  | 0.0591 | 0.0604 | 3.30E-01 |
| rs2143253 | G | A | 0.0188 | 0.0026 | 1.10E-12 | 7.41E-05 | 52.3 |  | 0.0564 | 0.0606 | 3.54E-01 |
| rs11951673 | C | T | 0.0123 | 0.0017 | 1.10E-13 | 7.23E-05 | 52.3 |  | -0.0072 | 0.0383 | 8.51E-01 |
| rs1941697 | A | G | 0.0123 | 0.0017 | 1.20E-12 | 7.50E-05 | 52.3 |  | 0.0226 | 0.0370 | 5.43E-01 |
| rs6477694 | C | T | 0.0123 | 0.0017 | 3.80E-13 | 6.94E-05 | 52.3 |  | -0.0402 | 0.0383 | 2.98E-01 |
| rs2124499 | G | C | 0.0123 | 0.0017 | 3.40E-13 | 7.07E-05 | 52.3 |  | -0.0098 | 0.0382 | 7.99E-01 |
| rs2467210 | G | A | 0.0145 | 0.002 | 7.90E-13 | 7.38E-05 | 52.6 |  | -0.0604 | 0.0433 | 1.65E-01 |
| rs4969387 | G | C | 0.0145 | 0.002 | 4.30E-13 | 7.96E-05 | 52.6 |  | -0.0294 | 0.0427 | 4.93E-01 |
| rs11855853 | C | T | 0.0145 | 0.002 | 2.40E-13 | 8.19E-05 | 52.6 |  | 0.0074 | 0.0421 | 8.61E-01 |
| rs740157 | A | G | 0.0116 | 0.0016 | 1.80E-12 | 6.60E-05 | 52.6 |  | -0.0182 | 0.0383 | 6.36E-01 |
| rs6448587 | A | C | 0.0167 | 0.0023 | 2.30E-13 | 8.55E-05 | 52.7 |  | 0.0583 | 0.0471 | 2.18E-01 |
| rs9077 | G | A | 0.0138 | 0.0019 | 3.50E-13 | 8.42E-05 | 52.8 |  | 0.0051 | 0.0393 | 8.98E-01 |
| rs2235564 | T | C | 0.0131 | 0.0018 | 3.70E-13 | 7.77E-05 | 53.0 |  | 0.0612 | 0.0393 | 1.22E-01 |
| rs3852012 | G | A | 0.0131 | 0.0018 | 1.40E-13 | 7.37E-05 | 53.0 |  | 0.0220 | 0.0398 | 5.82E-01 |
| rs7239114 | A | G | 0.0124 | 0.0017 | 1.20E-13 | 7.64E-05 | 53.2 |  | -0.0012 | 0.0375 | 9.74E-01 |
| rs340025 | C | T | 0.0124 | 0.0017 | 1.00E-13 | 7.53E-05 | 53.2 |  | 0.0080 | 0.0381 | 8.35E-01 |
| rs10797115 | T | C | 0.0124 | 0.0017 | 9.90E-13 | 7.65E-05 | 53.2 |  | 0.0304 | 0.0373 | 4.18E-01 |
| rs11629783 | C | G | 0.0146 | 0.002 | 9.80E-13 | 7.57E-05 | 53.3 |  | -0.0722 | 0.0443 | 1.05E-01 |
| rs12041258 | T | C | 0.0146 | 0.002 | 9.50E-13 | 7.52E-05 | 53.3 |  | 0.0585 | 0.0437 | 1.83E-01 |
| rs7630302 | G | C | 0.0212 | 0.0029 | 5.60E-13 | 8.07E-05 | 53.4 |  | 0.0927 | 0.0637 | 1.48E-01 |
| rs1899689 | T | C | 0.0117 | 0.0016 | 1.50E-12 | 6.56E-05 | 53.5 |  | -0.0226 | 0.0379 | 5.53E-01 |
| rs7607369 | A | G | 0.0117 | 0.0016 | 9.30E-13 | 6.74E-05 | 53.5 |  | -0.0087 | 0.0380 | 8.19E-01 |
| rs1038088 | G | T | 0.0117 | 0.0016 | 4.60E-13 | 6.84E-05 | 53.5 |  | -0.0281 | 0.0371 | 4.52E-01 |
| rs1150659 | G | A | 0.0139 | 0.0019 | 3.80E-13 | 6.75E-05 | 53.5 |  | -0.0007 | 0.0455 | 9.88E-01 |
| rs10795422 | G | A | 0.0139 | 0.0019 | 9.30E-14 | 8.26E-05 | 53.5 |  | 0.0533 | 0.0405 | 1.91E-01 |
| rs2682406 | T | A | 0.0132 | 0.0018 | 8.90E-14 | 8.47E-05 | 53.8 |  | -0.0388 | 0.0374 | 3.03E-01 |
| rs1784460 | A | T | 0.0132 | 0.0018 | 9.00E-14 | 8.39E-05 | 53.8 |  | 0.1013 | 0.0382 | 8.44E-03 |
| rs1330052 | G | C | 0.0132 | 0.0018 | 1.50E-13 | 7.93E-05 | 53.8 |  | 0.0204 | 0.0391 | 6.05E-01 |
| rs6138482 | T | C | 0.0147 | 0.002 | 5.80E-13 | 6.85E-05 | 54.0 |  | -0.0269 | 0.0456 | 5.57E-01 |
| rs16871902 | A | G | 0.0125 | 0.0017 | 4.60E-13 | 7.81E-05 | 54.1 |  | -0.0180 | 0.0374 | 6.33E-01 |
| rs10832778 | G | C | 0.0125 | 0.0017 | 1.30E-13 | 7.35E-05 | 54.1 |  | 0.0230 | 0.0379 | 5.46E-01 |
| rs329122 | G | A | 0.0125 | 0.0017 | 4.30E-14 | 7.63E-05 | 54.1 |  | -0.0147 | 0.0374 | 6.96E-01 |
| rs7519259 | A | G | 0.0125 | 0.0017 | 3.80E-13 | 7.77E-05 | 54.1 |  | -0.0162 | 0.0370 | 6.63E-01 |
| rs6864049 | G | A | 0.0125 | 0.0017 | 6.70E-14 | 7.76E-05 | 54.1 |  | 0.0175 | 0.0374 | 6.42E-01 |
| rs1358980 | C | T | 0.0125 | 0.0017 | 1.10E-13 | 7.80E-05 | 54.1 |  | 0.0163 | 0.0376 | 6.67E-01 |
| rs12439798 | T | G | 0.0125 | 0.0017 | 6.90E-13 | 7.63E-05 | 54.1 |  | 0.0344 | 0.0379 | 3.67E-01 |
| rs977540 | A | G | 0.014 | 0.0019 | 2.50E-13 | 7.12E-05 | 54.3 |  | -0.0217 | 0.0433 | 6.18E-01 |
| rs7869771 | A | C | 0.014 | 0.0019 | 4.90E-13 | 7.63E-05 | 54.3 |  | 0.0252 | 0.0443 | 5.72E-01 |
| rs10838465 | A | C | 0.014 | 0.0019 | 9.10E-14 | 8.28E-05 | 54.3 |  | 0.0623 | 0.0403 | 1.24E-01 |
| rs12147845 | T | C | 0.0199 | 0.0027 | 4.50E-13 | 7.97E-05 | 54.3 |  | -0.0623 | 0.0590 | 2.94E-01 |
| rs10923724 | C | T | 0.0118 | 0.0016 | 6.40E-13 | 6.82E-05 | 54.4 |  | -0.0047 | 0.0376 | 9.02E-01 |
| rs1689437 | G | A | 0.0251 | 0.0034 | 2.10E-13 | 7.82E-05 | 54.5 |  | 0.0059 | 0.0815 | 9.43E-01 |
| rs10007906 | A | C | 0.0133 | 0.0018 | 1.70E-13 | 8.15E-05 | 54.6 |  | -0.0056 | 0.0387 | 8.86E-01 |
| rs6738445 | C | T | 0.0133 | 0.0018 | 1.90E-13 | 7.19E-05 | 54.6 |  | -0.0220 | 0.0414 | 5.98E-01 |
| rs11150911 | A | C | 0.0133 | 0.0018 | 4.70E-13 | 7.15E-05 | 54.6 |  | 0.0260 | 0.0403 | 5.21E-01 |
| rs12680842 | A | G | 0.0133 | 0.0018 | 4.40E-14 | 7.70E-05 | 54.6 |  | -0.0242 | 0.0395 | 5.43E-01 |
| rs2890652 | C | T | 0.017 | 0.0023 | 2.50E-13 | 8.38E-05 | 54.6 |  | 0.0963 | 0.0479 | 4.57E-02 |
| rs9267677 | C | T | 0.0207 | 0.0028 | 1.60E-13 | 7.79E-05 | 54.7 |  | -0.0876 | 0.0722 | 2.28E-01 |
| rs11945861 | G | A | 0.0148 | 0.002 | 5.00E-13 | 7.92E-05 | 54.8 |  | -0.0243 | 0.0450 | 5.91E-01 |
| rs10818810 | A | G | 0.0126 | 0.0017 | 1.10E-13 | 7.56E-05 | 54.9 |  | 0.0327 | 0.0381 | 3.93E-01 |
| rs10878946 | C | T | 0.0141 | 0.0019 | 3.60E-13 | 8.12E-05 | 55.1 |  | 0.1049 | 0.0408 | 1.07E-02 |
| rs7235205 | G | A | 0.0141 | 0.0019 | 4.40E-14 | 7.76E-05 | 55.1 |  | -0.0182 | 0.0411 | 6.60E-01 |
| rs2907948 | G | A | 0.0141 | 0.0019 | 1.30E-13 | 7.31E-05 | 55.1 |  | -0.0343 | 0.0444 | 4.43E-01 |
| rs2190788 | T | G | 0.0141 | 0.0019 | 2.90E-14 | 8.51E-05 | 55.1 |  | -0.0218 | 0.0403 | 5.91E-01 |
| rs6471941 | A | G | 0.0156 | 0.0021 | 3.10E-13 | 6.82E-05 | 55.2 |  | -0.0316 | 0.0486 | 5.17E-01 |
| rs2367112 | T | G | 0.0119 | 0.0016 | 2.30E-13 | 7.08E-05 | 55.3 |  | 0.0228 | 0.0373 | 5.43E-01 |
| rs506338 | C | T | 0.0134 | 0.0018 | 7.80E-14 | 7.52E-05 | 55.4 |  | 0.0060 | 0.0406 | 8.83E-01 |
| rs6445538 | C | T | 0.0149 | 0.002 | 1.40E-13 | 7.99E-05 | 55.5 |  | 0.0634 | 0.0433 | 1.45E-01 |
| rs1522569 | T | G | 0.0164 | 0.0022 | 2.90E-13 | 8.00E-05 | 55.6 |  | -0.0032 | 0.0497 | 9.48E-01 |
| rs6710871 | A | G | 0.0179 | 0.0024 | 1.00E-13 | 7.80E-05 | 55.6 |  | -0.0214 | 0.0560 | 7.04E-01 |
| rs3807566 | G | T | 0.0127 | 0.0017 | 2.00E-13 | 7.96E-05 | 55.8 |  | -0.0262 | 0.0376 | 4.88E-01 |
| rs7874154 | C | T | 0.0127 | 0.0017 | 1.80E-13 | 8.06E-05 | 55.8 |  | 0.0598 | 0.0373 | 1.11E-01 |
| rs559231 | T | G | 0.0135 | 0.0018 | 2.40E-14 | 8.72E-05 | 56.3 |  | 0.0281 | 0.0380 | 4.62E-01 |
| rs769674 | A | T | 0.0135 | 0.0018 | 2.20E-13 | 7.96E-05 | 56.3 |  | 0.0152 | 0.0397 | 7.03E-01 |
| rs7947143 | G | A | 0.018 | 0.0024 | 2.40E-14 | 8.82E-05 | 56.3 |  | 0.0278 | 0.0531 | 6.03E-01 |
| rs2185027 | C | A | 0.0135 | 0.0018 | 5.00E-14 | 7.61E-05 | 56.3 |  | 0.0261 | 0.0404 | 5.21E-01 |
| rs12762034 | C | T | 0.024 | 0.0032 | 7.30E-14 | 8.07E-05 | 56.3 |  | 0.1264 | 0.0688 | 6.77E-02 |
| rs17203016 | G | A | 0.015 | 0.002 | 2.10E-13 | 7.09E-05 | 56.3 |  | 0.0017 | 0.0458 | 9.71E-01 |
| rs2289379 | C | T | 0.0135 | 0.0018 | 4.30E-14 | 8.73E-05 | 56.3 |  | -0.0530 | 0.0380 | 1.65E-01 |
| rs6707445 | A | G | 0.0128 | 0.0017 | 2.10E-13 | 8.12E-05 | 56.7 |  | -0.0586 | 0.0376 | 1.21E-01 |
| rs10887578 | C | G | 0.0128 | 0.0017 | 1.60E-13 | 8.19E-05 | 56.7 |  | 0.0548 | 0.0373 | 1.44E-01 |
| rs349088 | C | A | 0.0128 | 0.0017 | 1.80E-13 | 8.19E-05 | 56.7 |  | -0.0333 | 0.0375 | 3.76E-01 |
| rs12868881 | A | T | 0.0128 | 0.0017 | 1.70E-13 | 7.98E-05 | 56.7 |  | 0.0395 | 0.0376 | 2.96E-01 |
| rs12564992 | G | A | 0.0196 | 0.0026 | 5.30E-14 | 7.78E-05 | 56.8 |  | -0.0639 | 0.0595 | 2.85E-01 |
| rs10192119 | G | T | 0.0166 | 0.0022 | 3.00E-14 | 7.68E-05 | 56.9 |  | -0.0355 | 0.0493 | 4.74E-01 |
| rs6548221 | A | G | 0.0151 | 0.002 | 1.70E-14 | 7.94E-05 | 57.0 |  | 0.0195 | 0.0460 | 6.73E-01 |
| rs6548834 | A | G | 0.0136 | 0.0018 | 1.80E-14 | 8.55E-05 | 57.1 |  | 0.0332 | 0.0386 | 3.93E-01 |
| rs7640424 | C | T | 0.0136 | 0.0018 | 2.30E-14 | 7.72E-05 | 57.1 |  | 0.0253 | 0.0408 | 5.37E-01 |
| rs6606686 | G | C | 0.0136 | 0.0018 | 7.60E-15 | 7.91E-05 | 57.1 |  | -0.0203 | 0.0407 | 6.21E-01 |
| rs1075901 | C | T | 0.0121 | 0.0016 | 1.20E-13 | 7.20E-05 | 57.2 |  | 0.0161 | 0.0374 | 6.68E-01 |
| rs2516739 | G | A | 0.0159 | 0.0021 | 1.40E-14 | 8.59E-05 | 57.3 |  | 0.0210 | 0.0469 | 6.56E-01 |
| rs7561278 | T | C | 0.0159 | 0.0021 | 5.70E-14 | 8.50E-05 | 57.3 |  | 0.0014 | 0.0465 | 9.76E-01 |
| rs6461115 | A | G | 0.0144 | 0.0019 | 1.20E-13 | 7.31E-05 | 57.4 |  | -0.0441 | 0.0433 | 3.12E-01 |
| rs217671 | G | A | 0.0144 | 0.0019 | 1.30E-13 | 8.21E-05 | 57.4 |  | 0.0194 | 0.0423 | 6.48E-01 |
| rs2192158 | A | G | 0.0129 | 0.0017 | 8.30E-14 | 8.27E-05 | 57.6 |  | 0.0068 | 0.0372 | 8.56E-01 |
| rs2080454 | C | A | 0.0129 | 0.0017 | 1.70E-14 | 7.83E-05 | 57.6 |  | -0.0324 | 0.0381 | 3.98E-01 |
| rs1554790 | G | C | 0.0129 | 0.0017 | 6.80E-14 | 8.31E-05 | 57.6 |  | 0.0070 | 0.0377 | 8.53E-01 |
| rs10497810 | C | T | 0.0167 | 0.0022 | 5.90E-14 | 7.98E-05 | 57.6 |  | 0.0779 | 0.0500 | 1.21E-01 |
| rs16867703 | G | T | 0.0137 | 0.0018 | 1.40E-14 | 8.71E-05 | 57.9 |  | -0.0008 | 0.0390 | 9.84E-01 |
| rs11264483 | C | G | 0.0137 | 0.0018 | 3.00E-14 | 8.91E-05 | 57.9 |  | 0.0241 | 0.0382 | 5.32E-01 |
| rs1158805 | C | A | 0.0137 | 0.0018 | 1.20E-14 | 8.81E-05 | 57.9 |  | 0.0020 | 0.0380 | 9.58E-01 |
| rs2246664 | A | G | 0.0137 | 0.0018 | 1.80E-14 | 8.71E-05 | 57.9 |  | 0.0129 | 0.0389 | 7.42E-01 |
| rs1951455 | C | T | 0.0145 | 0.0019 | 4.50E-14 | 8.38E-05 | 58.2 |  | 0.0093 | 0.0415 | 8.24E-01 |
| rs1876359 | T | C | 0.013 | 0.0017 | 1.50E-14 | 7.79E-05 | 58.5 |  | -0.0178 | 0.0395 | 6.53E-01 |
| rs7217226 | G | T | 0.013 | 0.0017 | 2.60E-14 | 7.78E-05 | 58.5 |  | -0.0249 | 0.0388 | 5.24E-01 |
| rs733594 | T | C | 0.0138 | 0.0018 | 5.90E-14 | 7.71E-05 | 58.8 |  | -0.0073 | 0.0416 | 8.61E-01 |
| rs10510419 | G | T | 0.0177 | 0.0023 | 2.20E-14 | 7.62E-05 | 59.2 |  | -0.0164 | 0.0535 | 7.61E-01 |
| rs9650755 | G | A | 0.0154 | 0.002 | 2.80E-15 | 9.27E-05 | 59.3 |  | -0.0187 | 0.0420 | 6.59E-01 |
| rs13292976 | T | C | 0.0131 | 0.0017 | 3.10E-14 | 8.48E-05 | 59.4 |  | 0.0183 | 0.0373 | 6.25E-01 |
| rs2163188 | C | G | 0.0131 | 0.0017 | 2.00E-14 | 8.56E-05 | 59.4 |  | 0.0229 | 0.0368 | 5.37E-01 |
| rs10779751 | A | G | 0.0139 | 0.0018 | 2.50E-14 | 7.69E-05 | 59.6 |  | 0.0305 | 0.0419 | 4.69E-01 |
| rs7780752 | C | T | 0.0139 | 0.0018 | 1.00E-14 | 8.90E-05 | 59.6 |  | -0.0011 | 0.0388 | 9.77E-01 |
| rs284227 | C | T | 0.0147 | 0.0019 | 3.50E-15 | 8.21E-05 | 59.9 |  | 0.0420 | 0.0424 | 3.25E-01 |
| rs6019482 | C | T | 0.0178 | 0.0023 | 2.80E-14 | 8.75E-05 | 59.9 |  | -0.0250 | 0.0484 | 6.07E-01 |
| rs3849570 | A | C | 0.0132 | 0.0017 | 3.30E-14 | 7.84E-05 | 60.3 |  | -0.0755 | 0.0386 | 5.15E-02 |
| rs12705977 | T | G | 0.0132 | 0.0017 | 2.10E-14 | 8.64E-05 | 60.3 |  | 0.0321 | 0.0371 | 3.89E-01 |
| rs998732 | A | G | 0.0171 | 0.0022 | 2.00E-14 | 7.77E-05 | 60.4 |  | -0.0146 | 0.0519 | 7.80E-01 |
| rs13012099 | G | A | 0.014 | 0.0018 | 1.20E-14 | 8.80E-05 | 60.5 |  | -0.0164 | 0.0391 | 6.77E-01 |
| rs1045411 | C | T | 0.0148 | 0.0019 | 2.30E-15 | 8.53E-05 | 60.7 |  | 0.0980 | 0.0421 | 2.06E-02 |
| rs7871866 | C | G | 0.0187 | 0.0024 | 2.30E-14 | 9.07E-05 | 60.7 |  | -0.0196 | 0.0534 | 7.15E-01 |
| rs7536433 | C | T | 0.0156 | 0.002 | 9.80E-15 | 7.96E-05 | 60.8 |  | -0.0626 | 0.0454 | 1.70E-01 |
| rs329651 | T | G | 0.0164 | 0.0021 | 9.00E-15 | 8.43E-05 | 61.0 |  | -0.0427 | 0.0493 | 3.90E-01 |
| rs17327461 | T | C | 0.0125 | 0.0016 | 1.50E-14 | 7.73E-05 | 61.0 |  | 0.0094 | 0.0373 | 8.03E-01 |
| rs10269783 | A | G | 0.0133 | 0.0017 | 1.40E-15 | 8.41E-05 | 61.2 |  | 0.0018 | 0.0378 | 9.63E-01 |
| rs7195386 | T | C | 0.0133 | 0.0017 | 1.10E-14 | 8.84E-05 | 61.2 |  | -0.0154 | 0.0374 | 6.83E-01 |
| rs4012234 | G | T | 0.0141 | 0.0018 | 9.90E-16 | 9.60E-05 | 61.4 |  | 0.0108 | 0.0379 | 7.78E-01 |
| rs1452075 | T | C | 0.0141 | 0.0018 | 1.30E-14 | 7.88E-05 | 61.4 |  | -0.0243 | 0.0422 | 5.66E-01 |
| rs13290794 | G | A | 0.0141 | 0.0018 | 1.90E-15 | 9.14E-05 | 61.4 |  | 0.0582 | 0.0382 | 1.30E-01 |
| rs1336486 | G | T | 0.0141 | 0.0018 | 1.80E-14 | 8.66E-05 | 61.4 |  | -0.0876 | 0.0404 | 3.11E-02 |
| rs7601895 | C | G | 0.0149 | 0.0019 | 1.70E-15 | 9.25E-05 | 61.5 |  | -0.0265 | 0.0417 | 5.28E-01 |
| rs905938 | C | T | 0.0149 | 0.0019 | 1.20E-15 | 8.70E-05 | 61.5 |  | -0.0132 | 0.0414 | 7.52E-01 |
| rs12299814 | C | A | 0.0157 | 0.002 | 5.20E-15 | 9.30E-05 | 61.6 |  | -0.0087 | 0.0419 | 8.37E-01 |
| rs155510 | T | G | 0.0165 | 0.0021 | 1.70E-14 | 8.42E-05 | 61.7 |  | -0.0239 | 0.0457 | 6.03E-01 |
| rs2282231 | T | C | 0.0165 | 0.0021 | 4.80E-15 | 9.49E-05 | 61.7 |  | 0.0491 | 0.0441 | 2.68E-01 |
| rs11889536 | A | G | 0.0189 | 0.0024 | 6.40E-15 | 9.07E-05 | 62.0 |  | 0.0241 | 0.0547 | 6.61E-01 |
| rs10460960 | A | G | 0.0197 | 0.0025 | 8.10E-15 | 7.74E-05 | 62.1 |  | 0.1239 | 0.0562 | 2.85E-02 |
| rs4372836 | T | C | 0.0142 | 0.0018 | 7.30E-16 | 8.52E-05 | 62.2 |  | -0.0101 | 0.0409 | 8.06E-01 |
| rs7573263 | T | C | 0.0142 | 0.0018 | 1.60E-15 | 9.75E-05 | 62.2 |  | 0.0232 | 0.0376 | 5.40E-01 |
| rs33485 | C | T | 0.0158 | 0.002 | 1.10E-15 | 9.82E-05 | 62.4 |  | -0.0366 | 0.0415 | 3.80E-01 |
| rs946824 | T | C | 0.0206 | 0.0026 | 1.10E-15 | 1.03E-04 | 62.8 |  | -0.0342 | 0.0574 | 5.53E-01 |
| rs8088123 | C | A | 0.0262 | 0.0033 | 3.70E-15 | 9.75E-05 | 63.0 |  | -0.0159 | 0.0723 | 8.27E-01 |
| rs17820822 | T | G | 0.0143 | 0.0018 | 2.90E-15 | 9.37E-05 | 63.1 |  | -0.0008 | 0.0383 | 9.84E-01 |
| rs3209570 | G | A | 0.0143 | 0.0018 | 9.80E-16 | 9.58E-05 | 63.1 |  | 0.0930 | 0.0386 | 1.68E-02 |
| rs4711986 | A | G | 0.0143 | 0.0018 | 1.70E-15 | 9.35E-05 | 63.1 |  | -0.0167 | 0.0391 | 6.71E-01 |
| rs8097672 | T | A | 0.02 | 0.0025 | 8.40E-16 | 1.04E-04 | 64.0 |  | -0.0314 | 0.0519 | 5.48E-01 |
| rs2281819 | T | A | 0.016 | 0.002 | 5.10E-15 | 9.07E-05 | 64.0 |  | 0.0942 | 0.0444 | 3.49E-02 |
| rs3957285 | A | G | 0.0144 | 0.0018 | 2.90E-16 | 1.03E-04 | 64.0 |  | 0.0207 | 0.0380 | 5.89E-01 |
| rs10939792 | G | C | 0.0152 | 0.0019 | 1.40E-15 | 1.02E-04 | 64.0 |  | 0.0247 | 0.0396 | 5.36E-01 |
| rs12454712 | C | T | 0.0144 | 0.0018 | 4.40E-16 | 9.67E-05 | 64.0 |  | -0.0556 | 0.0387 | 1.53E-01 |
| rs10886017 | A | C | 0.0152 | 0.0019 | 1.40E-15 | 8.72E-05 | 64.0 |  | -0.0518 | 0.0436 | 2.38E-01 |
| rs13380104 | C | T | 0.0136 | 0.0017 | 7.70E-15 | 9.00E-05 | 64.0 |  | -0.0031 | 0.0379 | 9.35E-01 |
| rs2271189 | G | A | 0.0144 | 0.0018 | 5.00E-16 | 9.99E-05 | 64.0 |  | -0.1384 | 0.0378 | 2.75E-04 |
| rs9827823 | T | C | 0.0193 | 0.0024 | 1.20E-15 | 9.44E-05 | 64.7 |  | 0.0289 | 0.0532 | 5.89E-01 |
| rs10146527 | T | C | 0.0137 | 0.0017 | 2.30E-15 | 8.69E-05 | 64.9 |  | -0.0094 | 0.0390 | 8.10E-01 |
| rs10842240 | C | G | 0.0218 | 0.0027 | 3.10E-16 | 9.73E-05 | 65.2 |  | 0.0454 | 0.0594 | 4.47E-01 |
| rs498240 | G | A | 0.0267 | 0.0033 | 1.60E-15 | 8.74E-05 | 65.5 |  | 0.3216 | 0.0800 | 6.52E-05 |
| rs12364470 | G | T | 0.0178 | 0.0022 | 1.10E-15 | 8.63E-05 | 65.5 |  | 0.0544 | 0.0496 | 2.76E-01 |
| rs12593036 | A | G | 0.0154 | 0.0019 | 3.80E-16 | 9.95E-05 | 65.7 |  | 0.0666 | 0.0401 | 9.88E-02 |
| rs12033257 | A | G | 0.0146 | 0.0018 | 2.40E-15 | 1.01E-04 | 65.8 |  | 0.0178 | 0.0382 | 6.43E-01 |
| rs1895957 | G | T | 0.0171 | 0.0021 | 1.40E-16 | 1.01E-04 | 66.3 |  | -0.0897 | 0.0447 | 4.61E-02 |
| rs7925214 | T | C | 0.0147 | 0.0018 | 4.40E-17 | 1.08E-04 | 66.7 |  | 0.0381 | 0.0372 | 3.09E-01 |
| rs8123881 | G | A | 0.0196 | 0.0024 | 4.40E-16 | 8.68E-05 | 66.7 |  | 0.1338 | 0.0522 | 1.08E-02 |
| rs2228213 | G | A | 0.0139 | 0.0017 | 4.60E-16 | 8.77E-05 | 66.9 |  | 0.0544 | 0.0383 | 1.58E-01 |
| rs9540493 | A | G | 0.0139 | 0.0017 | 8.10E-17 | 9.53E-05 | 66.9 |  | 0.0031 | 0.0375 | 9.33E-01 |
| rs6841761 | G | T | 0.0131 | 0.0016 | 6.40E-16 | 8.56E-05 | 67.0 |  | -0.0451 | 0.0373 | 2.30E-01 |
| rs4556997 | A | C | 0.0197 | 0.0024 | 6.90E-17 | 9.06E-05 | 67.4 |  | -0.0255 | 0.0556 | 6.48E-01 |
| rs10942267 | A | G | 0.0156 | 0.0019 | 3.90E-17 | 1.04E-04 | 67.4 |  | -0.0003 | 0.0404 | 9.94E-01 |
| rs11866815 | C | T | 0.0156 | 0.0019 | 1.00E-16 | 9.03E-05 | 67.4 |  | 0.0259 | 0.0445 | 5.63E-01 |
| rs7730004 | T | C | 0.0148 | 0.0018 | 9.10E-16 | 9.70E-05 | 67.6 |  | -0.0381 | 0.0393 | 3.35E-01 |
| rs536445 | T | C | 0.014 | 0.0017 | 4.60E-16 | 9.77E-05 | 67.8 |  | -0.0224 | 0.0374 | 5.52E-01 |
| rs1503526 | C | T | 0.014 | 0.0017 | 5.50E-17 | 9.79E-05 | 67.8 |  | -0.0667 | 0.0375 | 7.70E-02 |
| rs1436344 | C | G | 0.0141 | 0.0017 | 4.10E-16 | 9.60E-05 | 68.8 |  | -0.0080 | 0.0379 | 8.33E-01 |
| rs811054 | T | C | 0.0141 | 0.0017 | 2.70E-17 | 9.89E-05 | 68.8 |  | -0.0437 | 0.0378 | 2.50E-01 |
| rs4864201 | T | C | 0.0141 | 0.0017 | 1.50E-16 | 9.08E-05 | 68.8 |  | 0.0330 | 0.0389 | 4.00E-01 |
| rs13184896 | G | T | 0.0133 | 0.0016 | 3.30E-16 | 8.69E-05 | 69.1 |  | -0.0146 | 0.0376 | 6.98E-01 |
| rs13174863 | G | A | 0.0192 | 0.0023 | 2.90E-16 | 9.65E-05 | 69.7 |  | -0.0461 | 0.0509 | 3.68E-01 |
| rs6692586 | A | G | 0.0192 | 0.0023 | 1.10E-16 | 1.03E-04 | 69.7 |  | 0.0154 | 0.0502 | 7.60E-01 |
| rs10145461 | G | T | 0.0142 | 0.0017 | 4.70E-16 | 1.00E-04 | 69.8 |  | 0.0041 | 0.0371 | 9.12E-01 |
| rs3007105 | T | C | 0.0142 | 0.0017 | 1.10E-17 | 1.00E-04 | 69.8 |  | 0.0291 | 0.0378 | 4.44E-01 |
| rs4673553 | G | T | 0.0142 | 0.0017 | 2.10E-16 | 1.00E-04 | 69.8 |  | 0.0173 | 0.0376 | 6.47E-01 |
| rs3736485 | A | G | 0.0134 | 0.0016 | 2.50E-16 | 8.91E-05 | 70.1 |  | -0.0495 | 0.0380 | 1.95E-01 |
| rs12602912 | T | C | 0.0176 | 0.0021 | 9.90E-18 | 1.01E-04 | 70.2 |  | -0.0701 | 0.0463 | 1.32E-01 |
| rs17636031 | C | T | 0.016 | 0.0019 | 1.20E-17 | 1.01E-04 | 70.9 |  | 0.0457 | 0.0432 | 2.93E-01 |
| rs29941 | G | A | 0.0152 | 0.0018 | 5.10E-18 | 9.94E-05 | 71.3 |  | -0.0052 | 0.0397 | 8.96E-01 |
| rs13209872 | G | C | 0.0152 | 0.0018 | 1.40E-16 | 1.04E-04 | 71.3 |  | 0.0749 | 0.0396 | 6.01E-02 |
| rs215634 | A | G | 0.0152 | 0.0018 | 2.60E-17 | 1.09E-04 | 71.3 |  | -0.0050 | 0.0386 | 8.97E-01 |
| rs8192675 | C | T | 0.0152 | 0.0018 | 1.40E-17 | 9.49E-05 | 71.3 |  | 0.1046 | 0.0403 | 9.83E-03 |
| rs7535528 | G | A | 0.0152 | 0.0018 | 1.40E-16 | 1.08E-04 | 71.3 |  | -0.0174 | 0.0385 | 6.53E-01 |
| rs995258 | A | C | 0.0144 | 0.0017 | 4.50E-17 | 1.01E-04 | 71.8 |  | -0.0799 | 0.0376 | 3.45E-02 |
| rs13417156 | C | T | 0.0144 | 0.0017 | 2.60E-17 | 1.02E-04 | 71.8 |  | 0.0007 | 0.0375 | 9.84E-01 |
| rs273504 | G | A | 0.0153 | 0.0018 | 4.40E-18 | 1.15E-04 | 72.3 |  | 0.0365 | 0.0379 | 3.38E-01 |
| rs13240600 | A | G | 0.0204 | 0.0024 | 3.50E-17 | 1.09E-04 | 72.3 |  | -0.0753 | 0.0512 | 1.44E-01 |
| rs2357760 | A | G | 0.0145 | 0.0017 | 6.80E-17 | 9.22E-05 | 72.8 |  | 0.0039 | 0.0395 | 9.22E-01 |
| rs12477088 | T | C | 0.0145 | 0.0017 | 8.90E-17 | 1.01E-04 | 72.8 |  | -0.0111 | 0.0381 | 7.72E-01 |
| rs13047416 | C | G | 0.0154 | 0.0018 | 2.20E-17 | 1.11E-04 | 73.2 |  | 0.0046 | 0.0391 | 9.06E-01 |
| rs2479958 | A | G | 0.0154 | 0.0018 | 1.50E-17 | 1.19E-04 | 73.2 |  | 0.0063 | 0.0373 | 8.67E-01 |
| rs13263601 | C | A | 0.0154 | 0.0018 | 2.20E-17 | 1.08E-04 | 73.2 |  | 0.0002 | 0.0398 | 9.97E-01 |
| rs1000940 | G | A | 0.0154 | 0.0018 | 1.10E-17 | 9.94E-05 | 73.2 |  | 0.0119 | 0.0409 | 7.73E-01 |
| rs901630 | C | T | 0.0146 | 0.0017 | 1.90E-18 | 1.02E-04 | 73.8 |  | 0.0077 | 0.0380 | 8.41E-01 |
| rs12454204 | T | G | 0.0172 | 0.002 | 3.80E-17 | 1.05E-04 | 74.0 |  | -0.1048 | 0.0435 | 1.67E-02 |
| rs10962549 | T | C | 0.0198 | 0.0023 | 2.50E-17 | 1.10E-04 | 74.1 |  | 0.0694 | 0.0513 | 1.79E-01 |
| rs2162524 | C | T | 0.0155 | 0.0018 | 4.10E-17 | 1.07E-04 | 74.2 |  | 0.0517 | 0.0393 | 1.91E-01 |
| rs592483 | C | T | 0.0147 | 0.0017 | 2.00E-18 | 1.06E-04 | 74.8 |  | -0.0127 | 0.0371 | 7.34E-01 |
| rs6587552 | A | G | 0.0173 | 0.002 | 1.60E-17 | 1.09E-04 | 74.8 |  | 0.0291 | 0.0432 | 5.02E-01 |
| rs1006896 | A | C | 0.0234 | 0.0027 | 5.50E-18 | 1.04E-04 | 75.1 |  | -0.0470 | 0.0605 | 4.40E-01 |
| rs4500930 | T | C | 0.0156 | 0.0018 | 6.50E-18 | 1.10E-04 | 75.1 |  | 0.0520 | 0.0386 | 1.81E-01 |
| rs1852006 | G | A | 0.0156 | 0.0018 | 4.90E-18 | 1.11E-04 | 75.1 |  | -0.0030 | 0.0391 | 9.39E-01 |
| rs9507983 | C | T | 0.0156 | 0.0018 | 1.40E-18 | 1.16E-04 | 75.1 |  | -0.0171 | 0.0382 | 6.57E-01 |
| rs756717 | G | A | 0.0148 | 0.0017 | 5.40E-18 | 1.05E-04 | 75.8 |  | -0.0026 | 0.0382 | 9.46E-01 |
| rs7025938 | G | C | 0.0166 | 0.0019 | 3.70E-19 | 1.20E-04 | 76.3 |  | -0.0589 | 0.0399 | 1.42E-01 |
| rs10197031 | C | T | 0.0166 | 0.0019 | 1.90E-18 | 1.12E-04 | 76.3 |  | 0.0630 | 0.0408 | 1.25E-01 |
| rs895330 | C | G | 0.0201 | 0.0023 | 5.50E-19 | 1.26E-04 | 76.4 |  | -0.0325 | 0.0464 | 4.85E-01 |
| rs11084553 | A | G | 0.021 | 0.0024 | 1.80E-18 | 1.14E-04 | 76.6 |  | 0.0705 | 0.0551 | 2.04E-01 |
| rs7102454 | C | T | 0.0158 | 0.0018 | 2.40E-18 | 1.13E-04 | 77.0 |  | -0.0396 | 0.0397 | 3.21E-01 |
| rs1928295 | T | C | 0.0141 | 0.0016 | 5.40E-18 | 9.82E-05 | 77.7 |  | 0.0086 | 0.0375 | 8.19E-01 |
| rs429343 | A | G | 0.015 | 0.0017 | 6.80E-18 | 1.10E-04 | 77.9 |  | -0.0144 | 0.0381 | 7.06E-01 |
| rs2478879 | A | G | 0.0159 | 0.0018 | 3.80E-19 | 1.21E-04 | 78.0 |  | -0.0142 | 0.0381 | 7.10E-01 |
| rs4820408 | T | G | 0.0151 | 0.0017 | 2.10E-19 | 1.10E-04 | 78.9 |  | -0.0378 | 0.0382 | 3.25E-01 |
| rs7557796 | T | C | 0.016 | 0.0018 | 2.30E-19 | 1.16E-04 | 79.0 |  | -0.0058 | 0.0395 | 8.85E-01 |
| rs11191548 | C | T | 0.0268 | 0.003 | 5.00E-19 | 1.08E-04 | 79.8 |  | 0.1048 | 0.0618 | 9.19E-02 |
| rs10929925 | C | A | 0.0143 | 0.0016 | 1.80E-18 | 1.00E-04 | 79.9 |  | -0.0338 | 0.0373 | 3.67E-01 |
| rs12044597 | G | A | 0.0143 | 0.0016 | 1.70E-18 | 1.02E-04 | 79.9 |  | -0.0380 | 0.0373 | 3.12E-01 |
| rs2396625 | T | A | 0.0162 | 0.0018 | 3.60E-20 | 1.29E-04 | 81.0 |  | 0.0023 | 0.0378 | 9.51E-01 |
| rs355777 | C | G | 0.0153 | 0.0017 | 1.40E-18 | 1.13E-04 | 81.0 |  | -0.0658 | 0.0379 | 8.41E-02 |
| rs17094222 | C | T | 0.0181 | 0.002 | 2.20E-19 | 1.06E-04 | 81.9 |  | -0.0326 | 0.0468 | 4.89E-01 |
| rs4639527 | G | A | 0.0172 | 0.0019 | 3.30E-20 | 1.25E-04 | 82.0 |  | 0.0591 | 0.0405 | 1.46E-01 |
| rs7181498 | T | C | 0.0163 | 0.0018 | 1.00E-19 | 1.24E-04 | 82.0 |  | 0.0022 | 0.0387 | 9.54E-01 |
| rs930295 | A | C | 0.0211 | 0.0023 | 1.00E-19 | 1.19E-04 | 84.2 |  | 0.0234 | 0.0495 | 6.39E-01 |
| rs6804842 | G | A | 0.0156 | 0.0017 | 3.60E-21 | 1.19E-04 | 84.2 |  | -0.0028 | 0.0378 | 9.40E-01 |
| rs9294260 | A | G | 0.0147 | 0.0016 | 1.80E-19 | 1.08E-04 | 84.4 |  | 0.0352 | 0.0374 | 3.48E-01 |
| rs12448257 | A | G | 0.0184 | 0.002 | 8.10E-20 | 1.15E-04 | 84.6 |  | 0.0166 | 0.0463 | 7.21E-01 |
| rs6235 | G | C | 0.0175 | 0.0019 | 1.50E-19 | 1.21E-04 | 84.8 |  | -0.0942 | 0.0411 | 2.28E-02 |
| rs12630999 | A | G | 0.0175 | 0.0019 | 8.50E-21 | 1.16E-04 | 84.8 |  | 0.0118 | 0.0435 | 7.88E-01 |
| rs2065418 | T | G | 0.0166 | 0.0018 | 3.60E-20 | 1.27E-04 | 85.0 |  | -0.0126 | 0.0390 | 7.48E-01 |
| rs13227658 | C | T | 0.0157 | 0.0017 | 1.70E-19 | 1.22E-04 | 85.3 |  | -0.0060 | 0.0374 | 8.74E-01 |
| rs2733287 | C | A | 0.0157 | 0.0017 | 6.80E-20 | 1.23E-04 | 85.3 |  | 0.0014 | 0.0370 | 9.70E-01 |
| rs7243357 | T | G | 0.0194 | 0.0021 | 9.10E-20 | 1.08E-04 | 85.3 |  | 0.0711 | 0.0504 | 1.61E-01 |
| rs1927790 | C | T | 0.0148 | 0.0016 | 1.80E-19 | 1.06E-04 | 85.6 |  | 0.1009 | 0.0381 | 8.55E-03 |
| rs7730898 | A | G | 0.0168 | 0.0018 | 4.50E-20 | 1.12E-04 | 87.1 |  | -0.0073 | 0.0413 | 8.61E-01 |
| rs7332115 | T | G | 0.0159 | 0.0017 | 3.60E-21 | 1.18E-04 | 87.5 |  | 0.0150 | 0.0385 | 6.99E-01 |
| rs11074446 | T | C | 0.0225 | 0.0024 | 1.80E-20 | 1.15E-04 | 87.9 |  | 0.0200 | 0.0553 | 7.19E-01 |
| rs4757144 | A | G | 0.0169 | 0.0018 | 5.60E-22 | 1.38E-04 | 88.2 |  | -0.0467 | 0.0383 | 2.26E-01 |
| rs2694047 | G | A | 0.0188 | 0.002 | 3.90E-21 | 1.34E-04 | 88.4 |  | 0.0509 | 0.0424 | 2.33E-01 |
| rs1048932 | C | A | 0.016 | 0.0017 | 3.80E-22 | 1.24E-04 | 88.6 |  | 0.0244 | 0.0376 | 5.19E-01 |
| rs13191362 | A | G | 0.0236 | 0.0025 | 5.90E-21 | 1.17E-04 | 89.1 |  | 0.0285 | 0.0577 | 6.23E-01 |
| rs2010281 | G | A | 0.0161 | 0.0017 | 6.70E-21 | 1.19E-04 | 89.7 |  | 0.0205 | 0.0386 | 5.99E-01 |
| rs1477199 | G | A | 0.0228 | 0.0024 | 9.40E-22 | 1.29E-04 | 90.3 |  | 0.0579 | 0.0526 | 2.74E-01 |
| rs4483850 | A | T | 0.0162 | 0.0017 | 5.20E-21 | 1.31E-04 | 90.8 |  | 0.0025 | 0.0374 | 9.47E-01 |
| rs9989141 | T | C | 0.0162 | 0.0017 | 3.60E-21 | 1.21E-04 | 90.8 |  | 0.1113 | 0.0388 | 4.31E-03 |
| rs12150665 | T | C | 0.0162 | 0.0017 | 1.60E-22 | 1.27E-04 | 90.8 |  | -0.0259 | 0.0374 | 4.91E-01 |
| rs12468863 | C | T | 0.0153 | 0.0016 | 5.10E-21 | 1.17E-04 | 91.4 |  | -0.0525 | 0.0375 | 1.63E-01 |
| rs17724992 | A | G | 0.0183 | 0.0019 | 1.00E-22 | 1.29E-04 | 92.8 |  | 0.0252 | 0.0434 | 5.65E-01 |
| rs9538141 | A | G | 0.0164 | 0.0017 | 3.50E-21 | 1.34E-04 | 93.1 |  | 0.0054 | 0.0370 | 8.85E-01 |
| rs1528435 | T | C | 0.0164 | 0.0017 | 9.10E-23 | 1.25E-04 | 93.1 |  | 0.0547 | 0.0385 | 1.57E-01 |
| rs10920678 | A | G | 0.0155 | 0.0016 | 1.50E-21 | 1.18E-04 | 93.8 |  | 0.0641 | 0.0374 | 8.87E-02 |
| rs3800229 | T | G | 0.0175 | 0.0018 | 1.40E-22 | 1.26E-04 | 94.5 |  | 0.0484 | 0.0417 | 2.49E-01 |
| rs577525 | C | T | 0.0166 | 0.0017 | 9.70E-22 | 1.35E-04 | 95.3 |  | 0.1012 | 0.0379 | 8.03E-03 |
| rs1361739 | G | A | 0.0176 | 0.0018 | 6.90E-22 | 1.36E-04 | 95.6 |  | 0.0348 | 0.0403 | 3.90E-01 |
| rs7599312 | G | A | 0.0186 | 0.0019 | 6.90E-24 | 1.35E-04 | 95.8 |  | -0.0664 | 0.0425 | 1.20E-01 |
| rs1218822 | A | G | 0.0168 | 0.0017 | 1.90E-22 | 1.26E-04 | 97.7 |  | -0.0084 | 0.0396 | 8.32E-01 |
| rs11079849 | C | T | 0.0188 | 0.0019 | 4.80E-24 | 1.54E-04 | 97.9 |  | 0.0656 | 0.0400 | 1.03E-01 |
| rs11496125 | T | C | 0.0169 | 0.0017 | 3.00E-22 | 1.39E-04 | 98.8 |  | 0.0147 | 0.0375 | 6.96E-01 |
| rs977747 | T | G | 0.0169 | 0.0017 | 1.30E-24 | 1.38E-04 | 98.8 |  | 0.0347 | 0.0382 | 3.67E-01 |
| rs1296328 | A | C | 0.0179 | 0.0018 | 4.90E-24 | 1.57E-04 | 98.9 |  | 0.0276 | 0.0374 | 4.64E-01 |
| rs427943 | C | A | 0.017 | 0.0017 | 7.30E-23 | 1.42E-04 | 100.0 |  | -0.0226 | 0.0376 | 5.50E-01 |
| rs4482463 | C | A | 0.0331 | 0.0033 | 2.80E-23 | 1.59E-04 | 100.6 |  | 0.0785 | 0.0692 | 2.59E-01 |
| rs3803286 | A | G | 0.0181 | 0.0018 | 4.10E-23 | 1.48E-04 | 101.1 |  | 0.0340 | 0.0394 | 3.91E-01 |
| rs7903146 | C | T | 0.0181 | 0.0018 | 1.30E-23 | 1.35E-04 | 101.1 |  | -0.0894 | 0.0412 | 3.11E-02 |
| rs2481665 | T | C | 0.0161 | 0.0016 | 7.20E-23 | 1.28E-04 | 101.3 |  | -0.0632 | 0.0375 | 9.44E-02 |
| rs7084454 | A | G | 0.0193 | 0.0019 | 4.00E-25 | 1.66E-04 | 103.2 |  | -0.0764 | 0.0398 | 5.66E-02 |
| rs6870983 | C | T | 0.0204 | 0.002 | 6.50E-25 | 1.42E-04 | 104.0 |  | 0.1054 | 0.0467 | 2.50E-02 |
| rs7488867 | C | T | 0.0204 | 0.002 | 8.40E-24 | 1.62E-04 | 104.0 |  | 0.0265 | 0.0430 | 5.40E-01 |
| rs2365389 | C | T | 0.0174 | 0.0017 | 1.30E-25 | 1.47E-04 | 104.8 |  | 0.0419 | 0.0378 | 2.70E-01 |
| rs4986044 | C | T | 0.0164 | 0.0016 | 3.30E-23 | 1.34E-04 | 105.1 |  | -0.0115 | 0.0374 | 7.60E-01 |
| rs1431659 | A | G | 0.0196 | 0.0019 | 6.00E-24 | 1.50E-04 | 106.4 |  | -0.0084 | 0.0424 | 8.45E-01 |
| rs4516268 | C | A | 0.0217 | 0.0021 | 5.20E-25 | 1.46E-04 | 106.8 |  | 0.0789 | 0.0469 | 9.46E-02 |
| rs17513613 | C | T | 0.0186 | 0.0018 | 3.60E-26 | 1.51E-04 | 106.8 |  | -0.0050 | 0.0395 | 8.99E-01 |
| rs7715256 | G | T | 0.0166 | 0.0016 | 2.20E-24 | 1.34E-04 | 107.6 |  | 0.0080 | 0.0376 | 8.33E-01 |
| rs6985109 | G | A | 0.0177 | 0.0017 | 1.50E-26 | 1.56E-04 | 108.4 |  | -0.0297 | 0.0505 | 5.59E-01 |
| rs208015 | T | C | 0.0356 | 0.0034 | 1.40E-25 | 1.83E-04 | 109.6 |  | -0.1078 | 0.0738 | 1.47E-01 |
| rs16851483 | T | G | 0.0369 | 0.0035 | 3.20E-26 | 1.76E-04 | 111.2 |  | 0.0711 | 0.0755 | 3.49E-01 |
| rs2075650 | A | G | 0.0244 | 0.0023 | 1.50E-25 | 1.43E-04 | 112.5 |  | 0.0519 | 0.0527 | 3.27E-01 |
| rs4929923 | C | T | 0.0181 | 0.0017 | 7.20E-27 | 1.51E-04 | 113.4 |  | 0.0083 | 0.0387 | 8.32E-01 |
| rs7164727 | T | C | 0.0182 | 0.0017 | 3.30E-25 | 1.44E-04 | 114.6 |  | 0.0287 | 0.0402 | 4.77E-01 |
| rs12885454 | C | A | 0.0185 | 0.0017 | 2.40E-27 | 1.54E-04 | 118.4 |  | 0.0346 | 0.0386 | 3.73E-01 |
| rs657452 | A | G | 0.0188 | 0.0017 | 7.20E-29 | 1.66E-04 | 122.3 |  | 0.0033 | 0.0381 | 9.31E-01 |
| rs11880870 | A | G | 0.0189 | 0.0017 | 1.00E-28 | 1.78E-04 | 123.6 |  | 0.0398 | 0.0372 | 2.89E-01 |
| rs2245368 | C | T | 0.0257 | 0.0023 | 4.90E-28 | 1.90E-04 | 124.9 |  | -0.0382 | 0.0499 | 4.47E-01 |
| rs1579557 | T | C | 0.0213 | 0.0019 | 1.10E-29 | 1.90E-04 | 125.7 |  | 0.0713 | 0.0408 | 8.21E-02 |
| rs6545714 | G | A | 0.0191 | 0.0017 | 9.10E-31 | 1.73E-04 | 126.2 |  | 0.0581 | 0.0382 | 1.31E-01 |
| rs12939549 | A | G | 0.018 | 0.0016 | 2.70E-28 | 1.59E-04 | 126.6 |  | -0.0037 | 0.0374 | 9.22E-01 |
| rs1884897 | G | A | 0.0194 | 0.0017 | 1.30E-30 | 1.75E-04 | 130.2 |  | 0.0091 | 0.0388 | 8.16E-01 |
| rs7551507 | C | T | 0.0184 | 0.0016 | 9.30E-30 | 1.67E-04 | 132.3 |  | 0.0331 | 0.0378 | 3.84E-01 |
| rs40067 | G | A | 0.0266 | 0.0023 | 7.10E-30 | 2.01E-04 | 133.8 |  | 0.0376 | 0.0501 | 4.56E-01 |
| rs12369179 | C | T | 0.0359 | 0.0031 | 2.50E-31 | 2.06E-04 | 134.1 |  | 0.0556 | 0.0673 | 4.12E-01 |
| rs4740619 | T | C | 0.0186 | 0.0016 | 2.30E-30 | 1.71E-04 | 135.1 |  | 0.0232 | 0.0374 | 5.38E-01 |
| rs17806379 | C | T | 0.0258 | 0.0022 | 1.50E-30 | 1.96E-04 | 137.5 |  | 0.0737 | 0.0496 | 1.39E-01 |
| rs4237643 | T | G | 0.0223 | 0.0019 | 4.30E-33 | 2.11E-04 | 137.8 |  | 0.0296 | 0.0406 | 4.69E-01 |
| rs2122042 | T | G | 0.0235 | 0.002 | 2.30E-31 | 1.80E-04 | 138.1 |  | 0.0602 | 0.0458 | 1.92E-01 |
| rs1454687 | C | G | 0.0202 | 0.0017 | 5.20E-32 | 2.04E-04 | 141.2 |  | -0.0327 | 0.0373 | 3.84E-01 |
| rs4889606 | A | G | 0.0202 | 0.0017 | 2.80E-33 | 1.92E-04 | 141.2 |  | -0.0390 | 0.0379 | 3.06E-01 |
| rs12964689 | A | G | 0.0203 | 0.0017 | 5.10E-32 | 2.06E-04 | 142.6 |  | -0.0058 | 0.0375 | 8.77E-01 |
| rs17405819 | T | C | 0.0215 | 0.0018 | 4.30E-33 | 1.95E-04 | 142.7 |  | 0.0234 | 0.0404 | 5.65E-01 |
| rs1320903 | A | G | 0.0216 | 0.0018 | 9.20E-32 | 2.02E-04 | 144.0 |  | 0.0288 | 0.0402 | 4.77E-01 |
| rs11611246 | T | G | 0.024 | 0.002 | 5.00E-32 | 1.91E-04 | 144.0 |  | -0.0248 | 0.0458 | 5.91E-01 |
| rs11165643 | T | C | 0.0206 | 0.0017 | 1.40E-35 | 2.06E-04 | 146.8 |  | -0.0225 | 0.0379 | 5.55E-01 |
| rs889398 | C | T | 0.0196 | 0.0016 | 1.30E-32 | 1.88E-04 | 150.1 |  | 0.0656 | 0.0378 | 8.42E-02 |
| rs17207196 | C | T | 0.0221 | 0.0018 | 2.10E-35 | 2.37E-04 | 150.7 |  | 0.0117 | 0.0383 | 7.61E-01 |
| rs10132280 | C | A | 0.0223 | 0.0018 | 5.60E-35 | 2.10E-04 | 153.5 |  | -0.0008 | 0.0401 | 9.85E-01 |
| rs12429545 | A | G | 0.0316 | 0.0025 | 9.60E-38 | 2.18E-04 | 159.8 |  | -0.0022 | 0.0547 | 9.68E-01 |
| rs17391694 | T | C | 0.0317 | 0.0025 | 7.50E-38 | 2.11E-04 | 160.8 |  | 0.0876 | 0.0571 | 1.28E-01 |
| rs16903285 | C | T | 0.0331 | 0.0026 | 7.60E-38 | 2.65E-04 | 162.1 |  | 0.0147 | 0.0560 | 7.94E-01 |
| rs879620 | T | C | 0.0231 | 0.0018 | 5.30E-38 | 2.52E-04 | 164.7 |  | -0.0135 | 0.0384 | 7.26E-01 |
| rs4671328 | T | G | 0.0219 | 0.0017 | 2.20E-36 | 2.37E-04 | 166.0 |  | 0.0420 | 0.0376 | 2.66E-01 |
| rs2820311 | G | A | 0.0235 | 0.0018 | 4.10E-38 | 2.47E-04 | 170.4 |  | 0.0791 | 0.0389 | 4.34E-02 |
| rs3814883 | T | C | 0.0232 | 0.0017 | 1.10E-40 | 2.69E-04 | 186.2 |  | -0.0455 | 0.0373 | 2.26E-01 |
| rs2744974 | T | C | 0.0249 | 0.0018 | 1.40E-45 | 2.77E-04 | 191.4 |  | 0.0512 | 0.0399 | 2.03E-01 |
| rs11713193 | A | G | 0.0239 | 0.0017 | 2.40E-44 | 2.86E-04 | 197.7 |  | 0.0183 | 0.0372 | 6.24E-01 |
| rs7144011 | T | G | 0.0282 | 0.002 | 5.20E-47 | 2.67E-04 | 198.8 |  | 0.0091 | 0.0459 | 8.44E-01 |
| rs1412235 | C | G | 0.0246 | 0.0017 | 6.00E-45 | 2.62E-04 | 209.4 |  | -0.0479 | 0.0402 | 2.36E-01 |
| rs13329567 | C | T | 0.0293 | 0.002 | 1.00E-50 | 3.05E-04 | 214.6 |  | -0.1082 | 0.0452 | 1.74E-02 |
| rs12446632 | G | A | 0.0352 | 0.0024 | 2.90E-50 | 3.03E-04 | 215.1 |  | -0.0275 | 0.0544 | 6.15E-01 |
| rs13107325 | T | C | 0.047 | 0.0032 | 1.10E-47 | 3.02E-04 | 215.7 |  | -0.0029 | 0.0784 | 9.71E-01 |
| rs3810291 | A | G | 0.0274 | 0.0018 | 2.10E-52 | 3.32E-04 | 231.7 |  | -0.0481 | 0.0399 | 2.31E-01 |
| rs9816226 | T | A | 0.0323 | 0.0021 | 1.60E-52 | 3.08E-04 | 236.6 |  | -0.0158 | 0.0493 | 7.50E-01 |
| rs1993709 | G | A | 0.0331 | 0.0021 | 1.90E-57 | 3.27E-04 | 248.4 |  | 0.0901 | 0.0477 | 6.04E-02 |
| rs7498665 | G | A | 0.0271 | 0.0017 | 5.60E-60 | 3.54E-04 | 254.1 |  | -0.0165 | 0.0381 | 6.67E-01 |
| rs11672660 | C | T | 0.034 | 0.0021 | 1.70E-60 | 3.77E-04 | 262.1 |  | -0.0670 | 0.0449 | 1.38E-01 |
| rs7124681 | A | C | 0.0263 | 0.0016 | 3.20E-58 | 3.35E-04 | 270.2 |  | -0.0202 | 0.0382 | 5.99E-01 |
| rs2307111 | T | C | 0.0265 | 0.0016 | 1.60E-58 | 3.36E-04 | 274.3 |  | 0.0952 | 0.0382 | 1.33E-02 |
| rs7138803 | A | G | 0.03 | 0.0017 | 2.30E-71 | 4.23E-04 | 311.4 |  | 0.0204 | 0.0380 | 5.94E-01 |
| rs987237 | G | A | 0.0409 | 0.0021 | 9.30E-84 | 4.94E-04 | 379.3 |  | 0.0311 | 0.0477 | 5.16E-01 |
| rs6265 | C | T | 0.0412 | 0.0021 | 1.00E-86 | 5.33E-04 | 384.9 |  | 0.0489 | 0.0470 | 3.01E-01 |
| rs10938397 | G | A | 0.0324 | 0.0016 | 3.40E-86 | 5.15E-04 | 410.1 |  | -0.0167 | 0.0380 | 6.61E-01 |
| rs10182181 | G | A | 0.0325 | 0.0016 | 6.70E-90 | 5.27E-04 | 412.6 |  | 0.0072 | 0.0373 | 8.49E-01 |
| rs543874 | G | A | 0.0475 | 0.002 | 1.20E-122 | 7.09E-04 | 564.1 |  | -0.0031 | 0.0455 | 9.46E-01 |
| rs13021737 | G | A | 0.0574 | 0.0021 | 7.50E-157 | 9.21E-04 | 747.1 |  | -0.0171 | 0.0493 | 7.30E-01 |
| rs663129 | A | G | 0.0545 | 0.0019 | 1.60E-178 | 1.05E-03 | 822.8 |  | 0.0856 | 0.0434 | 5.00E-02 |
| rs8047395 | A | G | 0.0642 | 0.0017 | 1.00E-305 | 2.06E-03 | 1426.2 |  | 0.0783 | 0.0370 | 3.56E-02 |

LADA: latent autoimmune diabetes in adults.

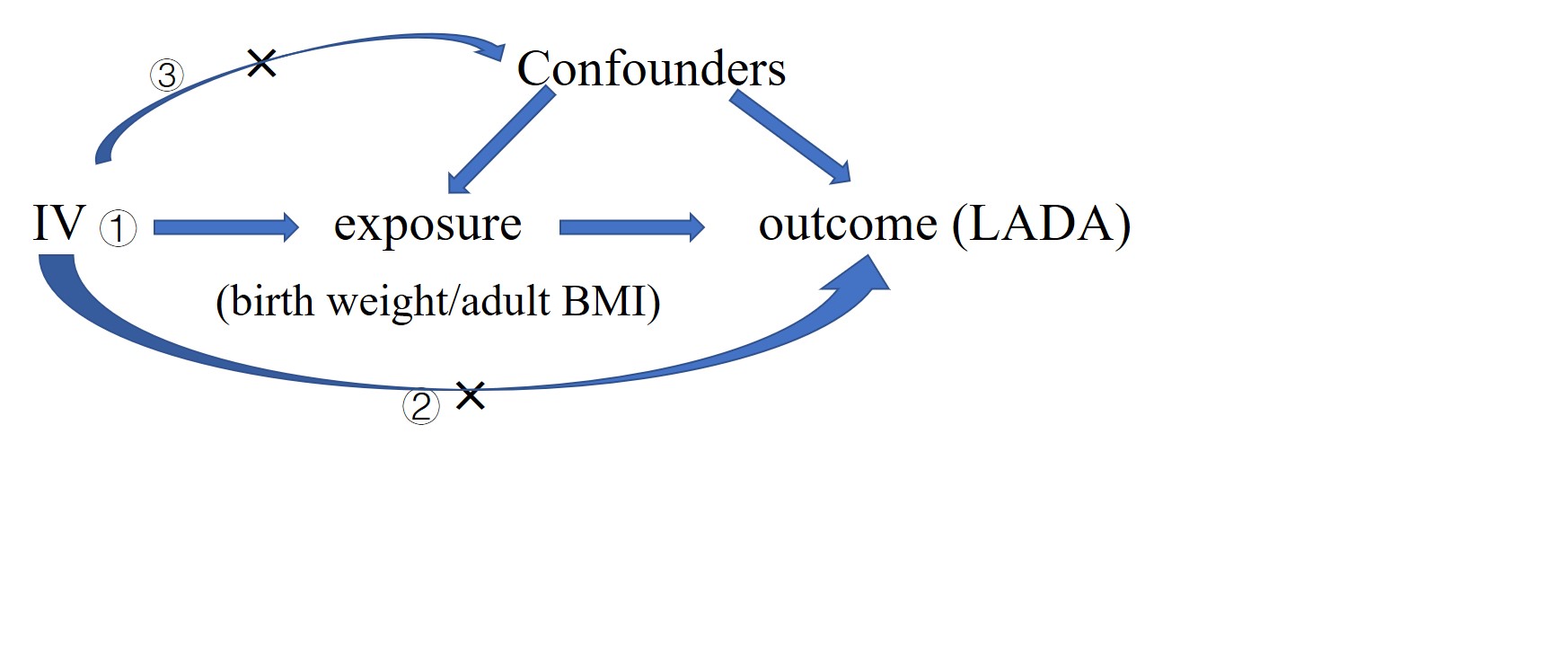

### eFigure 1. IV assumptions

IV: instrumental variable; LADA: latent autoimmune diabetes in adults.

IV can only affect the outcome through the exposure (①), not through a direct pathway (②) to the outcome or via a confounder (③).

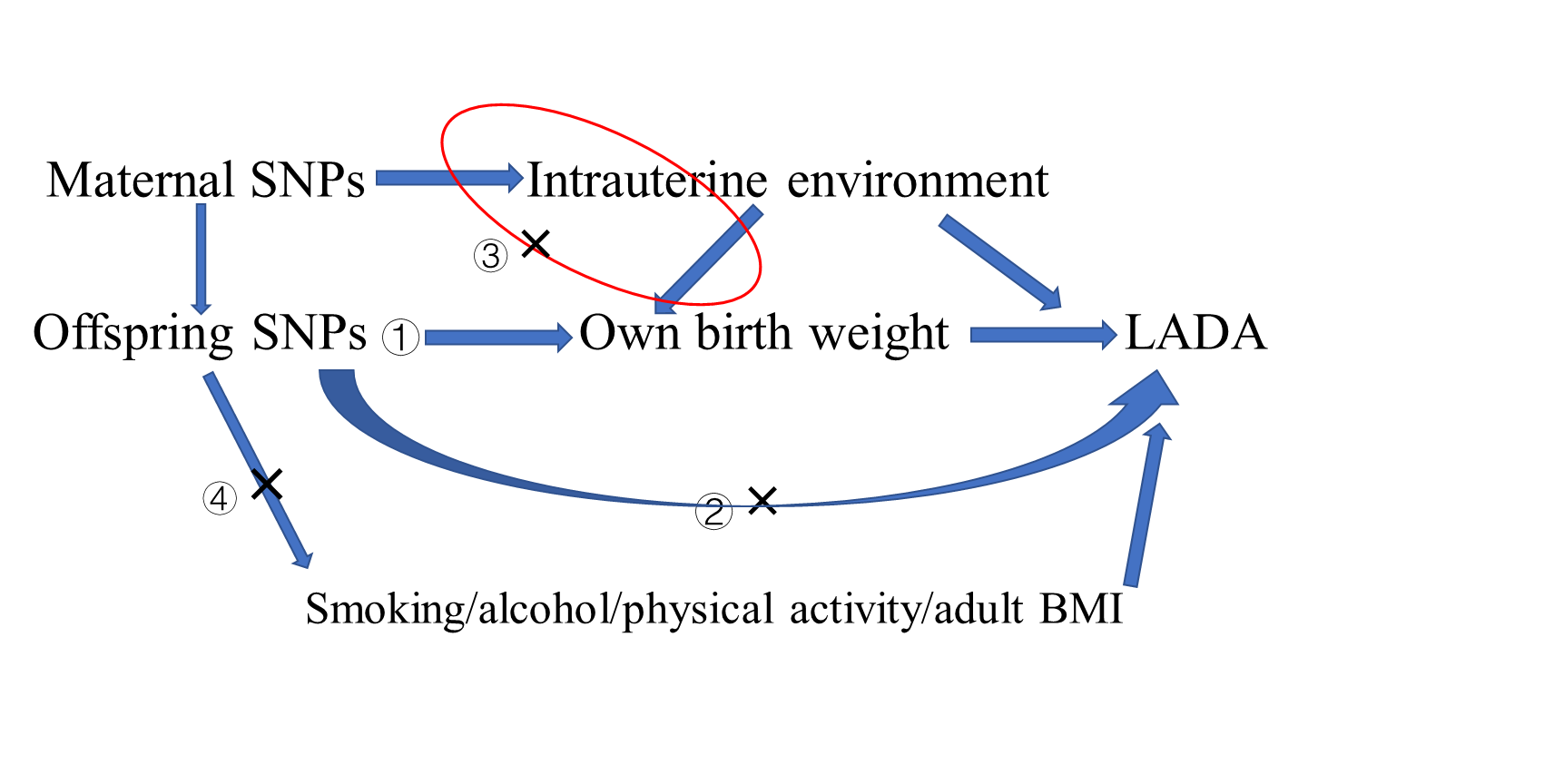

**(A)**

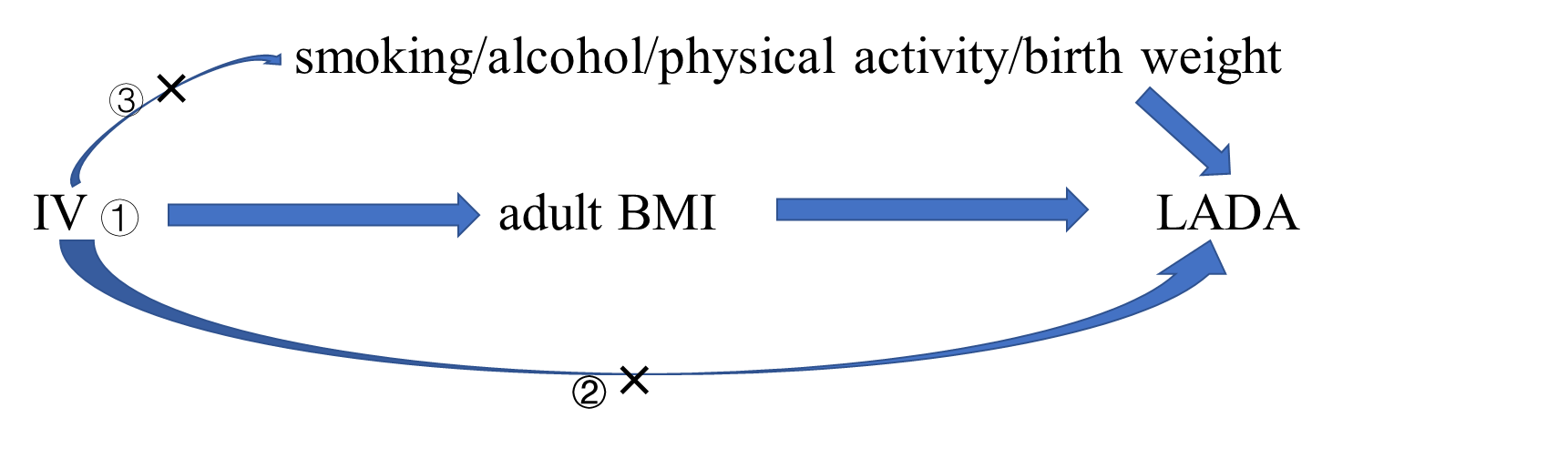

**(B)**

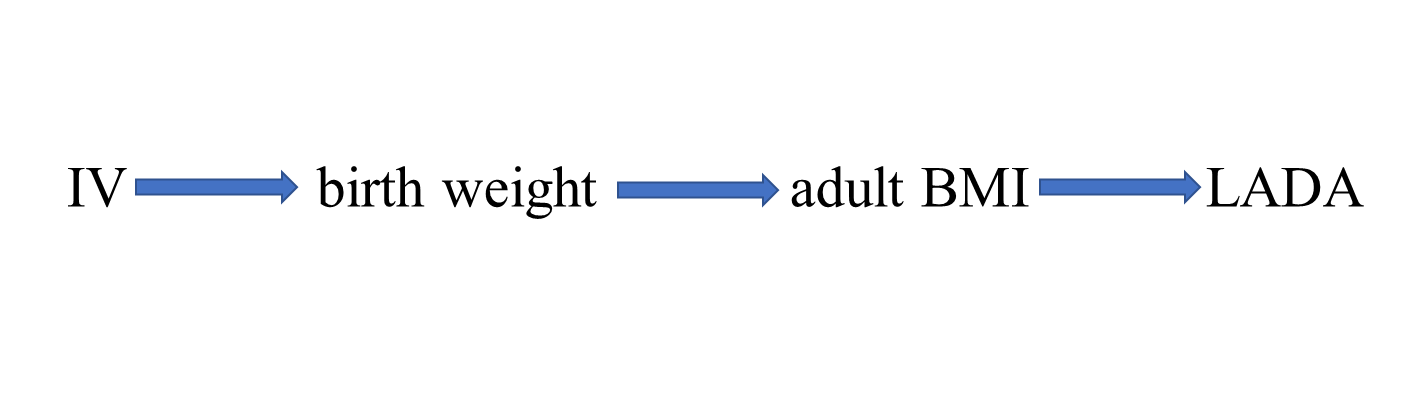

**(C)**

### eFigure 2. Directed acyclic graphs showing reasons for conservative analyses

IV: instrumental variable; LADA: latent autoimmune diabetes in adults.

(A) The IV for birth weight should not affect LADA directly (pathway ②), or through maternal confounders (pathway ③), or through other pathways (pathway ④). (B) The IV for adult BMI should not affect LADA directly (pathway ②) or through other pathways (pathway ③). (C) The IV might affect birth weight and adult BMI through the same pathway and there is no violation of IV assumption in this scenario.

### eTable 4. SNPs excluded from conservative analyses

| **Exposures** | **Excluded instruments** | **Reasons for exclusion** |
| --- | --- | --- |
| **Birth weight** |  |  |
| **Conservative analysis 1** | SNPs with maternal-only effects, or both maternal and fetal effects on birth weight, or unclassified SNPs **^a^**. | Only SNPs with fetal-only effects on birth weight were included as IVs. Fetal-only effects mean that these SNPs only affect own birth weight and maternal SNPs do not affect birth weight. This conservative analysis is to minimize the possibility that IVs affect LADA through maternal confounding factors (intrauterine environment, pathway ③ in **eFigure 2A**). |
| **Conservative analysis 2 ^a^** | SNPs excluded from conservative analysis 1, and SNPs associated with diabetes-related traits (any type of diabetes, 2 hour fasting glucose, HbA1c, insulin, and so on) at *P*<0.0003 (0.05/129). | To minimize the possibility that IVs affect LADA directly (pathway ② in **eFigure 2A**). |
| **Conservative analysis 3 ^a^** | SNPs excluded from conservative analysis 2, and SNPs associated with any trait (except body size-related traits at birth) at *P*<5×10^-8^. | To further minimize the possibility of pleiotropy. |
| **Conservative analysis 4 ^a^** | SNPs excluded from conservative analysis 3 and SNPs associated with smoking, alcohol, or physical activity at *P*<0.0003 (0.05/129). | To minimize the possibility that IVs affect LADA through lifestyle factors (pathway ④ in eFigure 2A). |
| **Conservative analysis 5 ^a^** | SNPs excluded from conservative analysis 4, and SNPs associated with adult body size at *P*<0.0003 (0.05/129). | Birth weight and adult BMI are both anthropometric measurements. IVs associated with both birth weight and adult BMI might be associated with LADA through the same pathway (**eFigure 2C**, no violation of IV assumption) or through two different pathways (pathway ④ in **eFigure 2A**, violation of IV assumption). This conservative analysis is to minimize the violation of IV assumption as shown in pathway ④ in **eFigure 2A**. |
| **BMI in adulthood** |  |  |
| **Conservative analysis 1 ^a^** | SNPs associated with diabetes-related traits (any type of diabetes, 2 hour fasting glucose, HbA1c, insulin, and so on) at *P*<6×10^-5^ (0.05/820) or any trait (except body size-related traits in adulthood) at *P*<5×10^-8^. | Excluding SNPs associated with diabetes-related traits is to reduce the possibility that IVs affect LADA directly (pathway ② in **eFigure 2B**). |
| **Conservative analysis 2 ^a^** | SNPs excluded from conservative analysis 1, and SNPs associated with smoking, alcohol, or physical activity at *P*<6×10^-5^ (0.05/820). | To reduce the possibility that IVs for adult BMI affect LADA through lifestyle factors (pathway ③ in **eFigure 2B**). |
| **Conservative analysis 3 ^a^** | SNPs excluded from conservative analysis 2, and SNPs associated with body size-related traits at birth at *P*<6×10^-5^ (0.05/820). | To minimize the violation of IV assumption as shown in pathway ③ in **eFigure 2B**. |

^a^ The association between SNPs and traits were identified from Phenoscanner[3,4].

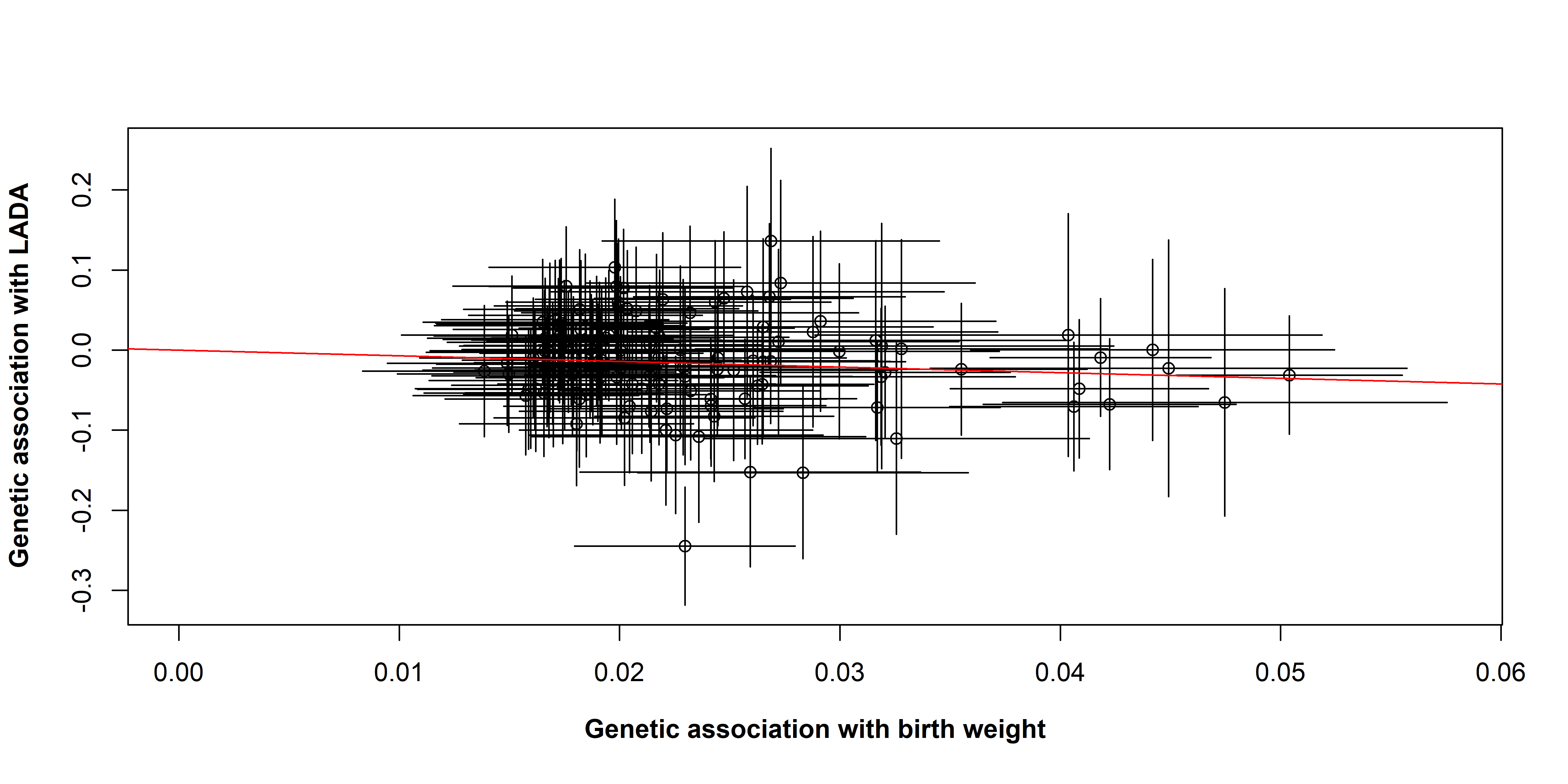

### eFigure 3. Scatter plot for associations of 129 SNPs with birth weight and LADA.

LADA: latent autoimmune diabetes in adults. The slope of the red line is the log OR of increase in LADA risk one SD (0.5 kg) increase in birth weight, based on the inverse-variance weighted method.

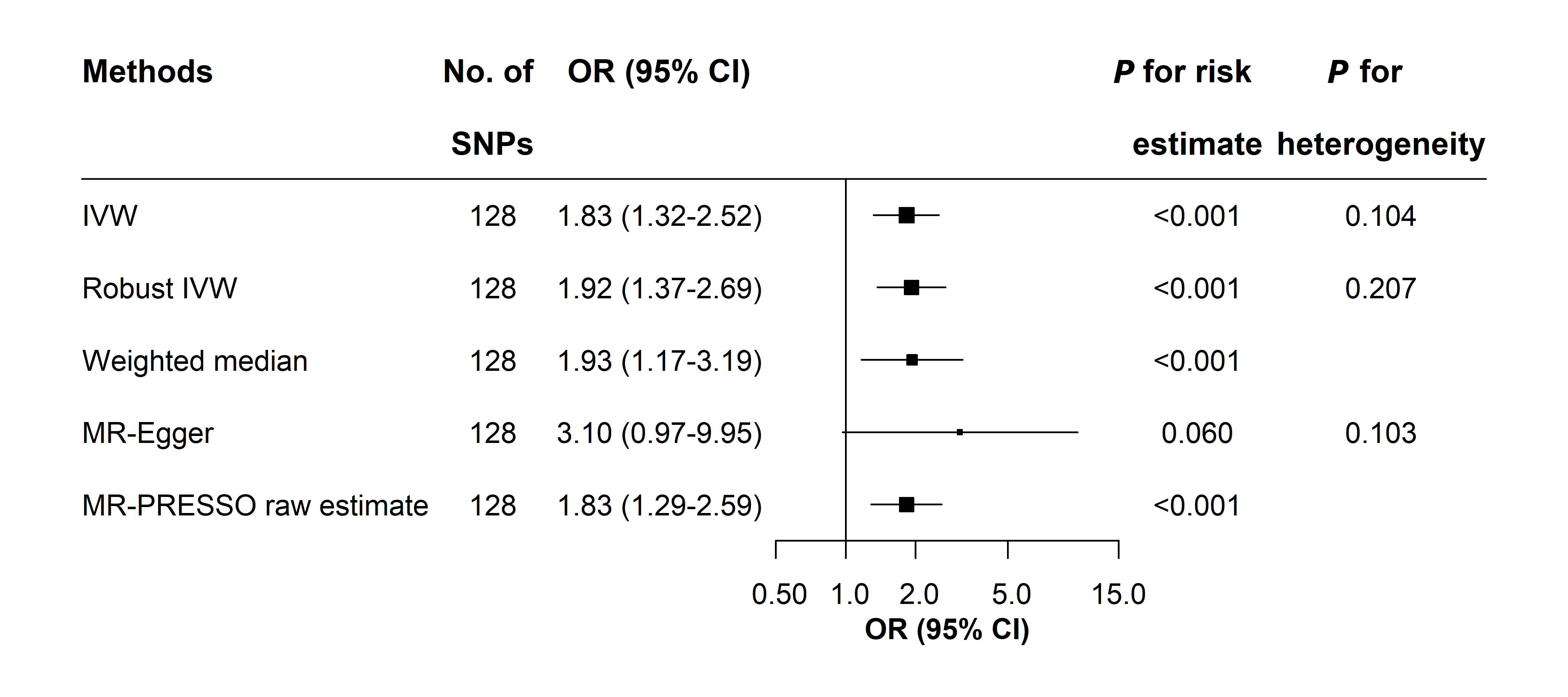

### eFigure 4. The risk of LADA in relation to one SD (0.5 kg) decrease in birth weight based on 128 SNPs

IVW: inverse-variance weighted. MR-Egger: Egger regression of Mendelian randomization; MR-PRESSO: the Mendelian randomization pleiotropy residual sum and outlier approach; LADA: latent autoimmune diabetes in adults.

MR-Egger intercept: -0.013, *P* for directional pleiotropy: 0.351.

MR-PRESSO detected no outlier and the raw estimate was reported.

### eTable 5. Conservative analyses for the association of birth weight and adult BMI with LADA

| **Exposures** | **Conservative**  **analyses ^a^** | **No. of excluded SNPs** | **No. of included SNPs** | **OR (95% CI) ^j^** | ***P* for risk estimate** | ***P* for heterogeneity** |
| --- | --- | --- | --- | --- | --- | --- |
| **Birth weight** | 1 ^b^ | 74 | 55 | 1.83 (1.15-2.91) | 0.010 | 0.643 |
|  | 2 ^c^ | 84 | 45 | 1.81 (1.03-3.17) | 0.038 | 0.567 |
|  | 3 ^d^ | 110 | 19 | 1.76 (0.71-4.32) | 0.220 | 0.949 |
|  | 4 ^e^ | 110 | 19 | 1.76 (0.71-4.32) | 0.220 | 0.949 |
|  | 5 ^f^ | 114 | 15 | 1.99 (0.71-5.55) | 0.189 | 0.900 |
| **BMI in adulthood** | 1 ^g^ | 364 | 456 | 1.56 (1.16-2.09) | 0.003 | 0.815 |
|  | 2 ^h^ | 396 | 424 | 1.57 (1.16-2.14) | 0.004 | 0.909 |
|  | 3 ^i^ | 399 | 421 | 1.59 (1.17-2.16) | 0.003 | 0.894 |

LADA latent autoimmune diabetes in adults.

^a^ The method of inverse-variance weighted was used in conservative analyses.

^b^ Conservative analysis 1 only included SNPs with fetal-only effects as IVs.

^c^ Conservative analysis 2 further excluded SNPs associated with diabetes-related traits at *P*<0.0003 (0.05/129).

^d^ Conservative analysis 3 further excluded SNPs associated with any trait (except birth weight) at *P*<5×10^-8^.

^e^ Conservative analysis 4 further excluded SNPs associated with smoking, alcohol or physical activity at *P*<0.0003 (0.05/129).

^f^ Conservative analysis 5 further excluded SNPs associated with body size in adulthood at *P*<0.0003 (0.05/129).

^g^ Conservative analysis 1 excluded SNPs associated with diabetes-related traits (any type of diabetes, 2 hour fasting glucose, HbA1c, insulin, and so on) at *P*<6×10^-5^ (0.05/820), or associated with any trait (except body size in adulthood) at *P*<5×10^-8^.

^h^ Conservative analysis 2 further excluded SNPs associate with smoking, alcohol, or physical activity at *P*<6×10^-5^.

^i^ Conservative analysis 3 further excluded SNPs associated with body size -related traits at birth at *P*<6×10^-5^.

^j^ OR (95% CI) for one SD (0.5 kg) decrease in birth weight, or one SD (4.8 kg/m^2^) increase in BMI in adulthood.

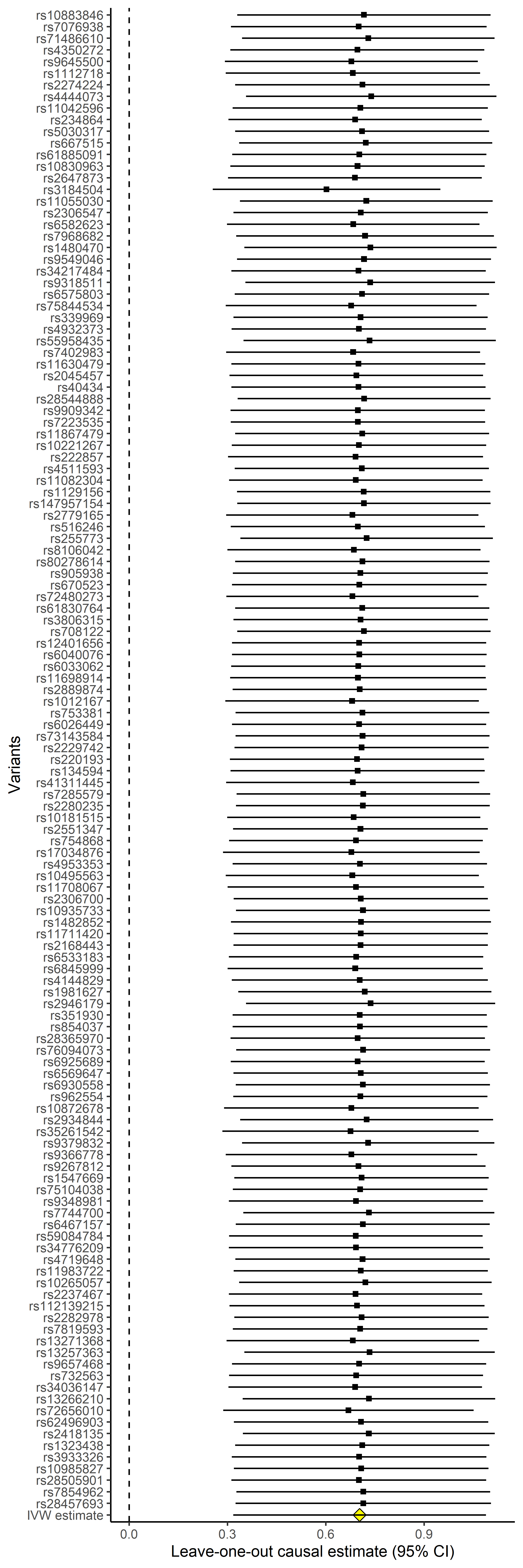

### eFigure 5. Leave-one-out analysis for risk of LADA one SD decrease in genetically determined birth weight

LADA: latent autoimmune diabetes in adults.

Each SNP included in the main analysis was left out in turn, leaving 128 SNPs in each causal estimate. Causal estimate was presented as log ORs and corresponding 95% CIs.

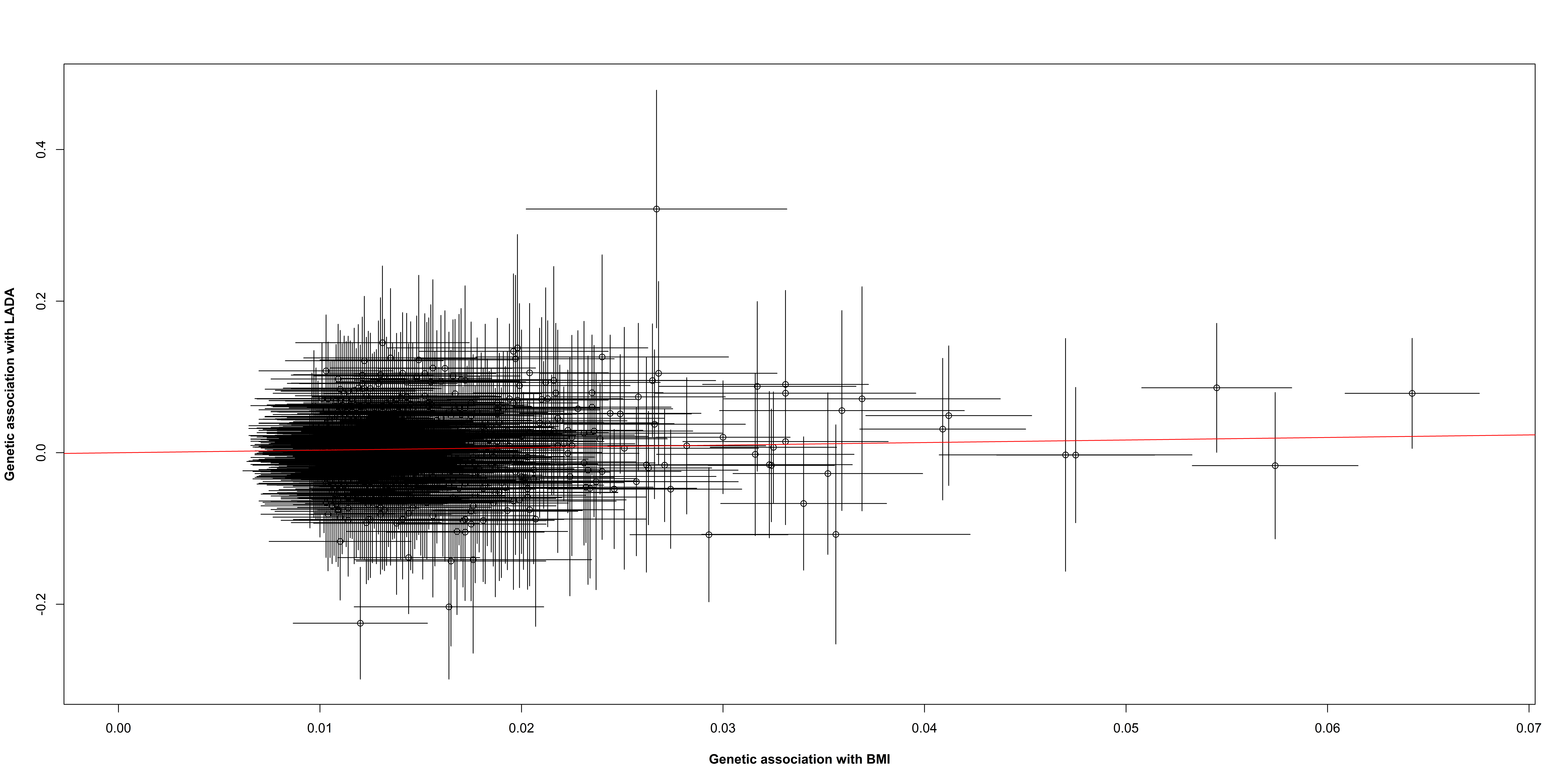

rs11066188

rs10840606

### eFigure 6. Scatter plot for associations of 820 SNPs with BMI and LADA

LADA: latent autoimmune diabetes in adults.

The slope of the red line is the log OR of increase in LADA risk one SD (4.8 kg/m^2^) increase in BMI, based on the inverse-variance weighted method.

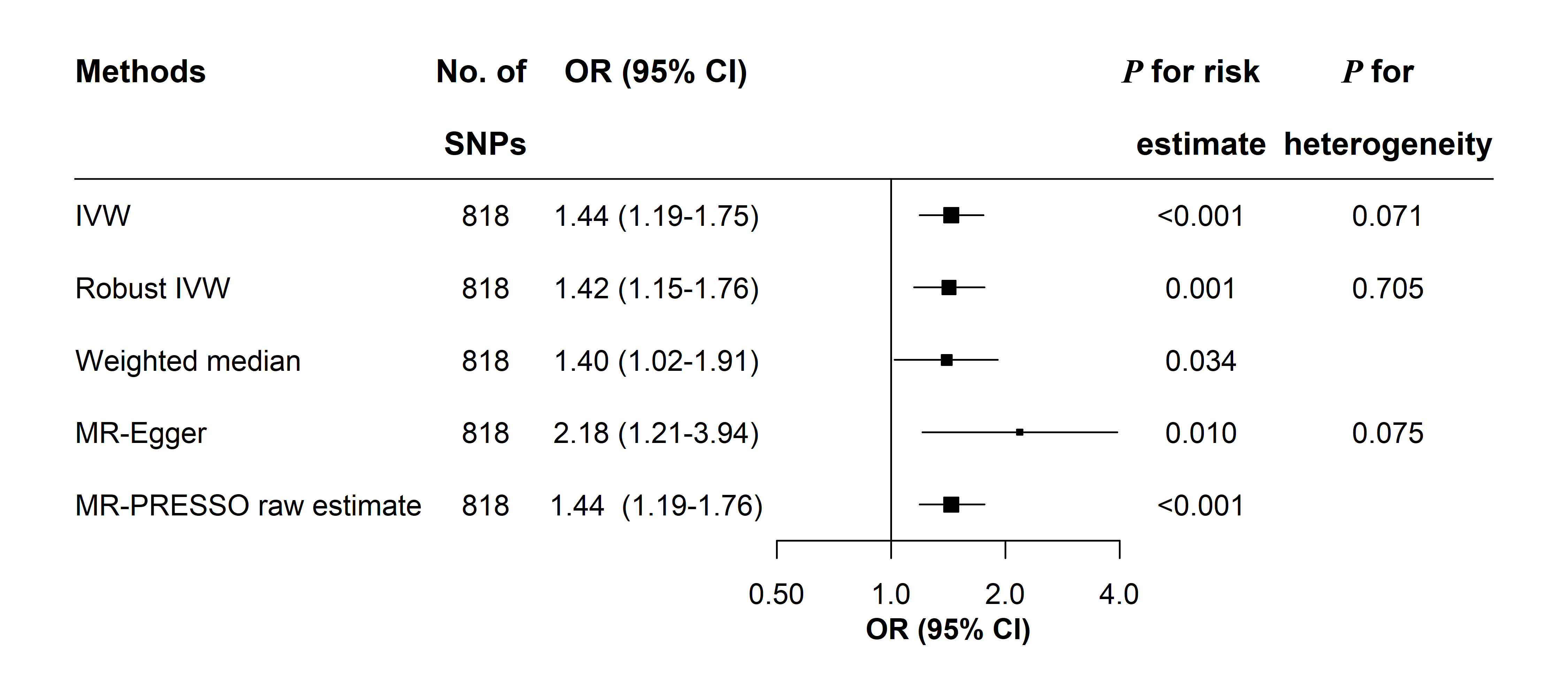

### eFigure 7. The risk of LADA in relation to one SD (0.5 kg) increase in adult BMI based on 818 SNPs

IVW: inverse-variance weighted. MR-Egger: Egger regression of Mendelian randomization; MR-PRESSO: the Mendelian randomization pleiotropy residual sum and outlier approach; LADA: latent autoimmune diabetes in adults.

MR-Egger intercept: -0.007, *P* for directional pleiotropy: 0.149.

MR-PRESSO detected no outlier and the raw estimate was reported.

### eTable 6. Risk of type 2 diabetes in relation to birth weight according to different methods

| **Methods** | **No. of SNPs** | **OR (95% CI)** | ***P* for risk estimate** | ***P* for heterogeneity** |
| --- | --- | --- | --- | --- |
| **IVW** | 129 | 1.76 (1.38-2.23) | <0.001 | <0.001 |
| **Robust IVW** | 129 | 1.44 (1.19-1.74) | <0.001 | <0.001 |
| **Weighted Median** | 129 | 1.49 (1.22-1.83) | <0.001 |  |
| **MR-Egger ^a^** | 129 | 3.36 (1.49-7.54) | 0.003 | <0.001 |
| **MR-PRESSO outlier-corrected ^b^** | 119 | 1.45 (1.25-1.70) | <0.001 |  |

IVW: inverse-variance weighted. MR-Egger: Egger regression of Mendelian randomization; MR-PRESSO: the Mendelian randomization pleiotropy residual sum and outlier approach.

^a^ MR-Egger intercept: -0.015, P for directional pleiotropy: 0.101.

^b^ MR-PRESSO detected rs71486610, rs1112718, rs234864, rs10830963, rs40434, rs222857, rs11708067, rs6925689, rs35261542, rs13266210 as outliers. *P* for distortion of risk estimate by outliers was 0.003. Outliers were excluded from the outlier-corrected estimate.

### eTable 7. Risk of LADA and type 2 diabetes in relation to adult BMI according to different methods using SNPs from UK Biobank

| **Methods** | **No. of SNPs** | **OR (95% CI)** | ***P* for risk estimate** | ***P* for heterogeneity** |
| --- | --- | --- | --- | --- |
| **LADA** |  |  |  |  |
| IVW | 734 | 1.45 (1.18-1.79) | <0.001 | <0.001 |
| Robust IVW | 734 | 1.41 (1.14-1.74) | 0.002 | 0.438 |
| Weighted Median | 734 | 1.51 (1.11-2.04) | 0.008 |  |
| MR-Egger ^a^ | 734 | 2.69 (1.40-5.18) | 0.003 | <0.001 |
| MR-PRESSO outlier-corrected ^b^ | 731 | 1.42 (1.18-1.72) | <0.001 |  |
| **Type 2 diabetes** |  |  |  |  |
| IVW | 734 | 2.26 (2.06-2.49) | <0.001 | <0.001 |
| Robust IVW | 734 | 2.37 (2.19-2.56) | <0.001 | <0.001 |
| Weighted Median | 734 | 2.63 (2.37-2.92) | <0.001 |  |
| MR-Egger ^c^ | 734 | 3.32 (2.47-4.47) | <0.001 | <0.001 |
| MR-PRESSO outlier-corrected ^d^ | 725 | 2.33 (2.16-2.51) | <0.001 |  |

LADA: latent autoimmune diabetes in adults; IVW: inverse-variance weighted. MR-Egger: Egger regression of Mendelian randomization; MR-PRESSO: the Mendelian randomization pleiotropy residual sum and outlier approach.

^a^ MR-Egger intercept: -0.011, *P* for directional pleiotropy: 0.05.

^b^ MR-PRESSO detected rs10840606, rs2271189, and rs1046080 as outliers. Outliers were excluded from the outlier-corrected estimate (*P* for distortion of estimate by outliers: 0.816).

^c^ MR-Egger intercept: -0.007, *P* for directional pleiotropy: 0.007.

^d^ MR-PRESSO detected rs36090025, rs1002226, rs56094641, rs429358, rs10423928, rs61791109, rs329118, rs9366863, and rs849133 as outliers. *P* for distortion of risk estimate by outliers was 0.457. Outliers were excluded from the outlier-corrected estimate.
